# Multi-organ Proteomic Landscape of COVID-19 Autopsies

**DOI:** 10.1101/2020.08.16.20176065

**Authors:** Xiu Nie, Liujia Qian, Rui Sun, Bo Huang, Xiaochuan Dong, Qi Xiao, Qiushi Zhang, Tian Lu, Liang Yue, Shuo Chen, Xiang Li, Yaoting Sun, Lu Li, Luang Xu, Yan Li, Ming Yang, Zhangzhi Xue, Shuang Liang, Xuan Ding, Chunhui Yuan, Li Peng, Wei Liu, Xiao Yi, Mengge Lyu, Guixiang Xiao, Xia Xu, Weigang Ge, Jiale He, Jun Fan, Junhua Wu, Meng Luo, Xiaona Chang, Huaxiong Pan, Xue Cai, Junjie Zhou, Jing Yu, Huanhuan Gao, Mingxing Xie, Sihua Wang, Guan Ruan, Hao Chen, Hua Su, Heng Mei, Danju Luo, Dashi Zhao, Fei Xu, Yan Li, Yi Zhu, Jiahong Xia, Yu Hu, Tiannan Guo

**Affiliations:** Department of Pathology, Union Hospital, Tongji Medical College, Huazhong University of Science and Technology, Wuhan 430022, China; Zhejiang Provincial Laboratory of Life Sciences and Biomedicine, Key Laboratory of Structural Biology of Zhejiang Province, School of Life Sciences, Westlake University, Hangzhou 310024, China; Institute of Basic Medical Sciences, Westlake Institute for Advanced Study, Hangzhou 310024, China; Department of Anatomy, College of Basic Medical Sciences, Dalian Medical University, Dalian 116044, China; Department of Ultrasound, Union Hospital, Tongji Medical College, Huazhong University of Science and Technology, Wuhan 430022, China; Department of Thoracic Surgery, Union Hospital, Tongji Medical College, Huazhong University of Science and Technology, Wuhan 430022, China; Department of Nephrology, Union Hospital, Tongji Medical College, Huazhong University of Science and Technology, Wuhan 430022, China; Institute of Hematology, Union Hospital, Tongji Medical College, Huazhong University of Science and Technology, Wuhan 430022, China; Department of Cardiovascular Surgery, Union Hospital, Tongji Medical College, Huazhong University of Science and Technology, Wuhan 430022, China; Department of Anatomy and Physiology, College of Basic Medical Sciences, Shanghai Jiao Tong University, Shanghai, 200025, China; Lead contact

**Author notes:** These authors contribute equally (Xiu Nie, Liujia Qian, Rui Sun, Bo Huang, Xiaochuan Dong, Qi Xiao, Qiushi Zhang). Correspondence (Y.Z.); (H.X.); (Y.H.); (T.G.).

## Abstract

The molecular pathology of multi-organ injuries in COVID-19 patients remains unclear, preventing effective therapeutics development. Here, we report an in-depth multi-organ proteomic landscape of COVID-19 patient autopsy samples. By integrative analysis of proteomes of seven organs, namely lung, spleen, liver, heart, kidney, thyroid and testis, we characterized 11,394 proteins, in which 5336 were perturbed in COVID-19 patients compared to controls. Our data showed that CTSL, rather than ACE2, was significantly upregulated in the lung from COVID-19 patients. Dysregulation of protein translation, glucose metabolism, fatty acid metabolism was detected in multiple organs. Our data suggested upon SARS-CoV-2 infection, hyperinflammation might be triggered which in turn induces damage of gas exchange barrier in the lung, leading to hypoxia, angiogenesis, coagulation and fibrosis in the lung, kidney, spleen, liver, heart and thyroid. Evidence for testicular injuries included reduced Leydig cells, suppressed cholesterol biosynthesis and sperm mobility. In summary, this study depicts the multi-organ proteomic landscape of COVID-19 autopsies, and uncovered dysregulated proteins and biological processes, offering novel therapeutic clues.

**HIGHLIGHTS:**

- Characterization of 5336 regulated proteins out of 11,394 quantified proteins in the lung, spleen, liver, kidney, heart, thyroid and testis autopsies from 19 patients died from COVID-19.
- CTSL, rather than ACE2, was significantly upregulated in the lung from COVID-19 patients.
- Evidence for suppression of glucose metabolism in the spleen, liver and kidney; suppression of fatty acid metabolism in the kidney; enhanced fatty acid metabolism in the lung, spleen, liver, heart and thyroid from COVID-19 patients; enhanced protein translation initiation in the lung, liver, renal medulla and thyroid.
- Tentative model for multi-organ injuries in patients died from COVID-19: SARS-CoV-2 infection triggers hyperinflammatory which in turn induces damage of gas exchange barrier in the lung, leading to hypoxia, angiogenesis, coagulation and fibrosis in the lung, kidney, spleen, liver, heart, kidney and thyroid.
- Testicular injuries in COVID-19 patients included reduced Leydig cells, suppressed cholesterol biosynthesis and sperm mobility.

## INTRODUCTION

The ongoing COVID-19 pandemic, caused by severe acute respiratory syndrome coronavirus 2 (SARS-CoV-2), has led to more than 20 million infected individuals and over 700,000 deaths by the middle of August 2020. Morphological characterization of autopsies, mainly focused on the pulmonary lesions, has greatly advanced our understanding of COVID-19-caused deaths (Carsana et al., 2020; Su et al., 2020a; Wichmann et al., 2020; Wu et al., 2020; Xu et al., 2020; Yao et al., 2020). Mechanistic studies of SARS-CoV-2 infected cell line models (Bojkova et al., 2020; Bouhaddou et al., 2020; Gordon et al., 2020) offer new insights into virus-perturbed biochemical processes of COVID-19 and suggest potentially novel therapies. SARS-CoV-2 infected mouse models (Bao et al., 2020; Hassan et al., 2020; Jiang et al., 2020) and rhesus macaque models (Chandrashekar et al., 2020; Deng et al., 2020) generated by adenovirus transduction of human ACE2 have been established for preclinical selection of antiviral therapeutic agents and vaccines as well as for investigating pathogenesis. Few studies have characterized host responses at molecular level from clinical specimens. We and others have studied the host responses by proteomic and metabolomic analysis of patient sera (Messner et al., 2020; Shen et al., 2020a), but molecular changes in infected tissues and consequentially affected organs remain elusive. To date, little knowledge has been obtained concerning how SARS-CoV-2 virus induces injuries in multiple organs (Bian, 2020; Tian et al., 2020; Wichmann et al., 2020) including lung, kidney (Kudose et al., 2020), liver, heart, spleen, thyroid and testis (Yang et al., 2020), and how to prevent and revert them.

Maximilian et. al analyzed the mRNA expression of autopsies of seven lungs from COVID-19 patients and reported intussusceptive angiogenesis which may be induced by hypoxemia (Ackermann et al., 2020). Latest advances of proteomics technologies allow effective and robust analysis of formalin-fixed tissue samples (Gao et al., 2020; Zhu et al., 2019). Comparative analysis of mRNA and protein expression in tissue samples showed that proteins measured by mass spectrometry was much more stable than transcripts (Shao et al., 2019). Here, we report a multi-organ proteomic profiling of 144 autopsy tissue samples collected from the lung, spleen, liver, heart, kidney, thyroid and testis of 19 patients died from COVID-19 and 74 control tissue samples from 56 non-COVID-19 patients. Using TMT-based shotgun proteomics, we quantified 11,394 proteins, out of which 5336 were significantly dysregulated proteins in at least one organ in COVID-19 patients. This data resource offers a unique channel to understand multi-organ injuries in the COVID-19 patients, and nominates potential therapeutics.

## RESULTS AND DISCUSSION

### Generation and characterization of proteomic landscape

We first performed proteomic profiling of 144 autopsy tissue samples from seven types of organs, namely lung (N = 15, n = 30; N represents the patient number, n represents sample number), spleen (N = 9, n = 9 from white pulp and N = 8, n = 8 from red pulp), liver (N = 10, n = 24), kidney (N = 10, n = 18 from renal cortex and N = 9, n = 16 from renal medulla), heart (N = 9, n = 19), testis (N = 5, n = 5) and thyroid (N = 15, n = 15). The samples were from 19 COVID-19 cases, ten of which have been described previously (Wu et al., 2020), compared with 74 control samples from 56 non-COVID-19 cases with other diseases via surgeries (Figure 1A, Tables S1,2). All 19 COVID-19 patients died from SARS-CoV-2 pneumonia or respiratory failure, among whom seven also developed terminal multi-organ failure. Detailed information of patients including medication history during hospitalization, laboratory test data, pathological changes, and cause of death are summarized in Tables S1, 5 and Figure S1, 2.

**Figure 1.**
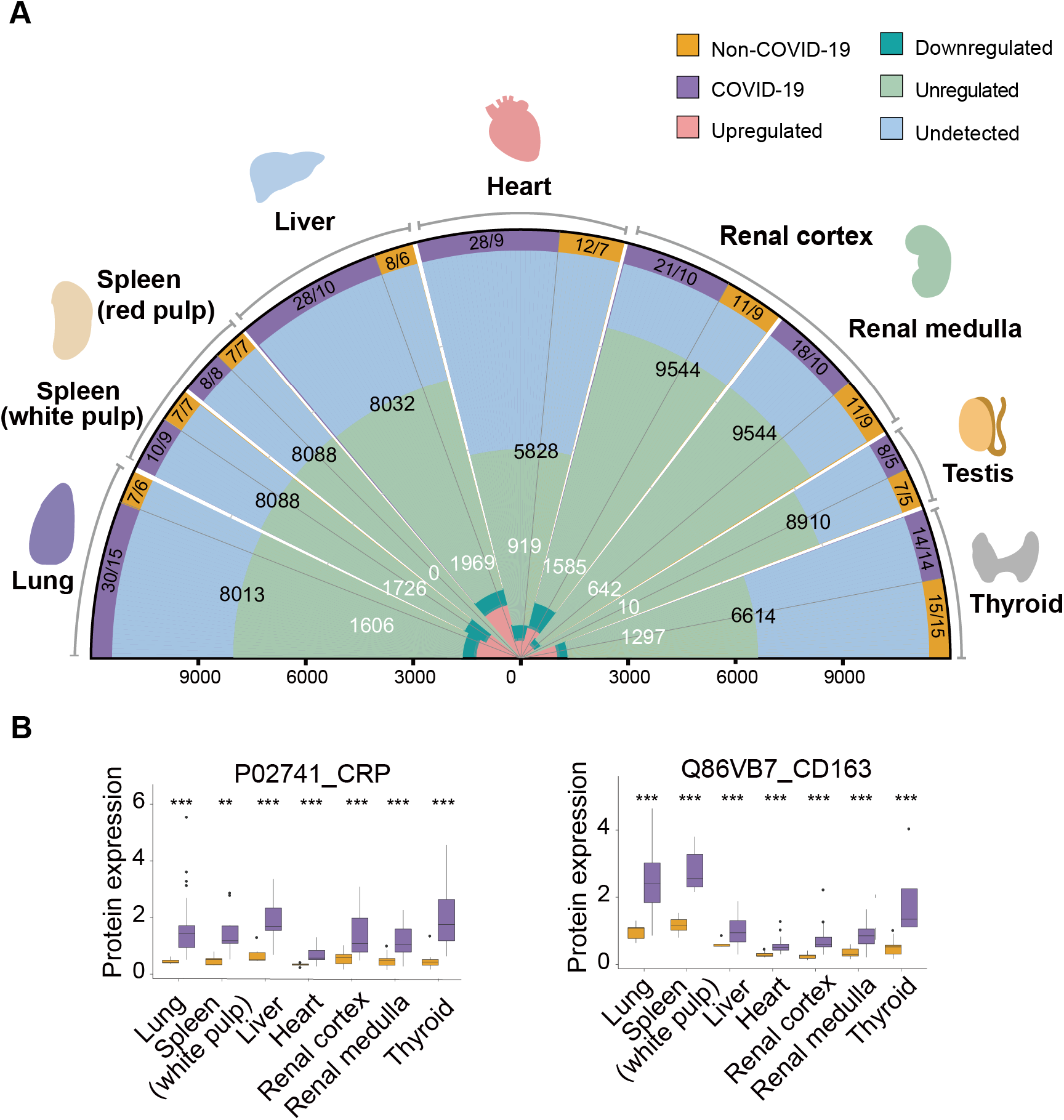
Multi-organ proteomic landscape of COVID-19 autopsies. **A**. The quantified and dysregulated proteins across multiple organs. The outermost (first) ring represents the type of samples. The number of samples and patients (n/N) are labeled respectively. The second ring (in blue) refers to the missing/undetected proteins for each organ. The numbers in black represent the quantified protein number in the specific organ. The third ring (in light green) refers to unregulated proteins for each type of organ. The numbers in white represent the significantly dysregulated protein number in specific organ type (B-H adjusted p value < 0.05; fold change (FC): comparing COVID-19 with non-COVID-19 patients; |log_2_(FC)| > log_2_(1.2)). The innermost ring refers to the significantly dysregulated proteins for each organ (pink: upregulated; dark green: downregulated). The length of the radius represents the protein number. **B**. Proteins expression of CRP and CD163 across six organs (except testis). The y-axis stands for the protein expression ratio by TMT-based quantitative proteomics. Pair-wise comparison of each protein between COVID-19 patients and non-COVID-19 control groups was performed with student’s *t* test. Note: The cutoff of dysregulated proteins has been set at B-H adjusted p value < 0.05 and |log_2_(FC)| > log_2_(1.2). *, p < 0.05; **, p < 0.01; ***, p < 0.001.

Altogether we quantified 11,394 proteins from the above samples with a false discovery rate (FDR) less than 1% at both peptide and protein levels (Table S3, Figure 1A). The number of identified proteins ranged from 5828 (heart) to 9544 (kidney) across seven types of organs. We included 37 technical replicates of randomly selected tissue samples, as well as 18 pooled controls for each TMT batch (Figure S3A, Table S2). Proteins quantified in these technical replicates and control samples showed relatively low median coefficient of variance (CV) of 6.88% and 2.47%, respectively (Figure S3B, C). A total of 5336 dysregulated proteins were characterized from the seven types of organs between COVID-19 and control groups (Benjamini & Hochberg (B-H) adjusted p value < 0.05 and |log_2_(fold change)| > log_2_(1.2), Table S4, Figure 1A). Splenic red pulp samples were excluded from downstream analysis since they did not show statistically significant proteomic regulations changes (Figure 1A). Hierarchical clustering of the differentially expressed proteins from each organ type (Figure S3D) showed that these proteins well separated COVID-19 samples and controls. The t-distributed stochastic neighbor embedding shows that dysregulated proteomes of each organ type clustered tightly apart from each other (Figure S3E), consolidating that these selected proteins well resolved different tissue types. Except testis, the other six tissue types shared only 27 dysregulated proteins, suggesting different organs responded via diverse pathways. Among them, the acute inflammatory protein, C-reactive protein (CRP), and the M2 type macrophage highly expressed marker (Etzerodt and Moestrup, 2013), scavenger receptor cysteine-rich type 1 protein M130 (CD163) were the most upregulated proteins (Figure 1B), probably reflecting both the hyperinflammatory and repairing state in patients died from COVID-19.

### Six clusters of proteins relevant to SARS-CoV-2 infection

We then focused on six clusters of proteins including viral receptors and proteases, transcription factors (TFs), cytokines (and their receptors), coagulation system, angiogenesis associated proteins, and fibrosis markers due to their relevance to SARS-CoV-2 infection (Figure 2A, B). After cellular entry mediated by receptors and proteases, SARS-CoV-2 hijacks the host translation machinery and induces host inflammatory response via TFs, leading to hyper-inflammatory state, which might be associated with the clinically observed blood hypercoagulability as measured by blood tests (Figure S2), fibrosis and microthrombosis as examined by pathologists (Figure S1), and enhanced angiogenesis as reported by morphologic and gene expression examination in the lung of COVID-19 (Ackermann et al., 2020).

**Figure 2.**
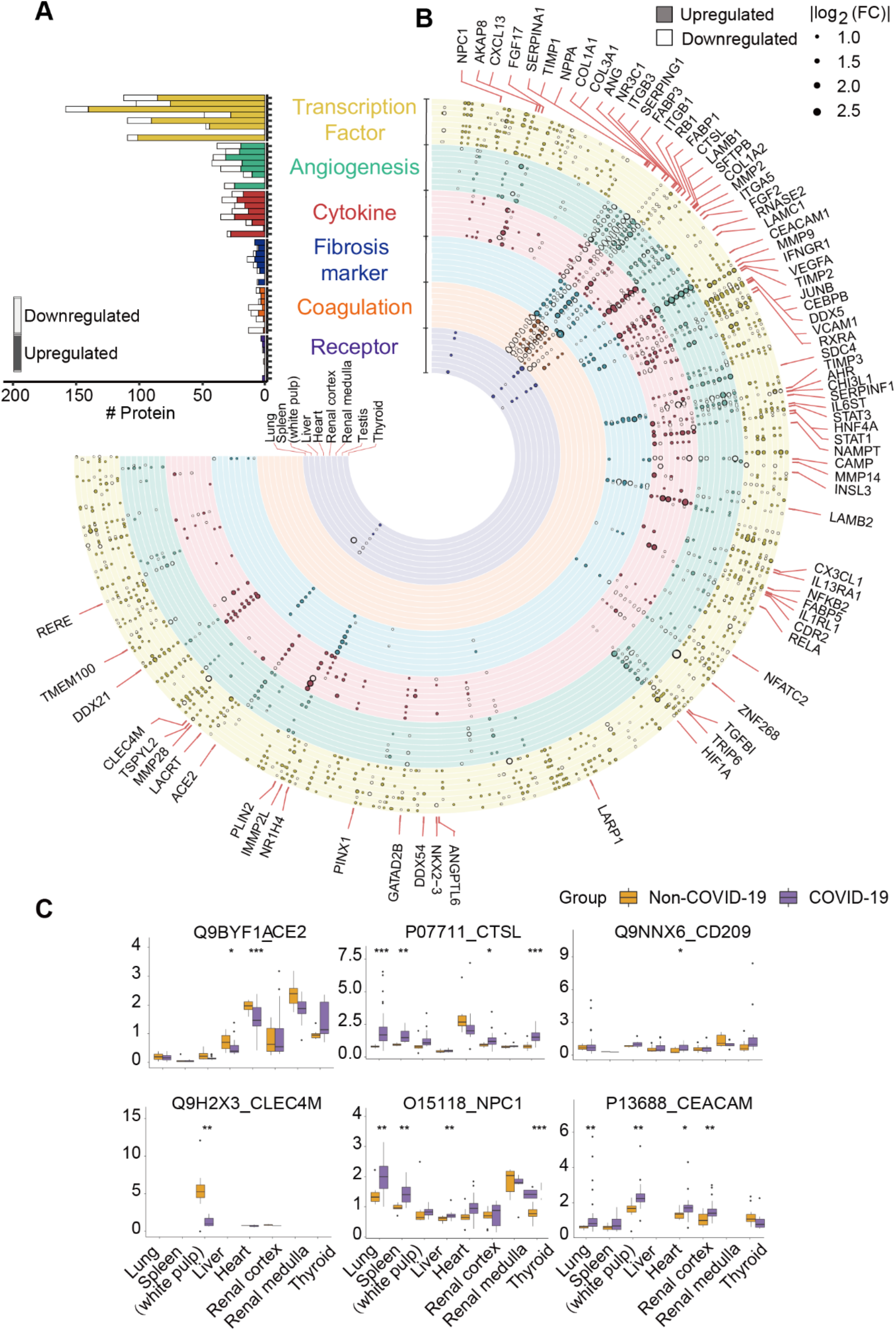
Six functional clusters of dysregulated proteins from seven organs between COVID-19 and non-COVID-19 patients. **A**. Counts of dysregulated proteins in six clusters of molecules, including potential virus receptors, fibrosis markers, cytokines (or its receptors), transcription factors (TF), coagulation system and angiogenesis associated proteins, were shown in a bar chart. Each column along y-axis represents a type of organ. The number of proteins is shown in x-axis. **B**. Landscape of 5336 significantly dysregulated proteins in seven organs. The dysregulated proteins in the six clusters are labeled as circles (solid: upregulated proteins; hollow: downregulated proteins). The size of circle indicates |log2(FC)|. Proteins discussed in Supplementary Discussion are labeled. **C**. Protein expression of potential virus receptors across multiple organs. The y-axis stands for the protein expression ratio by TMT-based quantitative proteomics. Pair-wise comparison of each protein between COVID-19 and non-COVID-19 patient groups was performed using student’s *t* test. Note: The cutoff of dysregulated proteins has been set at B-H adjusted p value < 0.05 and |log_2_(FC)| > log_2_(1.2). *, p < 0.05; **, p < 0.01; ***, p < 0.001.

Our data showed substantial regulation of TFs in COVID-19 autopsies. 395 out of 1117 quantified TFs were altered in at least one tissue type (Table S5, Figure S4A), and they were significantly enriched in spliceosome, viral carcinogenesis, among others as shown in Figure S4B. By matching the experiential fold-change with the predicted activation state in IPA, ten of these dysregulated TFs shown the same change trend, which participate in the hyperinflammation, hypoxia state and tissue injury in COVID-19 patients (Figure S4A, Supplementary Discussion).

Among the 242 quantified cytokines, 112 were significantly dysregulated and mapped to angiogenesis, response to growth factor and other pathways (Figure S4C, D, Table S5 and Supplementary Discussion). Nicotinamide phosphoribosyl transferase (NAMPT), glucocorticoid receptor (NR3C1) and interferon-gamma receptor 1 (IFNGR1) were dysregulated cytokines in most organs. NAMPT, which participates in multiple signaling pathways, such as IL6-STAT3 and NF-κB (Garten et al., 2015), was upregulated in the six organs except for testis. NR3C1, which is involved in the anti-inflammatory process(Baschant and Tuckermann, 2010), was downregulated in the five organs except for testis and thyroid. IFNGR1, which triggers host immune responses upon viral infection(Xia et al., 2018), was upregulated in the five organs except for testis and thyroid.

### CTSL, rather than ACE2, was upregulated in lungs

Our data identified six reported potential receptors or proteases for virus entry (Figure 2C, Supplementary Discussion), namely angiotensin-converting enzyme 2 (ACE2)(Hoffmann et al., 2020), C-type lectin domain family 4 member M (CLEC4M)(Jeffers et al., 2004) and member L (CD209)(Yang et al., 2004), Niemann-Pick C1(NPC1)(Cote et al., 2011), carcinoembryonic antigen-related cell adhesion molecule 1 (CEACAM1)(Tsai et al., 2003) and cathepsin L1 (CTSL)(Liu et al., 2020b). ACE2, the known receptor mediating SARS-CoV-2 entry, did not show significant regulation in lungs, but was downregulated in both kidney and heart in COVID-19 patients (Figure 2C). This observation is supported by the modulatory roles of ACE2 on angiotensin II, including inflammation, vasoconstriction and thrombosis(Liu et al., 2020a). Interestingly, our results showed CTSL, the serine protease of SARS-CoV-2 in the endosomal pathway, was significantly upregulated in the lung (Figure 2C), nominating it as a potential therapeutic target for COVID-19. In the SARS-CoV infection cell model, the inhibitor of CTSL has been proven to be effective for blocking the virus entry (Simmons et al., 2005).

### Multi-organ coagulation, angiogenesis and fibrosis

Our data suggested systematic dysregulation of coagulation, angiogenesis and fibrosis in COVID-19 patients, as shown in Figure 3 and Table S5. Microthrombi in the COVID-19 patients were observed in the lungs and kidneys of COVID-19 patients (Figure S1), in agreement with laboratory markers, such as D-dimer (Figure S2), indicating a prothrombotic state. Based on our analysis, the dysregulated proteins participating in coagulation, anticoagulation and fibrinolytic system could contribute to the coagulation disorders in COVID-19 (Figure 3A, Table S5, Supplementary Discussion). Besides, angiogenesis has also been reported in the lung (Ackermann et al., 2020). In our study, a total of 139 angiogenesis-related proteins from the nCounter PanCancer Progression Panel (NanoSring Technologies) were significantly dysregulated (Table S5, Figure 3B). Based on our data, multi-organ abnormal angiogenesis (not only in the lung) may have developed due to disordered coagulation, tissue hypoxia and excessive stimulation by cytokines in COVID-19 (Supplementary Discussion).

**Figure 3.**
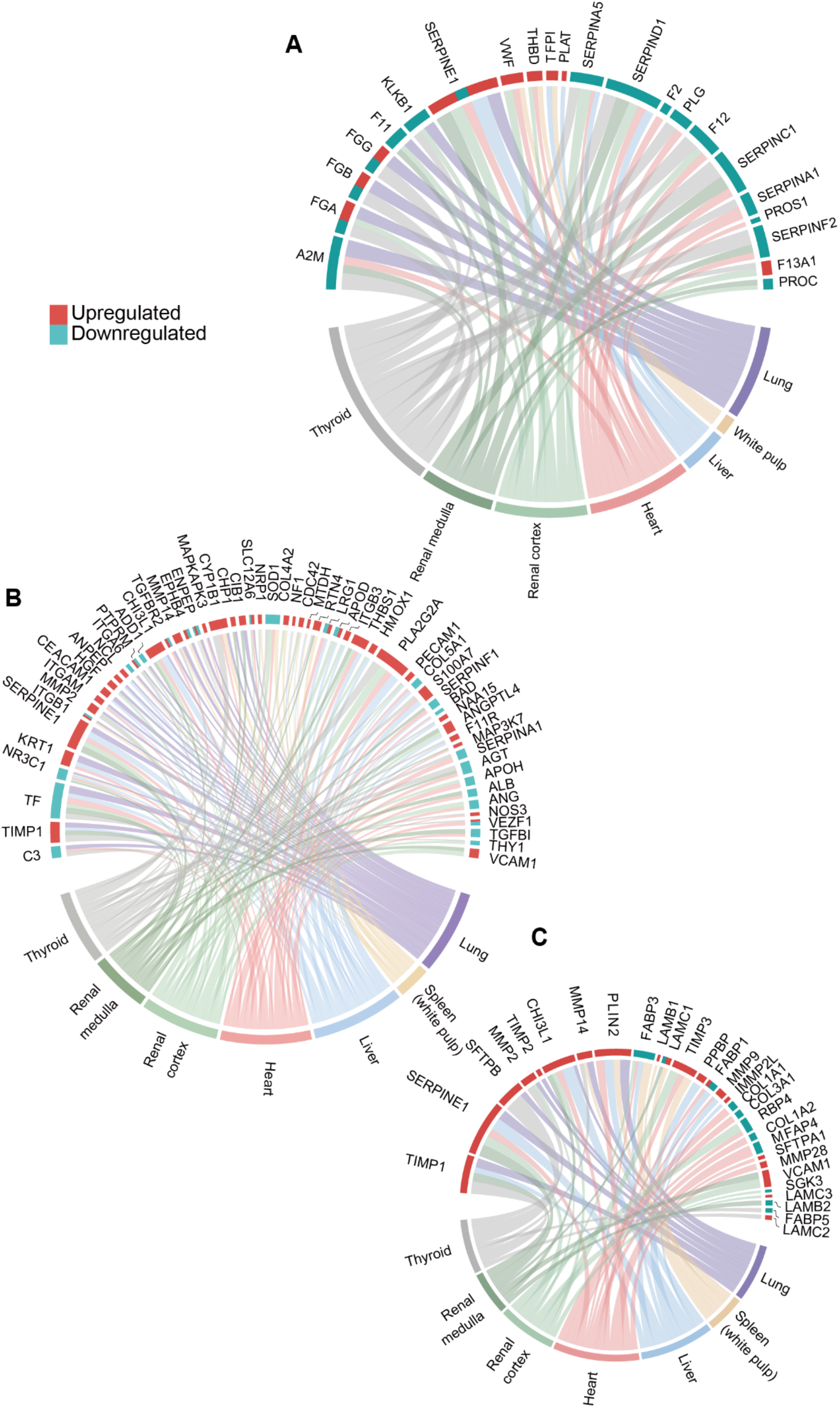
Coagulation, angiogenesis associated proteins and fibrosis markers regulated in multiple organs. Chord diagrams shows dysregulated and shared proteins in coagulation system (**A**), angiogenesis associated proteins (**B**), potential fibrosis markers (**C**) between COVID-19 and non-COVID-19 patients across multiple organs. The cutoff of dysregulated proteins has been set at B-H adjusted p value < 0.05 and |log_2_(FC)| > log_2_(1.2). The length of the brick for each protein corresponds to the sum of |log2(FC)| in multiple organs. The length of the brick for each organ corresponds to the sum of |log2(FC)| in one or more proteins.

As fibrosis has been observed in the lungs of COVID-19 patients (Figure S1, Table S1), we analyzed 29 fibrosis associated proteins in our dataset by matching with the reported potential fibrosis markers (Figure 3C, Table S5, Supplementary Discussion). Serpin family E member 1 (SERPINE1) and Chitinase 3 Like 1 (CHI3L1) were upregulated in the most organs of COVID-19 patients (Figure 3C). In the COVID-19 samples, we characterized 179 dysregulated proteins involved in the four stages of fibrosis, namely initiation, inflammation, proliferation and modification, according to nCounter Fibrosis Panel (NanoSring Technologies). These proteins formed 447 interactions according to String (Szklarczyk et al., 2019) and Cytoscape (Shannon et al., 2003) in the studied tissue types (Figure S5, Table S5, Supplementary Discussion). The dysregulated proteins associated with the fibrosis process are mostly in the lung, which is consistent with the pathological observation (Figure S1). Along with the fibrosis process, fewer proteins were dysregulated at the modification step compared with the other three stages, suggesting that chronic inflammation and fibrosis process may initiate at the molecular level without obvious pathological changes (Supplementary Discussion). The above observation may be instructive clinically to monitor COVID-19 patients after recovery for delayed development of tissue fibrosis.

### Dysregulated protein translation, glucose and fatty acid metabolism

To obtain a systematic understanding of biological processes represented by 5336 dysregulated proteins, we performed pathway enrichment in each tissue type using Ingenuity Pathway Analysis. Comparisons of the most enriched or dysregulated pathways (-log_10_ (p value) > 10 or ratio > 0.35 or absolute (Z-score) > 5) among seven organ types are shown in Figure 4A and Table S6. EIF2 signaling is involved in the regulation of mRNA translation (Roux and Topisirovic, 2018), which has been reported to be affected by virus infection (Bojkova et al., 2020). The lung, liver and thyroid shared a similar pattern of mRNA translation (Figure 4A), though there was little evidence of direct virus infection in the liver or thyroid. Most dysregulated proteins specific to the lung belonged to L13a-mediated translational silencing of ceruloplasmin expression (Figures 4B, 6A), which has been reported as an innate immune mechanism after the virus infection (Mazumder et al., 2014). We observed suppression of multiple metabolic processes including glycogenolysis, galactose degradation and glycolysis (Figure 4A, Table S6). In contrast, fatty acid β oxidation (FAO) and oxidative phosphorylation were activated in most organs, suggesting a switch to high-efficiency energy production mode to support virus replication in the lung and mRNA translation in the liver (Heaton and Randall, 2011). In addition, dysregulated FAO and oxidative phosphorylation also led to excessive generation of reactive oxygen species (ROS) and release of proapoptotic proteins, which induced liver necrosis (Figure S1). In the kidney, FAO was inhibited, which is thought to be a contributor to acute kidney injury (AKI) induced renal fibrogenesis (Kang et al., 2015). Indeed, AKI has been observed in most COVID-19 patients in our study (Table S1).

**Figure 4.**
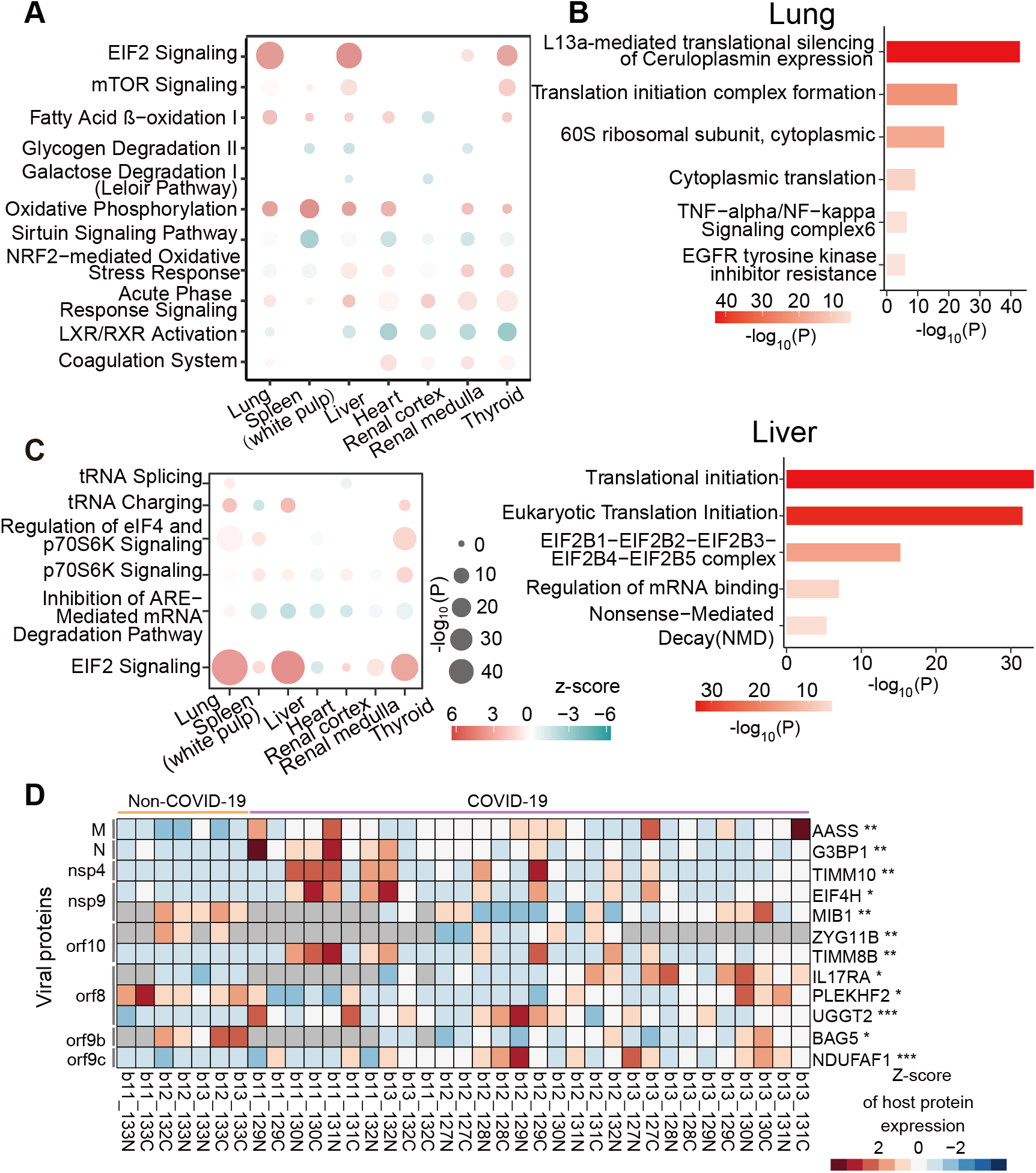
Dysregulated pathways in multiple organs. **A**. The top pathways dysregulated across multiple organs. Pathway analysis was performed using all dysregulated proteins in the specific organ using IPA. The top enriched pathways among seven organ types were selected by cutoff of either –log_10_(P) > 10, or ratio > 0.35, or absolute (Z-score) > 5. The size of circle represents the –log_10_(p value) and the color represent the Z score by IPA. **B**. Translation-associated pathway comparison across multiple organs. The size of circle represents the –log_10_(p) and the color represent the Z score by IPA. **C**. The pathways enriched by Metascape for translation initiation relating proteins that differentially expressed only in lung or liver respectively. **D**. Heatmap of SARS-CoV-2 interacting proteins dysregulated in the lung. The significance (‘Sig.’ as the short term in figures) was calculated using Student’s *t* test (p value: *, < 0.05, **, < 0.01; *** < 0.001). Note: The cutoff of dysregulated proteins has been set at B-H adjusted p value < 0.05 and |log_2_(FC)| > log_2_(1.2).

### Inter-organ crosstalk

We next investigated inter-organ crosstalk in COVID-19 patients based on our proteomics data (Figures 5, 6, Figures S7, Supplementary Discussion). The lung is the major target attacked by SARS-CoV-2. In our data, COVID-19 lung proteome showed unique enrichment of pathways which are known to be associated with virus infection, including mRNA decay and translation shutoff (Tanaka et al., 2012) (Figure 4C). Further, double-stranded RNA (dsRNA) and uncapped mRNA of the virus act as viral pathogen-associated molecular patterns (PAMPs) which trigger the innate immune response through recognition by pattern recognition receptors (PRRs) in the cytoplasm (Lin and Cao, 2020). By comparing with the SARS-CoV-2 protein interaction map (Gordon et al., 2020), 12 virus-host interacting proteins were dysregulated only in the lung, including stress granule-related factor G3BP1, mitochondrial protein TIM10, transcription regulator eIF4H, RING-type E3 ubiquitin ligase MIB1, pro-inflammatory cytokine receptor IL17RA, and member of Cullin RING E3 ligase 2 complex ZYG11B (Figure 4D). These proteins have been reported to promote virus replication, inhibit host mRNA expression, mediate the delivery of viral DNA through viral nuclear pore complex, participate in pulmonary fibrosis and degrade virus restriction factors (Gordon et al., 2020). G3BP1 was reported to interact with SARS-CoV-2 nucleocapsid (N) protein, which could be sequestered by viruses to promote their replication (Gordon et al., 2020). Mitochondrial proteins known to be targeted by viruses to enhance their replication (Williamson et al., 2012). The interaction between SARS-CoV-2 Nsp9 and eIF4H may indicate the inhibition of host mRNA expression (Gordon et al., 2020). The interaction between Nsp9 and MIB1 may mediate the delivery of viral DNA through nuclear pore complex (Gordon et al., 2020). IL17RA is associated with elevation of collagen and pulmonary fibrosis, and its inhibitor has been reported to reduce fibrosis in SARS infection (Mi et al., 2011). The interaction between Orf10 and ZYG11B may be hijacked for degradation of virus restriction factors or be blocked to protect itself from degradation (Gordon et al., 2020).

**Figure 5.**
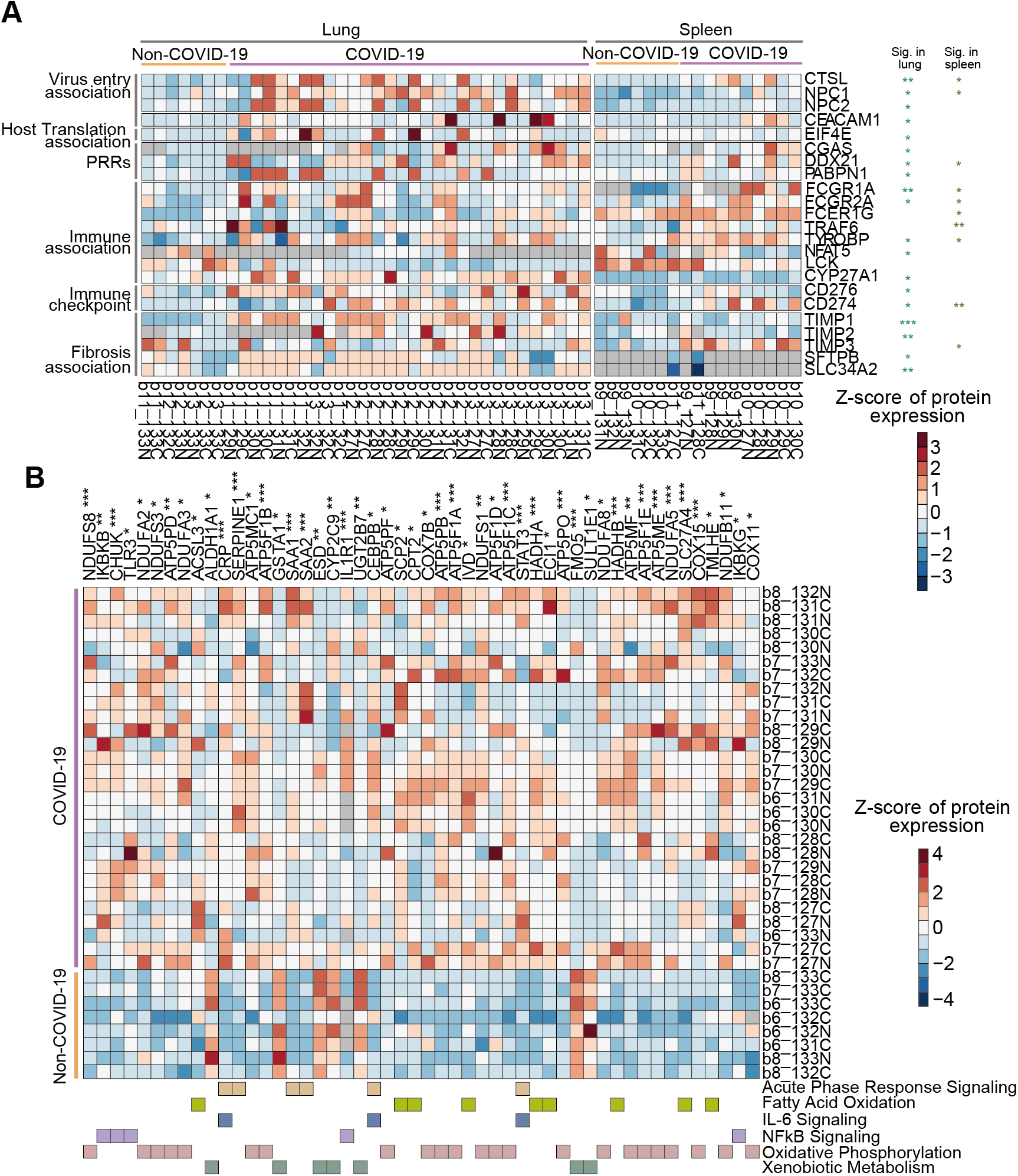
The heatmap of key dysregulated proteins in the lung, spleen and liver, respectively. **A**. The heatmap of key proteins in associated pathways in the lung and spleen model of Figure 6B. **B**. The heatmap of key proteins in associated pathways in the liver model of Figure 6B. Note: The significance (Sig.) of them in lung, spleen and liver was calculated using Student’s *t* test (p value: *, < 0.05, **, < 0.01; *** < 0.001).

**Figure 6.**
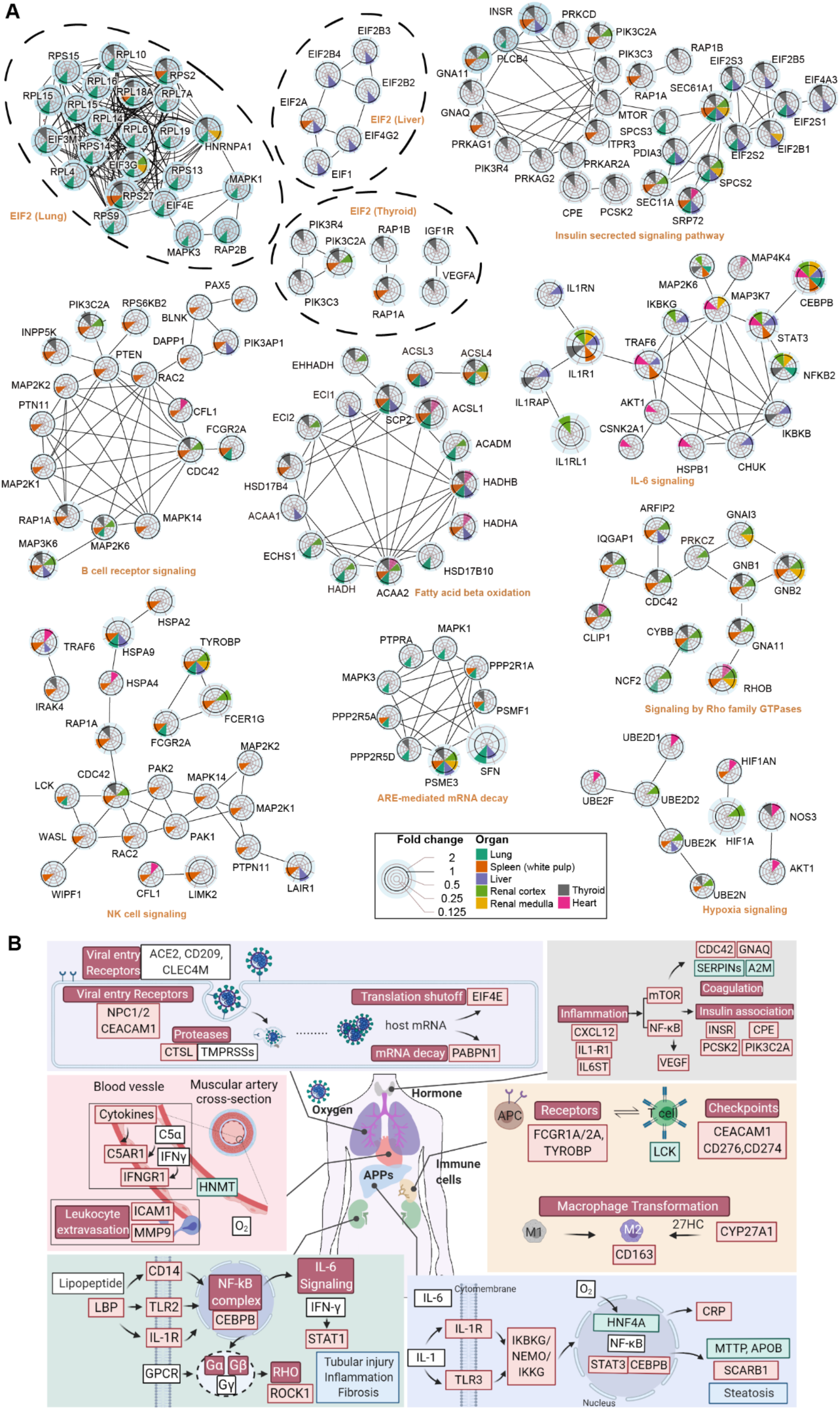
Dysregulated proteins and networks in six organs. **A**. Significantly enriched networks from the dysregulated proteins in the six organs. Each protein is depicted with radar chart for the six organs. Different organs are labeled with different colors. The shadow areas covering the circles indicate the FC values for each protein. **B**. A hypothetical systems view of the multiple organs’ responses to SARS-CoV-2 infection. In the lung, the virus and its released RNA could induce immune response and hijack the host translation mechanism. The innate and adaptive immune cells in the spleen and the cytokine induced acute phase proteins secreted by hepatic cells respond to antiviral defense. Such hyperinflammatory status across the whole body through circulatory system led to multi-organ injuries. More details are discussed in Supplementary information. Red box: upregulated proteins/pathways; green box: downregulated proteins/pathways; blue box, pathology of the organ.

We found that spleen and lung exhibited similar immune response patterns. T cell exhaustion and upregulation of monocytes biomarkers in the spleen (Figures 5A, 6B) suggested hyperinflammatory, which may have damaged the integrity of the gas exchange barrier and induced hypoxia. The hypoxia state would further stimulate the inflammatory responses (Eltzschig and Carmeliet, 2011). We further checked multiple markers for various immune cells in the spleen samples using immunohistochemistry (IHC) and found that the number of T and B lymphocytes were significantly reduced by IHC staining of CD3, CD4, CD8 and CD20, especially in the white pulp of COVID-19 patients, while the number of macrophages, M2 macrophages increased in the red pulp of COVID-19 patients by IHC staining of CD68 and CD163 respectively. These findings agreed with our proteomic data (Figure S6). Besides, the cytokines and immune cells in the blood cycling could induce acute phase response and NF-κB signaling in the liver (Figure 5B) (Alonzi et al., 2001; Israel, 2010), which sequentially leads to vascular hyperpermeability (Nagy et al., 2008) and vasogenic edema in the heart (Figures S1, S7A). In the thyroid and kidney, inflammatory infiltration and activated inflammation associated signals and proteins were also identified (Figures S7B, C, D).

### Testicular injuries

Compared with the other organ types, the number of differentially expressed proteins in testes was negligible. Only ten proteins were downregulated (Figure 7A). Insulin like factor 3 (INSL3), the most abundantly expressed proteins in Leydig cells (Uhlen et al., 2015), was the most dramatically decreased protein in COVID-19 testicular tissue (Figures 7A, D), suggesting impaired Leydig cell functions or a reduced Leydig cell population. Indeed, the histological examination revealed a reduction of Leydig cells (Figures 7B, C), in consistent with our previous pathological report (Yang et al., 2020). We also found five down-regulated proteins related to cholesterol biosynthesis (Figures 7A, D). All steroid hormones, including testosterone, are derived from cholesterol. Downregulation of dynein regulatory complex subunit 7 (DRC7), a sperm motility factor, suggested impaired sperm mobility (Morohoshi et al., 2020) (Figures 7A, D).

**Figure 7.**
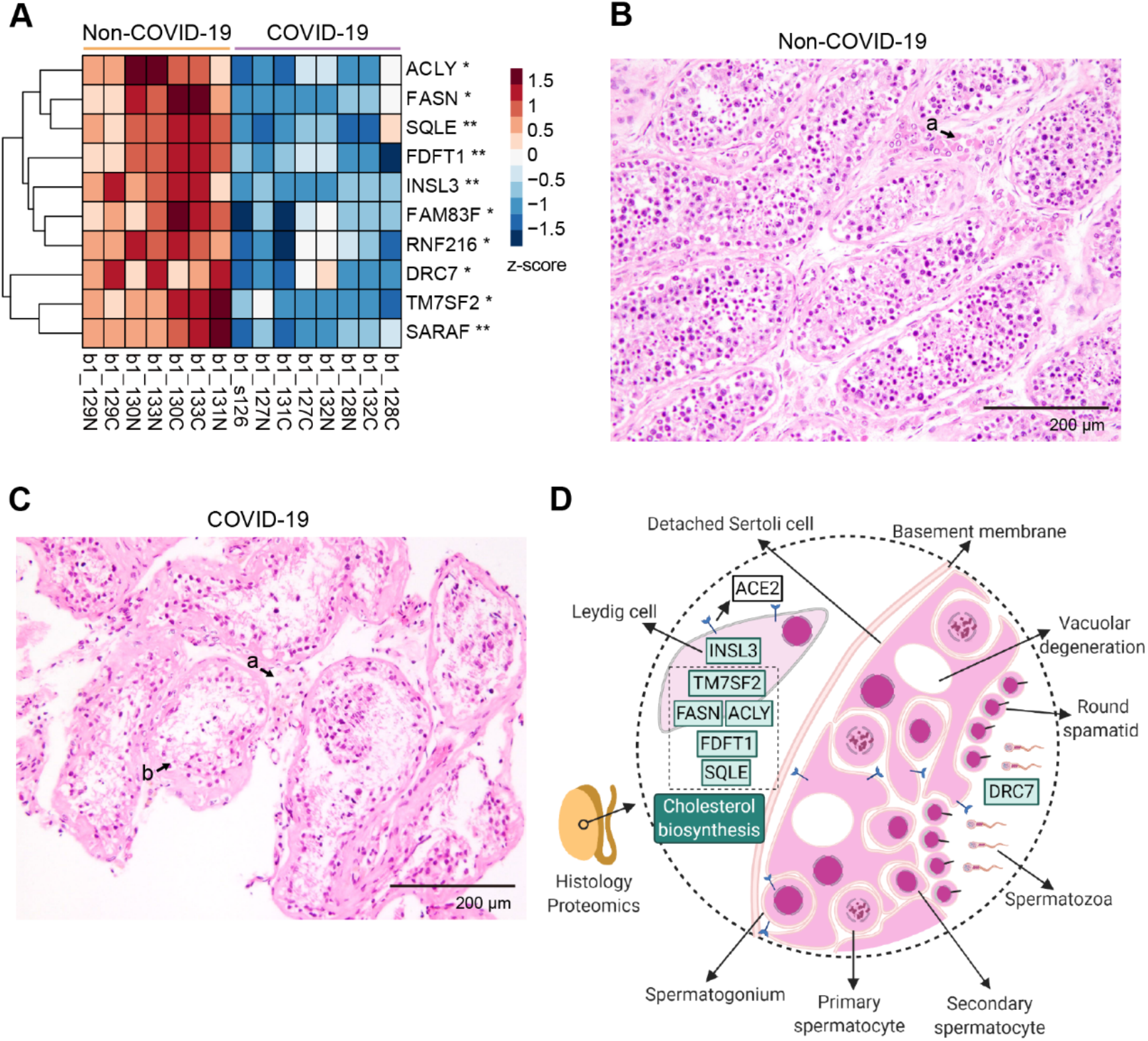
Proteomic and histopathological characterization of COVID-19 testes. **A**. The comparison of ten dysregulated proteins between COVID-19 and control samples. Pair-wise comparison of each protein between COVID-19 patients and control groups was performed with student’s *t* test. *, p < 0.05; **, p < 0.01. **B**. The H&E staining of testis from a control patient. H&E stained seminiferous tubules at high power (× 200) showed normal spermatogenesis. Clusters of Leydig cells are seen in the interstitium (a). **C**. The H&E staining of testis from a COVID-19 patient. In COVID-19 testes, H&E stained seminiferous tubules at high power (× 200) showed sparse intratubular cells with swollen and vacuolated Sertoli cells (b) and decreased number of Leydig cells in the interstitium (a). **D**. Diagram of the pathology in the COVID-19 testis, with seven dysregulated proteins indicated. The green box with black font inside shows the downregulated protein. The downregulation pathway is in green box with white font.

## CONCLUSION

In summary, we have quantified 11,394 proteins in seven types of tissue from patients died from COVID-19, and identified 5336 significantly dysregulated proteins compared to non-COVID-19 patients. This proteomic atlas uncovered multiple biological and pathological processes regulated in COVID-19, including immune response, protein translation, coagulation disorder, angiogenesis and profibrotic process. Crosstalk among multiple organs further linked the aforementioned processes by the hyperinflammatory environment with tissue hypoxia after SARS-CoV-2 infection. This systematic proteomic investigation provides a rich resource for improving our understanding of the molecular pathogenesis of SARS-CoV-2 infection, and offers clues for therapeutics. This study is limited by the sample size. In addition, future in-depth investigation of the perturbed pathways and the nominated therapeutics is needed.

## Data Availability

The proteomics data is deposited in ProteomeXchange Consortium (https://www.iprox.org/). Project ID: IPX0002393000. The data will be publicly available upon publication in a journal.

## ACKNOWLEDGMENTS

This work is supported by grants from Tencent Foundation (2020), National Natural Science Foundation of China (81972492, 21904107, 81672086, 81773022), Zhejiang Provincial Natural Science Foundation for Distinguished Young Scholars (LR19C050001), the Key Special Project of Ministry of Science and Technology, China (No.2020YFC0845700), the Fundamental Research Funds for the Central Universities (No.2020kfyXGYJ101), and Hangzhou Agriculture and Society Advancement Program (20190101A04). We thank Drs D.S. Li, O.L. Kon and the Guomics team for helpful comments to this study, and Westlake University Supercomputer Center and biomedical research core facilities for assistance in data generation and storage.

## AUTHOR CONTRIBUTIONS

T.G., X.N., Y.Z., Y.H. and J.X. designed and supervised the project. X.N., B.H and X.D summarized the pathological changes and SARS-COV-2 test. L.Q., R.S., Q.X., Q.Z., T.L., L.Y., W.G., W.L., Y.S., L.L., H.C., X.Y., M.L., S.L., X.D., C.Y., L.X., J.H., M.L., X.C., H.G., J.Y., Z.X. and G.R. conducted proteomic analysis. X.L., J.Z. and S.W. collected the autopsies. X.L, J.Z, J.F, X.C and D.Z organized the clinical data. H.M., H.S. and M.X. participated in clinical-pathological analysis. S.C, M.Y., L.P., G.X., X.X., J.W., H.P. performed H&E and IHC analysis. D.L., B.H. and Y.L. (Union Hospital) performed SARS-CoV-2 detection for lungs and analyzed the relevant data. L.Q., R.S., Q.X., T.L, L.Y, X.N., Y.Z., and T.G. interpreted the data with inputs from all co-authors. L.Q., R.S., Q.X., T.L, Y.Z. and T.G. wrote the manuscript with inputs from co-authors.

## DECLARTION OF INTERESTS

NA

## MATERIALS AND METHODS

### Clinical specimens and histological analysis

This study was approved by the Medical Ethics Committee of Union Hospital affiliated to Tongji Medical College of Huazhong University of Science and Technology (No.2020–0043–1) and Medical Ethical Committee of Westlake University (No.20200329GTN001).

All COVID-19 patients were from the Union Hospital affiliated to Tongji Medical College of Huazhong University of Science and Technology and met the diagnostic criteria by the National Health Commission of China. With the consent by the fatal COVID-19 patients’ families, ultrasound-guided percutaneous multi-point postmortem core biopsies were performed within two hours after death in a negative pressure isolation ward, from February to April 2020. Ultrasound examinations were performed using an ultrasound scanner (EPIQ 7C, Philips Medical Systems, Andover, MA, USA) equipped with an L12–5/S5–1 probe, or a China Mindray portable Ultrasound M9 GD equipped with an L10–3 probe. All autopsies were performed using the large core (14-guage) needle by the multitask team consisting of pathologists, thoracic surgeons, and ultrasound doctors. Procured tissues were preserved in the 10% neutral formalin immediately after the procedure and fixed for over 24 hours, and then routinely processed for paraffin blocks. Histological slices were prepared using a microtome and stained with hematoxylin and eosin (H&E) as described previously (Su et al., 2020b).Immunohistochemical stains (IHC) were performed for CD3 (Dako, Copenhagen, Denmark), CD4 (Dako), CD8 (Dako), CD20 (Roche, Tucson, AZ, USA), CD68 (Dako) and CD163(Cell Margue) according to the manufacturers’ protocols on a Dako Link 48 automated stainer (for CD3, CD4, CD8, CD68 and CD163) or a Roche Benchmark XT Ultra system (for CD20). All slides were examined by at least two senior pathologists independently. A total of 144 microscopy-guided dissection samples were analyzed from 73 specimens, including heart (9 specimens), lung (15 specimens), liver (10 specimens), kidney (10 specimens), spleen (9 specimens), testis (5 specimens), and thyroid (15 specimens), see Tables S1, S2. We also collected 74 control samples from archived blocks of 56 non-COVID-19 patients before December, 2019 (Tables S1, S2).

### Proteomics data acquisition

Around 1–1.5mg FFPE tissue samples were processed to generate peptide samples using accelerated pressure cycling technology (PCT) assisted sample preparation method as described previously(Gao et al., 2020; Zhu et al., 2019). Briefly, the samples were dewaxed by heptane and rehydrated by ethanol solution of different concentrations; then undergone acidic hydrolysis by 0.1% formic acid, and basic hydrolysis by 0.1 M Tris-HCl (pH 10.0); followed by tissue lysis by 6 M urea/2M thiourea (Sigma) buffer, reduction by Tris(2 carboxyethyl)phosphine (TCEP, Sigma) and alkylation by iodoacetamide (IAA, Sigma) in PCT. Then the lysates were digested using PCT by a mix of LysC and trypsin (Hualishi Tech. Ltd, Beijing, China) and the digestion was quenched by trifluoroacetic acid. After C18 cleaning, 7 μg of peptides from each of the 218 tissue samples, and 37 technical replicates, 34 pooling samples including common pooled samples and tissue specific pooled samples (Table S2) were taken for the following TMTpro 16plex labeling based proteomics analysis. The common pooled samples were prepared by mixing equal amount of peptide from 6 different human organs including testis, lung, kidney, spleen, heart and liver. The tissue specific pooled samples were prepared by mixing equal amount of peptides from all samples of a specific organ) were labeled with TMTpro 16plex reagent (Thermo Fisher Scientific™, San Jose, USA)(Li et al., 2020). After quenching the labeling reaction by hydroxylamine, 16 TMT-labeled peptide samples were combined and cleaned by C18 columns (Waters, Sep-Pak Vac tC18 1cc, 50mg). Fractionation was performed on Thermo Ultimate Dinex 3000 (Thermo Fisher Scientific™, San Jose, USA) with an XBridge Peptide BEH C18 column (300Å, 5 μm × 4.6 mm × 250 mm) (Waters, Milford, MA, USA). The sample was separated using a 60 min LC gradient from 5% to 35% acetonitrile (ACN) in 10 mM ammonia (pH = 10.0) at a flow rate of 1 mL/min to 60 fractions. The fractions were combined using the following strategy: 1) combine the 1^st^ to 12^th^, 59^th^ and 60^th^ fraction; 2) combine the 13^rd^, 20^th^ and 52^nd^ fraction; 3) combine the 14^th^, 19^th^ 54^th^ and 55^th^ fraction; 4) combine the 15^th^, 18^th^ and 58^th^ fraction; 5) combine the 16^th^, 22^nd^ and 53^rd^ fraction; 6) combine the 17^th^, 21^st^, 56^th^ and 57^th^ fraction; 7) combine the 23^rd^ and 50^th^ fraction; 8) combine the 24^th^ and 48^th^ fraction; 9) combine the 25^th^ and 47^th^ fraction; 10) combine the 26^th^ and 49^th^ fraction; 11) combine the 27^th^ and 51^st^ fraction; 12) no combination for 28^th^ to 46^th^ fraction. Together, we got 30 combinations of fractionates and dried them in vacuum. Dried peptides were re-dissolved in 2% ACN/0.1% formic acid and then analyzed with QE HFX or QE HF with the same LC-MS/MS settings as described in Shen et al (Shen et al., 2020b). The MS raw data were analyzed by Proteome Discoverer (Version 2.4.1.15, Thermo Fisher Scientific) using a combination of the FASTA files of 20,365 reviewed Homo sapiens proteins downloaded from UniProt website on 14 April, 2020 and the SARS-CoV-2 virus FASTA of 10 SARS-CoV-2 proteins downloaded from NCBI (version MN908947.3). Precursor ion mass tolerance was set to 10 ppm, and product ion mass tolerance was set to 0.02 Da. Other parameters in Proteome Discoverer analysis are identical to our previous study(Shen et al., 2020b). The grouped abundance ratio of 15 tissue samples to the pooled sample in the same batch was selected as the value of proteins in the protein matrix for the statistical analysis. As for the lung, spleen, liver, heart, kidney and testis samples, the pooled sample for ratio calculation was chosen as the common pooled sample in the corresponding batch. As for the thyroid samples, the pooled sample for ratio calculation was chosen as the tissue specific pooled sample in the corresponding batch.

### Quality control of proteomics data

The quality control (QC) of proteomics data was similar as described in Shen et al(Shen et al., 2020b). We performed QC in the following levels: i) As to the batch design, one plex of TMTpro 16plex was set as a pooled sample for batch alignment and quantitative accuracy (as calculated in the grouped abundance ratio); ii) During the MS acquisition, we analyzed mouse liver protein digests for MS instrument performance evaluation every two batches, and analyzed MS buffer A (2% ACN/0.1% formic acid) as blanks every six LC-MS/MS injections; iii) We random distributed the peptide samples from every organ of COVID-19 patients and non-COVID-19 patients into each batch; iv) For data analysis, we calculated the median coefficient of variation (CV) of the proteomics data for reproducibility. As for the pooled controls, we calculated the CV by the log_2_(abundance) of quantified proteins in the pooled controls of each organ (Figure S3B). The pooled samples for CV calculation was the same as the ones for grouped ratio calculation. As for the technical replicates, we calculated the CV by expression ratio of quantified proteins in the technical replicates of each sample (Figure S3C). Besides, unsupervised clustering for the proteomics data were performed, including heatmapand tSNE (Figures S3D, E). After the selection of differentially expressed proteins (adjusted p value by B-H correction < 0.05 and |log_2_(FC)| > log_2_(1.2)), the heatmap of each organ by the dysregulated proteins was performed and the few samples that couldn’t be grouped correctly were excluded for further statistical analysis, probably due to unknown pre-analytical reasons.

### Statistical analysis

Log_2_ fold-change (FC) was calculated by the mean of proteins expression ratio in a specific tissue between COVID-19 and non-COVID-19 patient groups. Student’s *t* test was performed for each pair of comparing groups. Adjusted p values were calculated using B-H correction. The criteria for significantly dysregulated proteins selection was that the adjusted p value should be less than 0.05 and |log_2_FC| should be larger than log_2_(1.2). Statistical analysis was performed using R (version 3.6.3).

### Pathway/network analysis

The pathway enrichment was analyzed by either ingenuine pathway analysis (IPA)(Kramer et al., 2014) (with p value < 0.05, Z score > 0 or < 0), or Metascape web-based platform(Zhou et al., 2019) (with p-value < 0.05), or string web-based platform(Szklarczyk et al., 2019). The immunological proteins were mapped against GSEA-immunologic gene sets in Metascape platform with our differentially expressed proteins and then the enriched pathways were distinguished by IPA analysis.

## SUPPLEMENTARY FIGURE LEGENDS

**Figure S1.**
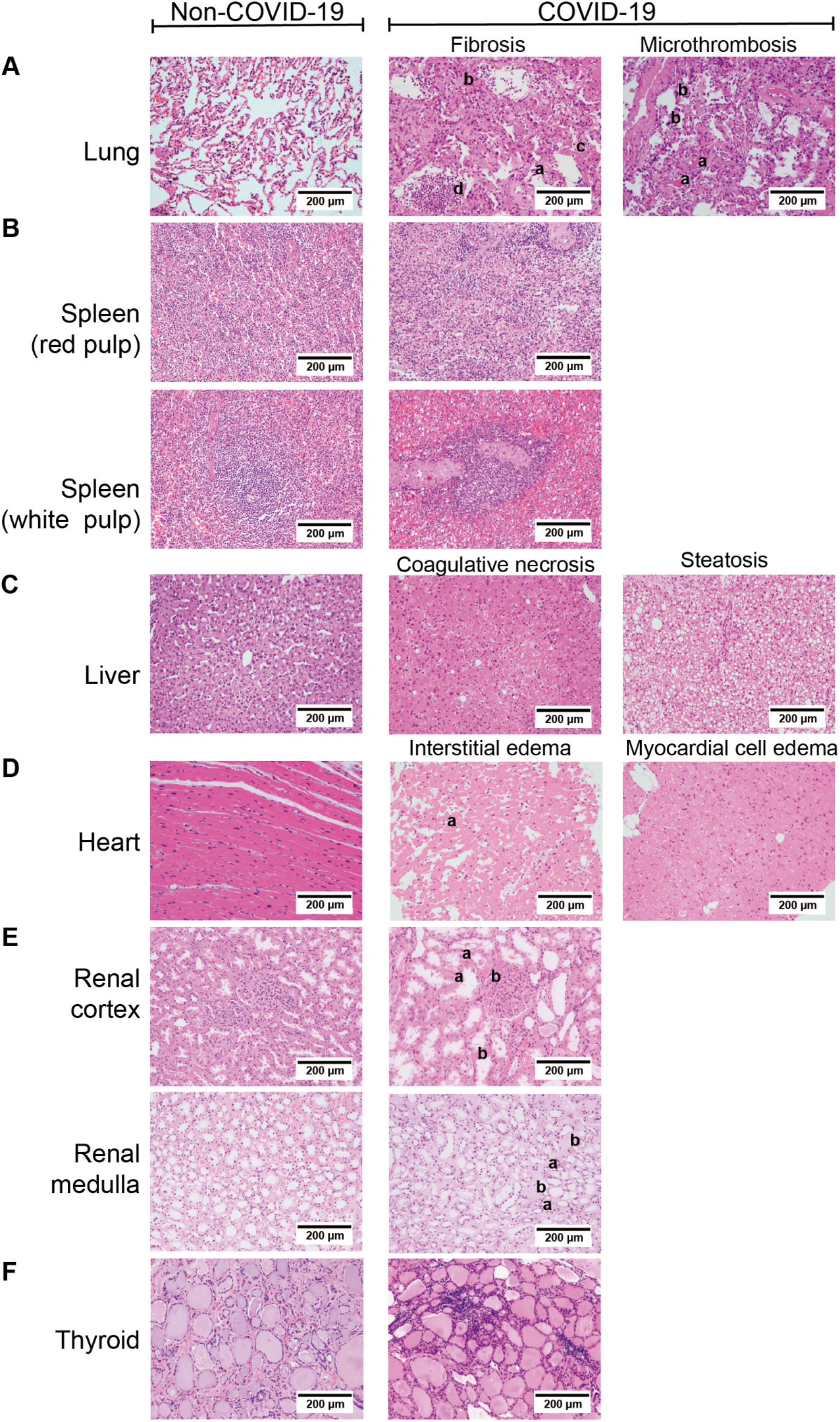
Comparison of histopathological features of organs between COVID-19 and non-COVID-19 patients. (H&E ×200). The left column shows the results of non-COVID-19 patients and the right part shows the results of COVID-19 patients. **A.** Pathological features of lungs from COVID-19 patients. (i) The lung showed diffuse alveolar damage, with the alveolar epithelia being replaced by hyperplastic type II alveolar epithelia and falling of type II alveolar epithelia (a). Alveolar septa were thickened with the proliferation of fibroblasts/myofibroblasts and fibrosis (b). Fibrinous exudation and hyaline membrane formation were found (c), with neutrophils aggregation in the alveolar cavity (d). (ii) There were some macrothrombusus in small vessels of the lung (a), and megakaryocytes in the alveolar septal capillaries (b). **B**. Pathological findings of the spleen from COVID-19 patients. (i) The red pulp of the spleen expanded, and splenic sinus extended with hyperemia accompanying with macrophage proliferation. (ii) The white pulp of spleen was appeared atrophy with lymphocytes significantly reduced. **C**. Pathological findings of the liver from COVID-19 patients. (i) Coagulative necrosis of hepatocytes was observed in zone III. (ii) The hepatocytes exhibited prominent steatosis. **D**. Pathological findings of the heart from COVID-19 patients. (i) Atrophic myocardia and scant lymphocytes (a) were present in the edematous cardiac interstitium. (ii) The myocardium showed hydropic degenerative change. **E**. Pathological findings of the kidney from COVID-19 patients. (i) In the renal cortex, the proximal tubules showed prominent acute tubule injury manifested as the loss of brush border and epithelial cells necrosis in the tubular lumens (a). Microthrombi in peritubular capillaries and glomeruli were frequently seen (b). (ii) In the renal medulla, the collecting ducts showed occasional cellular swelling and atrophy (a), and edema without significant inflammation (b). **F**. Pathological findings of the thyroid from COVID-19 patients. Lymphoid infiltration was found in some of interfollicular region. Neither neutrophilic infiltration nor necrosis was present.

**Figure S2.**
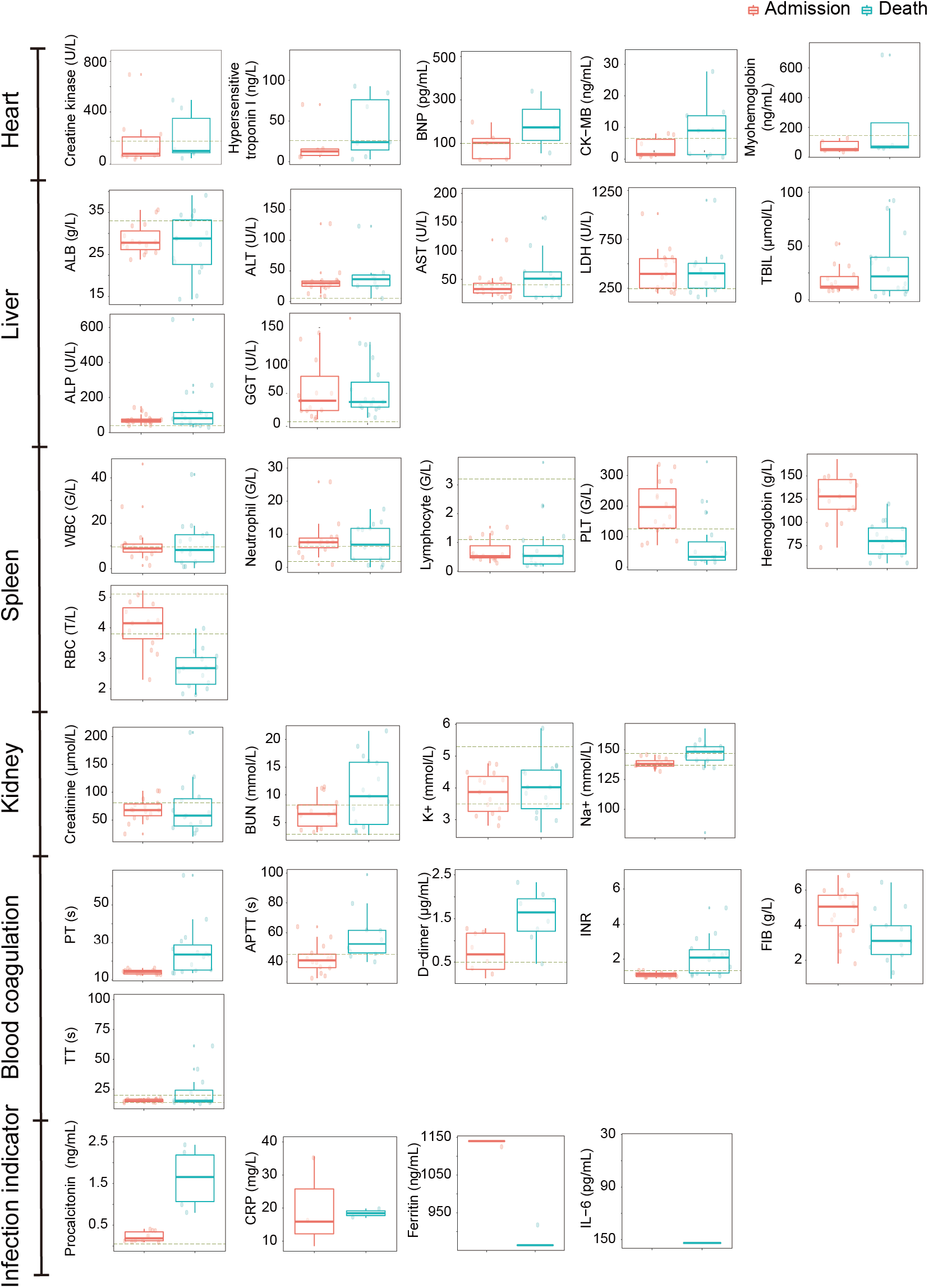
Laboratory characteristics of multi-organ dysfunction and infection indicators with COVID-19. Boxplots display the blood biochemistry tests based on samples collected from COVID-19 patients on the day of admission (red) and death (green). The dash line represents the normal range of laboratory characteristic. BNP, brain natriuretic peptide; CK-MB, creatine kinase-MB; ALB, albumin; ALT, alanine aminotransferase; AST, aspartate aminotransferase; LDH, lactate dehydrogenase; TBIL, total bilirubin; ALP, alkaline phosphatase; GGT, gamma-glutamyl transferase; WBC, white blood cells; PLT, platelets; RBC, red blood cells; BUN, blood urea nitrogen; PT, prothrombin time; APTT, activated partial thromboplastin time; INR, international normalized ratio; FIB, fibrinogen; TT, thrombin time; CRP, C-reactive protein; IL-6, interleukin-6.

**Figure S3.**
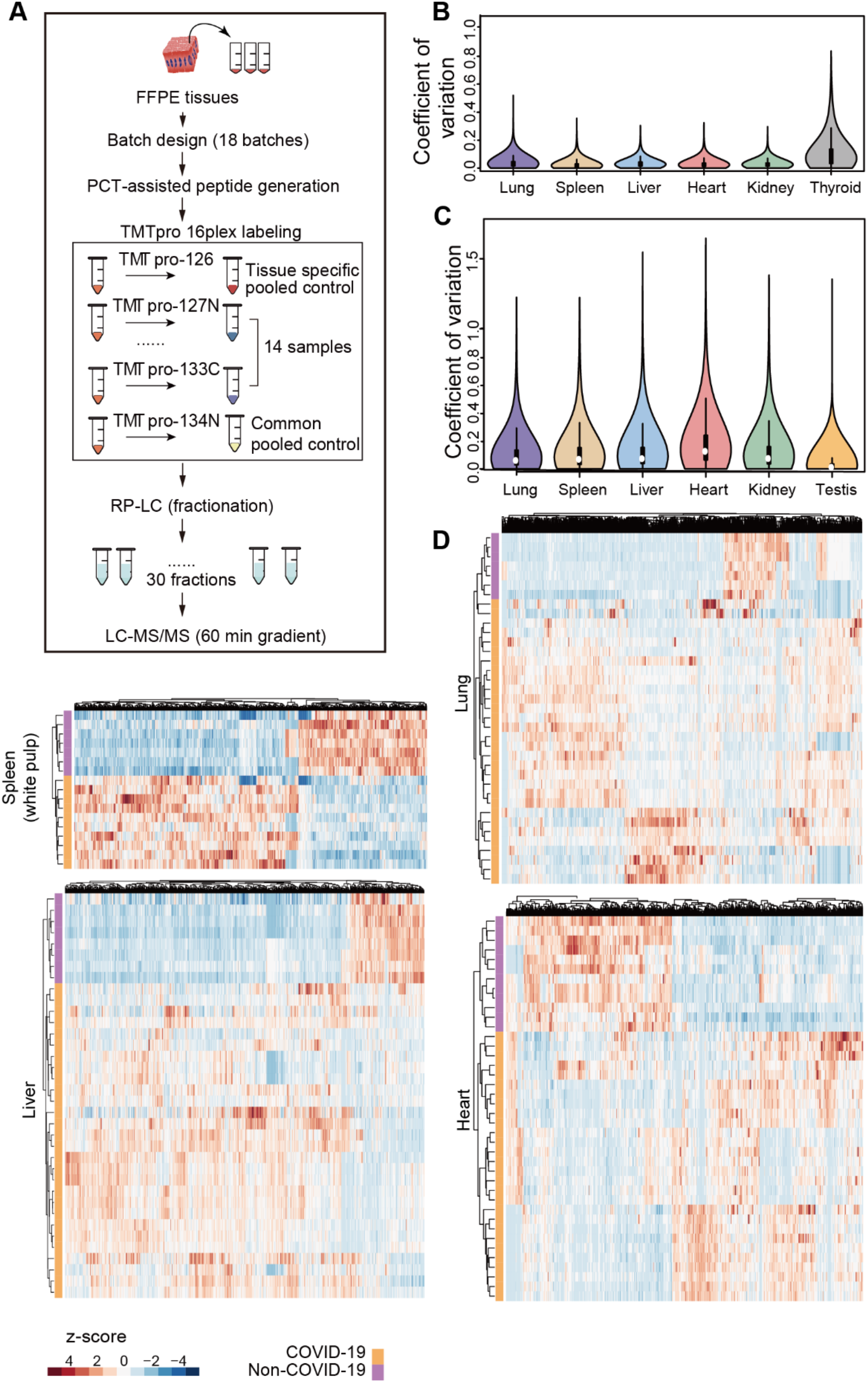
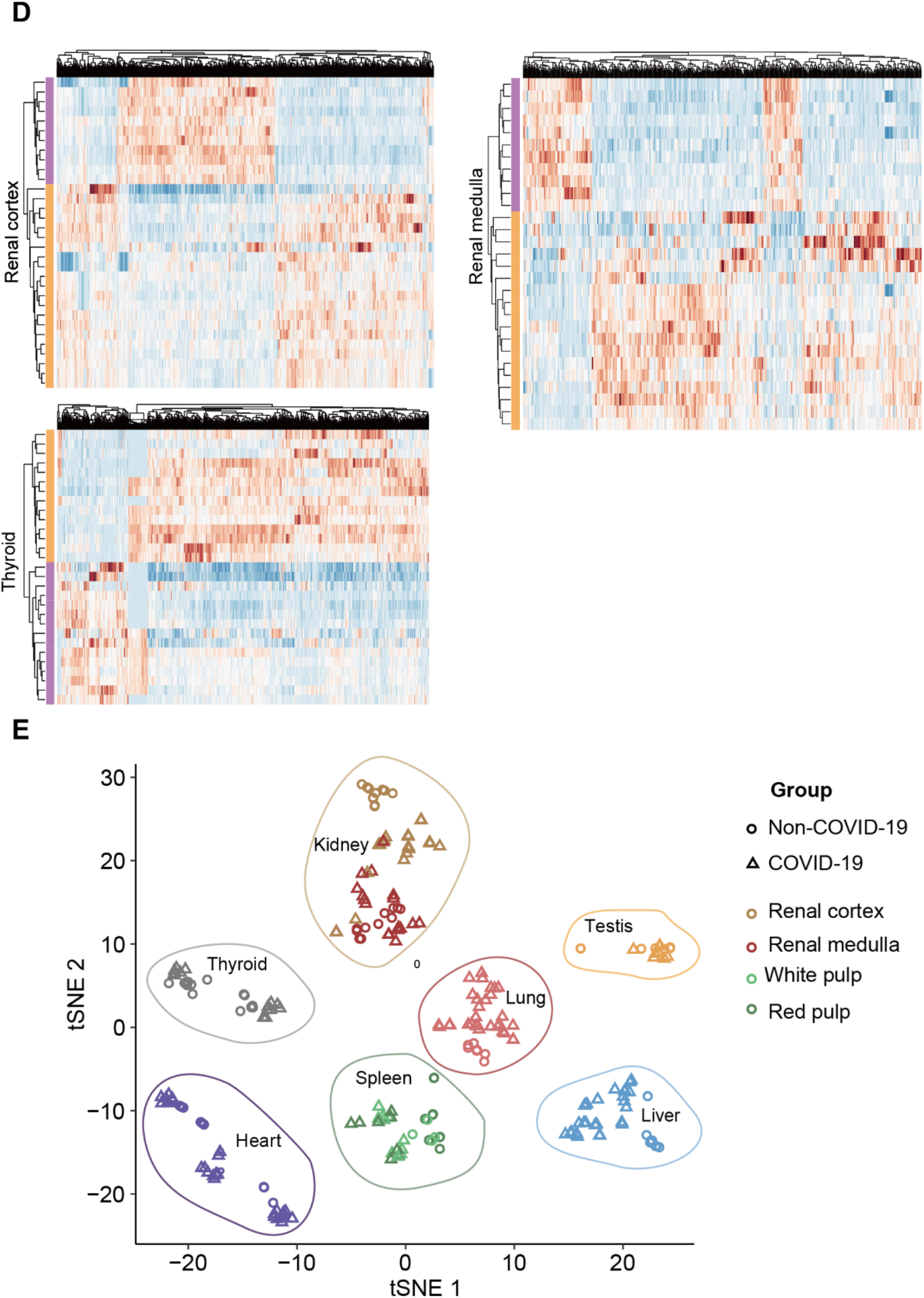
Proteomic workflow and quality control of proteome data analysis. **A**. The workflow of TMT-labeling based quantitative proteomics analysis employed in our study. 288 peptide samples containing 37 technique replicates, 16 common pooled controls and 17 tissue specific pooled controls were distributed into 18 batches and analyzed by TMT 16-plex labeling based proteomic strategy. FFPE, formalin-fixed and paraffin-embedded; PCT, pressure cycling technology; RP-LC, reversed phase liquid chromatography; LC-MS, liquid chromatography-mass spectrometry. **B**. The median CV of the proteomics data is calculated by the log_2_(abundance) of quantified proteins in the pooled controls of each organ. **C**. The median CV of the proteomics data is calculated by expression ratio of quantified proteins in the technique replicates of each organ. **D**. Heatmap of organ-specific dysregulated proteins for lung (37 samples * 1606 proteins), splenic white pulp (17 samples * 1726 proteins), liver (36 samples * 1969 proteins), heart (40 samples * 919 proteins), renal cortex (32 samples * 1585 proteins), renal medulla (29 samples * 642 proteins) and thyroid (29 samples * 1297 proteins) between COVID-19 patients and control groups were shown. **E**. The t-distributed stochastic neighbor embedding (tSNE) visualization of significantly dysregulated proteins in multi-organ proteomes of COVID-19 (triangle) and non-COVID-19 (circle) patients. The color of triangle or circle represents the organ type. Note: The cutoff of dysregulated proteins has been set at B-H adjusted p value < 0.05 and |log_2_(FC)| > log_2_(1.2).

**Figure S4.**
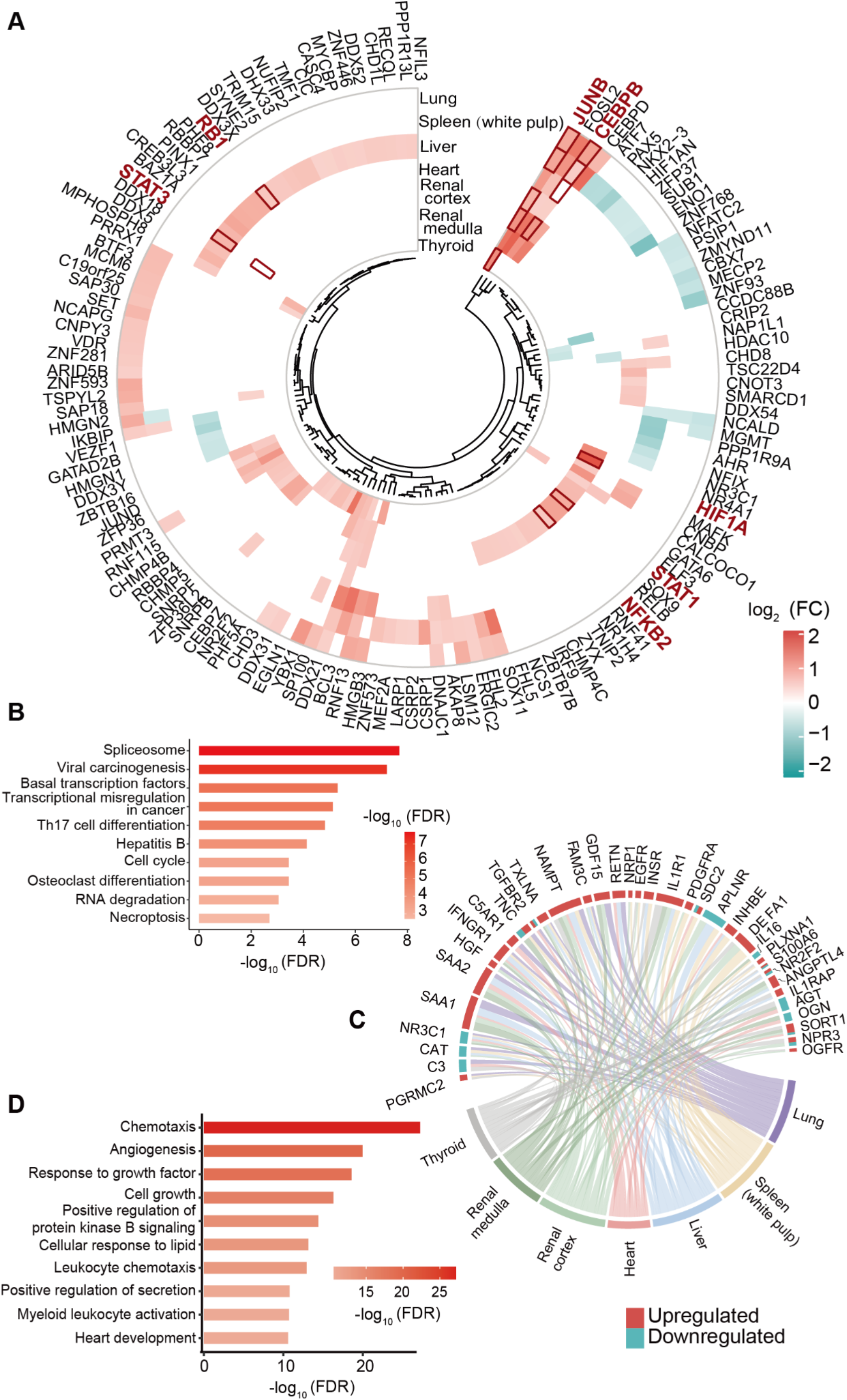
Transcription factors and cytokines dysregulated in multiple organs. **A.** Circular heatmap of dysregulated transcription factors. The selected transcription factors with FC larger than 1.5 from dysregulated proteins are shown. Some of the transcription factors in specific tissue with the same trend of changes, and distinguished out by upstream analysis are highlighted in red frame with red and bold font. **B**. Pathway enrichment analysis of dysregulated transcription factors between COVID-19 patients and control groups. Reactome pathways were enriched by String. FC: fold change; FDR: false discovery rate. **C**. The chord diagram shows the shared cytokines (or their receptors) dysregulated between COVID-19 and non-COVID-19 patient groups across multiple organs. **D**. Pathway enrichment analysis of the dysregulated cytokines. Gene ontology (GO) pathways were enriched by Metascape. The cutoff of dysregulated proteins has been set at B-H adjusted p value < 0.05 and |log_2_(FC)| > log_2_(1.2). The color of protein represents the upregulation or downregulation pattern. The length of the brick for each protein corresponds to the sum of |log2(FC)| in multiple organs. The length of the brick for each organ corresponds to the sum of |log2(FC)| in one or more proteins.

**Figure S5.**
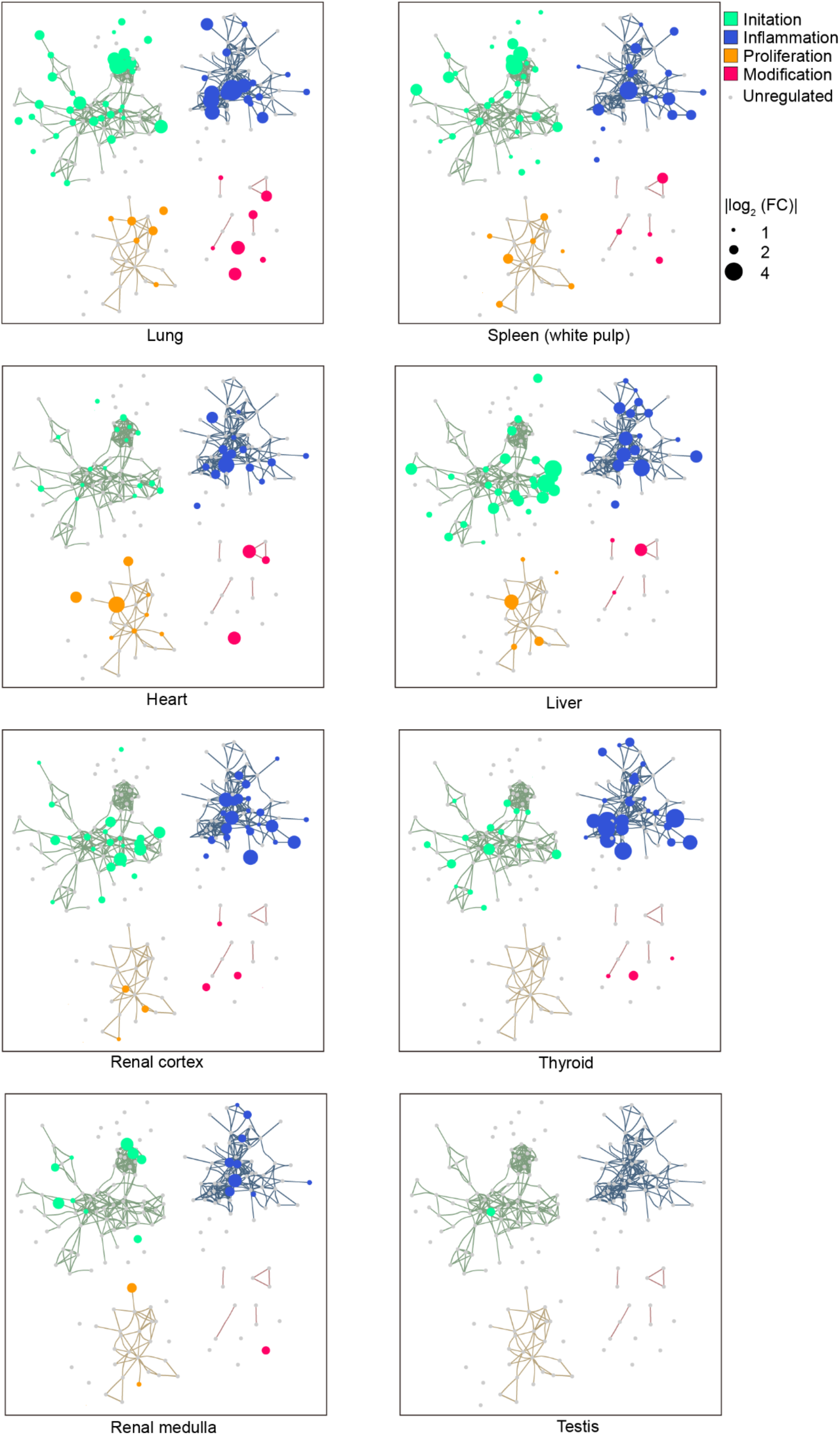
The network of fibrosis associated proteins over four fibrosis stages in multiple organs. The network shows the interactions among 179 stage-specific dysregulated proteins between COVID-19 and non-COVID-19 patient groups in each organ over the four fibrosis stages: initiation (green), inflammation (blue), proliferation (orange), and modification (red). The network analysis was performed using String and Cytoscape. The size of the dot represents the value of |log2(FC)|. Note: The cutoff of dysregulated proteins has been set at B-H adjusted p value < 0.05 and |log_2_(FC)| > log_2_(1.2).

**Figure S6.**
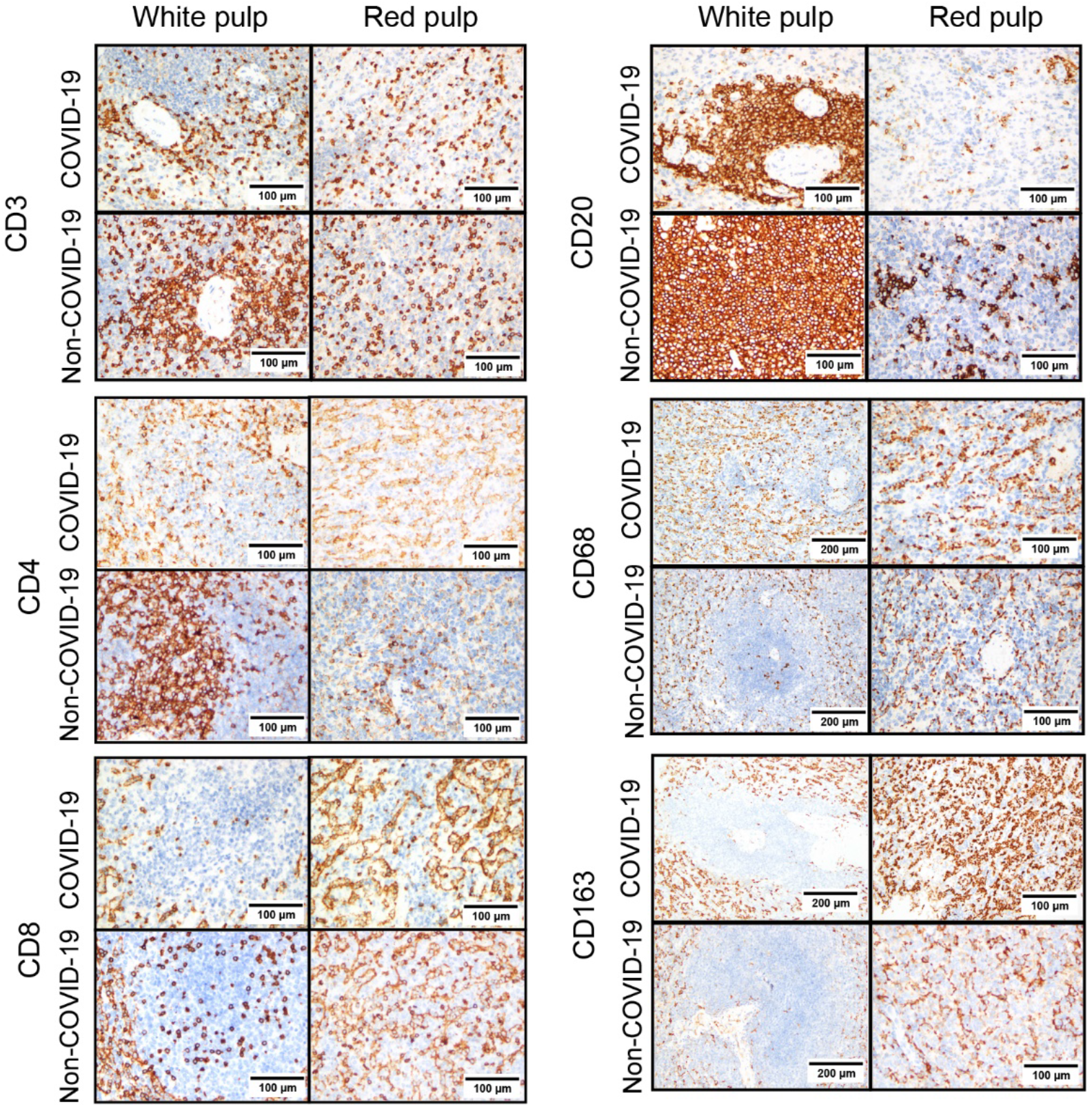
Representative images of immunohistochemical (IHC) staining for immune cells in spleens of COVID-19 and non-COVID-19 patients. White pulp and red pulp samples were from patient P1 (COVID-19) and patient C57 (non-COVID-19), respectively. The multiple markers for various immune cells in the spleen samples using IHC including CD3 for total T cell, CD4 for CD4 positive T cell, CD8 for CD8 positive T cell, CD20 for B cell, CD68 for macrophage and CD163 for M2 macrophage were shown.

**Figure S7.**
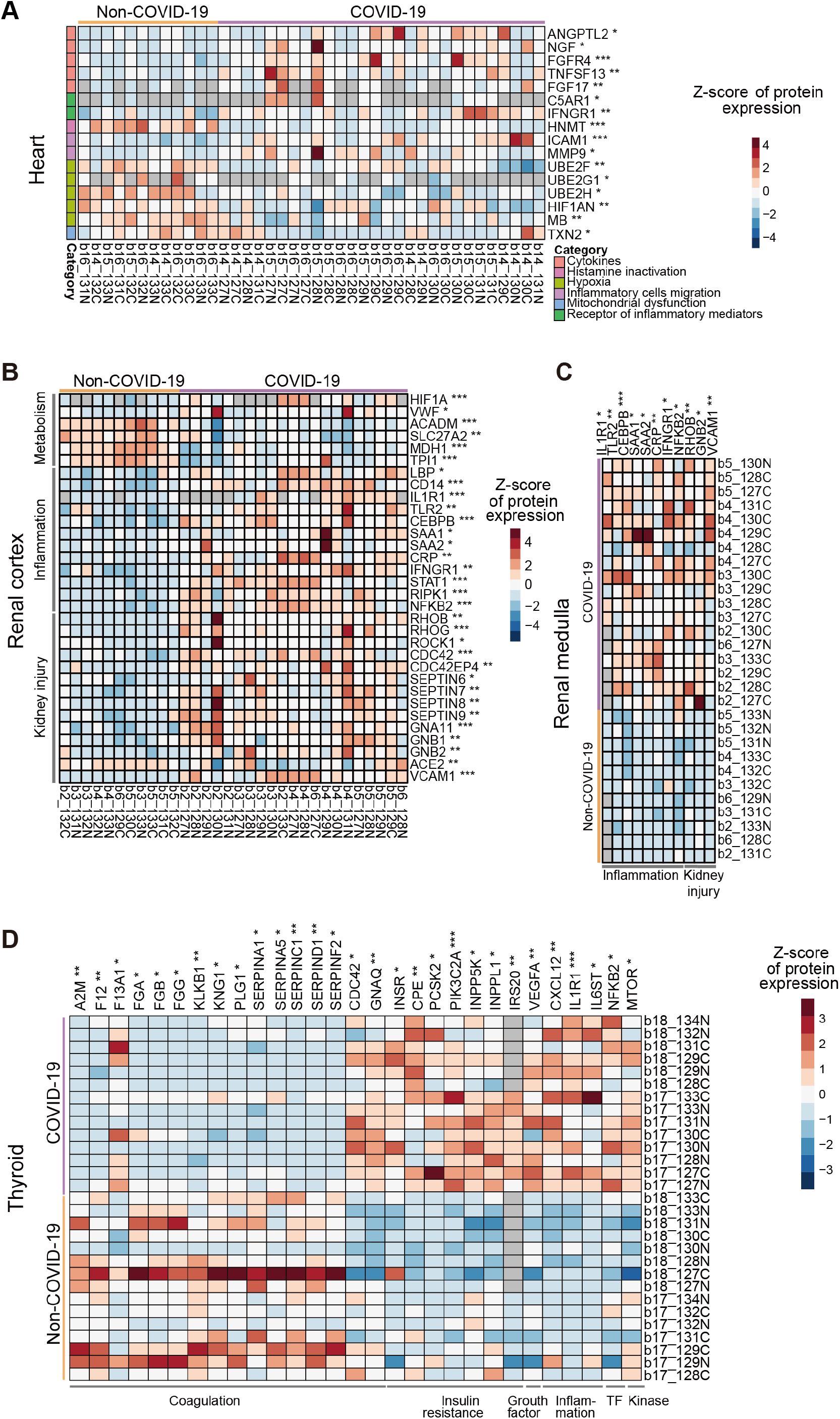
The key dysregulated proteins in the heart, renal cortex, renal medulla and thyroid associated with the enriched pathways by IPA. The heatmap of key proteins in associated pathways in the heart (**A**), renal cortex (**B**), renal medulla (**C**), and thyroid (**D**). The significance (Sig.) of them in each type of organ was calculated using Student’s t test (p value: *, < 0.05, **, < 0.01; *** < 0.001).

## Overview

In this supplementary file, we discussed in greater details the six clusters in Figure 2 and proteomic changes in the seven organs as briefly discussed in Figure 6 and 7.

## Six clusters (supple for Figures 2 and 3)

### Cluster 1: Receptors

Known receptors for SARS-CoV-2

To investigate the virus induced pathologic features at the protein level, we first focused on the lung proteome. As the entry of the virus, the known ACE2 receptor and cathepsin L1 (CTSL) for SARS-CoV-2^1^, known C-type lectin domain family 4 member L(CD209) and member M (CLEC4M) receptors for SARS^2,3^ were identified in the lung proteome in this study. Interestingly, ACE2, CD209 and CLEC4M were not significantly dysregulated in the lung compared with other organs (Figure 2C). This might be associated with the multiple functions of these receptors. ACE2 is not only the receptor for SARS-CoV-2, but also regulate the inflammation response. It could directly mediate the macrophage inflammatory function and its angiotensin converting activity may protect the lung from injury by controlling excess inflammation^4,5^. In this study, no difference of ACE2 in the lung between COVID-19 patients and control groups has been found, indicating that ACE2 inhibitors might not be an ideal therapy in the treatment for COVID-19^6^. On the other hand, we found that the ACE2 was downregulated in the kidney and heart of COVID-19 patients (Figure 2C), which might be associated with its catalytic activity in the local renin-angiotensin system (RAS) in these two organs^7^. CLEC4M is a highly expressed dendritic cell (DC) specific adhesion receptor in the liver, and it recognizes T cells to promote immune response^8^. CLEC4M was found to be downregulated in the liver of COVID-19 patients in this study (Figure 2C). It has been reported that the hepatocyte specific expression of CLEC4M could promote clearance of Von Willebrand factor (vWF) in mouse model^9^, which might be associated with hypercoagulability. CD209, in the same family with CLEC4M, however, showed no differential expression in neither organ. CD209 and CLEC4M are all associated with HCV infection^10^. On the contrary, CTSL was upregulated in the lung, spleen and renal medulla of COVID-19 patients, which has been reported to promote the SARS-CoV-2 entry and its inhibitor could suppress virus entry effectively^11^. The inhibitor of CTSL might be an effective drug for COVID-19 treatment. The above examples indicate that our data might complement the protein interaction data. The proteins which interact directly with virus could also be dysregulated at protein level, which might be potential targets for treatments.

### Other receptors

To search for other potential targets for COVID-19, we reviewed validated virus entry associated receptors and proteases^12^ and checked their abundance among the seven organs. Intracellular cholesterol transporter 1 (NPC1) binds to the glycoprotein of Ebola and its inhibitor has been recognized as a potential target for the treatment of Ebola^13^. NPC1 is significantly upregulated in most organs of COVID-19 patients, including the lung, white pulp of spleen, heart and thyroid (Figure 2C). Considering the critical need for lipid and its metabolism during viral replication^14^, NPC1 might be a potential drug target for the COVID-19. The carcinoembryonic antigen-related cell adhesion molecule 1 (CEACAM1), which is the major receptor for murine coronavirus on its host cells^15^, was significantly upregulated in the lung, liver, kidney, and thyroid (Figure 2C). Its dimerization with the immune checkpoint TIM3 would further suppress adaptive immunity^16^, so the inhibitor of CEACAM1 might be a potential target for COVID-19 treatment. In the hepatocytes, CEACAM1 could respond to insulin^17^, which may be associated with the hyperglycemia in the COVID-19 patient. In summary, this study might indicate the SARS-CoV-2 infection or potential drug target by the differential expression across eight organs between COVID-19 patients and the control group.

### Cluster 2: Transcription factors

Remarkably, we identified 1,107 transcription factors (TFs) among seven organs and 396 of them were differentially expressed between COVID-19 and control groups (B-H adjusted p value < 0.05 and |log_2_(fold-change, FC)| > log_2_(1.2), Table S5). By matching the experiential FC with the predicted activation state in upstream regulator analysis using Ingenuity Pathway Analysis (IPA), we found ten transcription factors expressed with the same trend, including nine upregulated proteins and one downregulated protein (SI Figure 1, Table S5).

Among the ten upregulated TFs (SI Figure 1), seven of them were involved in the inflammatory response, such as the acute phase response and NF-κB signaling, reflecting the extensive inflammatory infiltration in these tissues in COVID-19 patients. NF-κB subunit 2 (NFKB2) is expressed in various cell types and plays a key role in proinflammation^18^. It functions mainly through Toll-like receptor signaling pathway^19^ and NF-κB signaling pathway^20^. Transcription factor p65 (RELA) is a proto-oncogene and also encodes an NF-κB subunit protein. The heterodimeric RELA-NFKB1 complex has been reported to be the most abundant form of NF-κB, and plays a key role in immune, inflammatory and acute phase responses and affect cell proliferation and apoptosis^21^. In this study, RELA was upregulated in the thyroid and NFKB2 was elevated in the lung, kidney and thyroid (SI Figure 1), indicating a strong immune reaction in these three organs. CCAAT/enhancer-binding protein β (C/EBPB) is a transcription enhancing protein, which plays an important role in liver regeneration^22^, immune and inflammatory responses^22^. NF-IL6 is a member of the CCAAT/enhancer-binding protein (C/EBP) family and it is essential to induce G-CSF in macrophages and fibroblasts^23^. Upregulation of C/EBPB was detected in the lung, spleen, liver, heart and kidney (SI Figure 1), indicating potential immune dysregulation or liver damage. Signal transducer and activator of transcription 1 and 3 (STAT1/3) encode proteins of the STAT protein family. STAT is regulated and controlled by some cytokines and growth factors^24^ and then transmit the signal to downstream pathways such as cell growth and apoptosis^25,26^. STAT3 can contribute to the inflammation activation in tumors by NF-κB and IL-6-GP130-JAK pathways^27^. Upregulation of STAT3 was observed in the spleen, liver, heart and kidney, while STAT1 was only observed to be upregulated in the kidney (SI Figure 1). This may indicate potential dysregulation of immune state of COVID-19 patients. Transcription factor jun-B (JUNB) is a proto-oncogene encoding a transcription factor protein. It belongs to activate protein 1 (AP-1), which is regulated by varied physiological or pathological stimuli, such as cytokines, growth factors and infections^28^. It acts as a negative regulator of proliferation by modulating cell-cycle regulators^29^. Upregulation of JUNB was observed across over the lung, spleen, liver, heart, kidney and thyroid based on our analysis.

Other transcription factors either reflect the tissue injury or hypoxia state in COVID-19 patients in this study. The RB transcriptional corepressor 1 (RB1) was upregulated in the liver and kidney (SI Figure 1), which may negatively modulate the cell cycle^30^ and induce a higher degree of mitochondria permeabilization and apoptosis^31^. The La-related protein 1 (LARP1) was upregulated in the lung, spleen, liver, kidney and thyroid (SI Figure 1). LARP1 can regulate mRNA translation and is associated with mTOR signaling^32^, and it has been reported to interact with SARS-CoV-2^33^. The hypoxia-inducible factor 1-alpha (HIF1A) was upregulated in renal cortex (SI Figure 1) and was also predicted active in the lung, liver, renal cortex and medulla by upstream regulator analysis, reflecting the systemic hypoxia state^34^. Hepatocyte nuclear factor 4 alpha (HNF4A) can modulate gene transcription of lipid and bile acid synthesis, gluconeogenesis, and cytokines^35^. This function may be associated with the hepatic steatosis in COVID-19 patients because HNF4A was downregulated in the liver in this study (SI Figure 1).

### Cluster 3: Coagulation system

Blood coagulation occurred in most patients died from COVID-19 in our study with clotting in lower extremity vein (Table S1), which resulted from the induced coagulation cascade and the imbalance between coagulation, anticoagulation system and the impaired fibrinolytic system^36^.

The activities of inflammatory cells and cytokines following virus infection would result in endothelial injury and the secretion of tissue factors, which could initiate the coagulation cascade^37,38^. Tissue factor pathway inhibitor (TFPI) could inhibit the activated factor Xa (F10a), tissue factor catalytic complex and then inhibit the subsequent coagulation cascade^39^. In COVID-19 patients, TFPI was upregulated in the white pulp of the spleen and liver (SI Figure 2), which may occur with coagulation. Meanwhile, some coagulation factors were dysregulated in COVID-19 patients. For example, coagulation factor XII (F12) was downregulated in the heart, renal cortex and thyroid, coagulation factor XI (F11) was decreased in the lung, renal cortex and medulla while plasma kallikrein (KLKB1) was decreased in the lung, renal cortex and thyroid (SI Figure 2). F12 has been reported to be activated by KLKB1 and the activated F12 could catalyze F11. The activated F11 could activate factor IX (F9), which will lead to thrombosis and the formation of fibrin^40^. In COVID-19 patients, F12 was decreased in the heart, renal cortex and thyroid. Coagulation factor XIIIa (F13a) is activated in the last step of coagulation, which would induce hemostasis. In addition, it would stabilize the fibrin clot to avoid fibrinolysis^41^. It was increased in the renal cortex and thyroid, which could contribute to the formation of blood clotting. vWF is a glycoprotein, which could bind to the factor VIII (F8) and protect F8 from degradation by Vitamin K-dependent protein C (PROC). Besides, it could mediate the platelet aggregation following the vascular injury^42^. The VWF was increased in the white pulp of the spleen and renal cortex, which indicates the high risk of thrombosis risk. In contrast, it was decreased in the heart and lung of the COVID-19 patients Von Willebrand factor (VWF). Fibrinogen alpha chain, gamma chain and beta chain (FGA, FGG and FGB), which could be cleaved to fibrin and generate the blood clots^43^, were all increased in the lung and all decreased in the thyroid, while FGA was only increased in the renal cortex (SI Figure 2). The prothrombin (F2) was decreased in the heart (SI Figure 2).

The Vitamin K-dependent protein S (PROS1) and PROC belong to the protein C anticoagulant system. PROS1 is the cofactor of PROC, which could degrade factor VIIIa (F8a), factor Va (F5a) and protease-activated receptor 1 (PAR-1) once activated^44^. Alpha-1-antitrypsin (SERPINA1) is an inhibitor of PROC^45^. Plasma serine protease inhibitor (SERPINA5) is also a serine proteinase inhibitor and acts as an inhibitor of protein C and other coagulation enzymes^46^. In the COVID-19 patients, PROC was decreased in the heart and PROS1 was decreased in the kidney (SI Figure 2), which may contribute to the thrombosis. SERPINA1 is also decreased in the heart and thyroid. SERPINA5 is decreased in the liver, heart, renal cortex and thyroid. Antithrombin Ⅲ (SERPINC1) inhibits thrombin and several serine proteases, such as factors IXa (F9a), Xa (F10a), XIa(F11a), XIIa (F12a), plasmin and kallikrein. It would consume rapidly during disseminated intravascular coagulation (DIC)^47^. In the COVID-19 patients, SERPINC1 was decreased in the heart, renal medulla and thyroid (SI Figure 2). Thrombomodulin (THBD) is also an anticoagulant protein, which could inhibit fibrinogen clotting and FV activation^48^. It is increased in the white pulp of spleen, heart and renal cortex (SI Figure 2). Heparin cofactor 2 (SERPIND1) is a serine proteinase inhibitor, which acts as the inhibitor of thrombin^49^ and the cofactor for heparin. In the COVID-19 patients, SERPIND1 significantly decreased in the liver, heart, renal cortex, medulla and thyroid (SI Figure 2), which could lead to the generation of thrombin or the procoagulant state in the body.

In the fibrinolytic system, plasminogen (PLG), the zymogen, could convert into serine proteinase plasmin by two serine peptidases, the tissue plasminogen activator (PLAT) and urokinase plasminogen activator (uPA)^50^. Then the plasmin could breakdown the blood clotting. In this study, PLG was downregulated in the heart and thyroid while PLTA was upregulated in the liver of the COVID-19 patients (SI Figure 2). Alpha-2-antiplasmin (SERPINF2) was downregulated in the heart, renal medulla and thyroid of the COVID-19 patients (SI Figure 2), which may indicate the increased fibrinolysis^51^. Plasminogen activator inhibitor 1 (SERPINE1) is a serine proteinase inhibitor and acts as a major inhibitor of plasminogen activators. In the COVID-19 patients, SERPINE1 significantly upregulated in the lung, liver, heart, renal cortex and medulla (SI Figure 2), which could increase the risk of thrombosis and fibrosis^52^. Alpha-2-macroglobulin (A2M) could bind to varied proteinases, such as plasmin and kallikrein, and inactive them. And in sepsis, it would decrease because of the formation of complex^53^. Here, we found A2M was decreased in lung, heart, renal medulla and thyroid (SI Figure 2).

### Cluster 4: Cytokines

When the SARS-CoV-2 virus enters the lung mediated by ACE2 or other receptors through the superior respiratory system, it induces the secretion of inflammatory cytokines and transmits the information from lesions to the whole body through the circulatory system. Then the inflammation cells are attracted to the infection sites. Meanwhile, the hyperinflammatory state harms the alveolar cells and influences multiple organs. The damage of the integrity of the gas exchange barrier may induce hypoxia, which strengthens inflammation response^54^ in the whole body. The following paragraphs described the significantly dysregulated protein associated with cytokines.

Our data showed that nicotinamide phosphoribosyl transferase (NAMPT) was upregulated in the six organs (except for testis) of COVID-19 patients (SI Figure 3). NAMPT is involved in the resistance of cells to oxidative stress and may help immunity cells’ survival under stress such as inflammation^55^. Extracellular NAMPT (eNAMPT) was identified as the pre-B cell colony enhancing factor^56^ and an essential factor in granulocyte-colony-stimulating factor-(G-CSF)-induced myeloid differentiation^57^, which also could promote B cell maturation and inhibit neutrophil apoptosis^58^. The overexpression of NAMPT in various organs may indicate that the COVID-19 patients are under activated immune response.

Glucocorticoid receptor (NR3C1) was significantly downregulated in five organs (except for testis and thyroid) in our study (SI Figure 3). NR3C1 usually exists in the cytoplasm and would be translocated to the nucleus after binding to its ligand. It could upregulate the anti-inflammatory protein or inhibit the expression of pro-inflammatory protein in the nucleus^59^. The significant downregulation of NR3C1 in the five organs may indicate an increased inflammatory response of various organs after SARS-CoV-2 infection.

Our data indicated that interleukin-6 receptor subunit beta (IL6ST) was significantly upregulated in COVID-19 patients’ thyroid (SI Figure 3). IL6ST is involved in the signal transduction of IL-6-type cytokines. After binding to its receptor, IL6ST is driven to form a complex by IL6, then activating Janus kinases (JAKs) and STAT3 and participating in acute phase response, T cell differentiation, antibody production and proliferation^60,61^. IL6 is a key pro-inflammatory factor in the early stage of infection and is associated with prognosis^60^. It has also been reported to be overexpressed in the serum of severe COVID-19 patients^62^.

The abundance of angiopoietin like 6 (ANGPTL6) in the thyroid tissue of COVID-19 patients was significantly downregulated (SI Figure 3). ANGPTL6 plays a role in angiogenesis, lipid metabolism and glucose metabolism^63^. It has been reported that in the ANGPTL6 deficient mouse model, obesity, lipid accumulation and insulin resistance were developed. In contrast, the mouse would be more sensitive to insulin by activating the ANGPTL6, which indicates that ANGPTL6 is related to insulin resistance^64^. The downregulation of ANGPTL6 in the thyroid of COVID-19 patients indicates a potential insulin resistance status in COVID-19 patients.

Our data showed that the expression of natriuretic peptides A (NPPA) in the heart tissue of COVID-19 patients was significantly downregulated (SI Figure 3). NPPA belongs to the natriuretic peptide family, which regulates the extracellular electrolyte homeostasis. The cardiac natriuretic peptide, also known as vasodilation, is mainly produced, stored, and secreted by atrial cardiomyocytes. When the blood volume increases and the heart or blood vessel wall is stimulated to release atrial natriuretic peptide by a large stretch which causes powerful natriuretic and diuretic effects. The knockout of NPPA receptor (NPR1) in the heart of mice exhibited cardiac hypertrophy^65^ and the knockout of NPPA could cause heart failure and hypertension^66^. The downregulation of NPPA implies potential heart damage in COVID-19 patients.

Downregulation of fibroblast growth factor 2 (FGF2) was detected in the liver of COVID-19 patients (SI Figure 3). FGF2 is a growth factor that belongs to the heparin-binding (fibroblast) growth factor family. FGF2 can induce angiogenesis by promoting endothelial cell proliferation and transferring its physical organization^67^ and is involved in the regulation of many other important biological processes such as cell survival and cell differentiation^68^. The decrease of FGF2 may contribute to the liver fibrosis of COVID-19 patients.

Fibroblast growth factor 17 (FGF17) was significantly overexpressed in the heart of COVID-19 (SI Figure 3). FGF17 belongs to the family of fibroblast growth factor (FGF), which participates in a variety of biological processes, including cell proliferation, cell migration during embryonic development, wound healing immune responses, cell differentiation and angiogenesis^67^. The upregulation of FGF17 may indicate a repair status of heart damage in the patients.

In the renal medulla of COVID-19 patients, IL13RA1 was significantly upregulated (SI Figure 3). IL13RA1 is a subunit of the interleukin 13 receptor, which mainly mediates IL-4 signal transduction. IL-4 and IL-13 both belongs to the profibrogenic cytokines^69^. It has been reported to protect the lung from bleomycin induced injury^70^.

Interferon-gamma receptor 1 (IFNGR1) was overexpressed in multiple kinds of organs except for the thyroid and testis (SI Figure 3). IFNGR1 is a ligand-binding chain of heterodimeric gamma interferon receptors on macrophages and is a functional receptor for interferon-gamma^71^, which triggers host immunity to virus infection^72^, such as induction of phagocyte oxidase system, NO production and lysosomal enzymes for microbe destruction in macrophages^73^. The overexpression status of IFNGR1 implies the systemic immune response activated by SARS-CoV-2.

Interleukin 1 receptor-like 1 (IL1RL1) was found to be upregulated in the kidney cortex of COVID-19 patients in this study (SI Figure 3). IL1RL1 belongs to the interleukin 1 receptor family, which is a receptor for interleukin-33 (IL-33)^65^. IL-33 signal transduction could lead to the activation of NF-κB, MAP kinase and induction of Th2 cytokines^66^. It has been reported that IL1RL1 was overexpressed in aged SARS-CoV infected animals, and the overexpression of IL1RL1 indicates that older macaques have a stronger host response upon viral infections than young macaques^74^. This may indicate that a pro-inflammatory status in COVID-19 patients.

Eosinophil-derived neurotoxin (RNASE2) was found to be downregulated in the COVID-19 patient’s kidney cortex (SI Figure 3). RNASE2 is derived from eosinophils and has antiviral activity and selective chemotactic properties to dendritic cells^75^. It can reduce the activity of single-stranded RNA virus through ribonucleolytic enzyme activity and interaction with the extracellular virus by its unique structure^76^. The downregulation of RNASE2 in the renal cortex of COVID-19 patients may indicate dysregulation of the immune system or may be a result of consuming itself to binding with and inhibit extracellular virus in COVID-19 patients.

Syndecan-4 (SDC4) was upregulated in the liver of the COVID-19 patients (SI Figure 3). SDC4 is a membrane proteoglycan. It can support exosome biogenesis and engagement of FGF with its receptor, which plays a role in angiogenesis, inflammation and infection^77^.

C-X-C motif chemokine 13 (CXCL13) was observed to be upregulated in the spleen of CVOID-19 patients (SI Figure 3). CXCL13 is a member of the CXC chemokine family, which interacts with the chemokine receptor CXCR5 to selectively chemotactic for B cells^78^.

CXCL13/CXCR5 enhances antigen encounter and BCR-mediated B-cell activation through regulation of actin cytoskeleton and activity of motor protein^79^. The upregulation of CXCL13 may indicate the elevated humoral immune response in the spleen of COVID-19 patients.

Cathelicidin antimicrobial peptides (CAMP), an antimicrobial peptide family member, was downregulated in the spleen of the COVID-19 patients (SI Figure 3). When mammals encounter a large number of bacterial invasions, it plays a role in the innate immune defense by inducing leukocyte chemotaxis, degranulation, the response of macrophages and stimulating wound healing^80^. The downregulation of CAMP may indicate the dysregulation of innate immunity in COVID-19 patients.

Retinoic acid receptor RXR-alpha (RXRA) was detected to be downregulated in the lung of the COVID-19 patients (SI Figure 3). RXRA is a nuclear receptor for retinoic acid^81^ and regulates lipid metabolism and inflammation in macrophages^82^. In response to viral infections, it plays a role in weakening the innate immune system. It has been reported that the downregulation of RXRA would induce the production of type I IFN and antiviral response^83^. Downregulation of RXRA may indicate a proinflammatory state in the lung of COVID-19 patients.

### Cluster 5: Angiogenesis

Angiogenesis is the forming of new blood vessels from existing vessels by sprouting and splitting. Both types are discovered in almost all the organs in human bodies^84^. The normal regulation of angiogenesis is governed by a fine balance between factors that induce the formation of blood vessels, such as pro-angiogenic factors and those that halt or inhibit the process. Generally, cells activated by hypoxia would release pro-angiogenic molecules, such as hypoxia-inducible factors (HIF)^85^, vascular endothelial growth factor (VEGF)^86^ and angiotensin (Ang)^87^. In response to the pro-angiogenic factors, the endothelial cells would proliferate to form the neighboring blood vessels and secret matrix metalloproteases (MMP) to digest the blood vessel walls for migration. Several protein fragments are produced by the digestion of the blood vessel walls, which would intensify the proliferative and migratory activity of endothelial cells and then form a capillary tube by altering the arrangement of their adherence membrane proteins, such as vascular cell adhesion molecule (VCAM) and integrin^88^. Finally, new blood vessels become mature and stabilized by Ang, PDGF (platelet derived growth factor) and transforming growth factor beta (TGFB), resulting in a continuous blood flow^89^.

It is known that hyperbaric oxygen therapy increases the expression of HIF and actives VEGF production^90^. Thereby, we can speculate that severe COVID patients with ARDS after ventilator treatment would be pro-angiogenesis. However, the angiogenesis may not be specific to COVID cases. Recently, a study which compared tissues from seven lung samples obtained from patients who died from COVID-19, tissues from 7 lungs obtained from ARDS secondary to influenza A(H1N1) infected corpses and tissues from 10 age-matched uninfected lungs found that the amount of new vessel generated by intussusceptive angiogenesis was 2.7 times as high as that in the lungs from patients with influenza^91^. These results support a more profound evidence of pro-angiogenesis in COVID-19 patients.

It is worth noting that 139 proteins involved in seven organs are related to angiogenesis by our proteomic data analysis. It implies that the angiogenesis process has differences between COVID-19 patients and healthy controls.

As a supplement to that study, our multi-organ data found a multi-organ angiogenesis process, not merely in the lung. The vascular endothelial growth factors A (VEGFA) was upregulated in the thyroid, and the VCAM1 was upregulated in the kidney (cortex as well as medulla) of COVID-19 (SI Figure 4). The MMP protein 14 were found upregulated in the spleen, lung and thyroid, and the MMP2 were induced in thyroid, lung and renal cortex in COVID-19 cases (SI Figure 4). We also quantified transforming growth factor beta related proteins (TGFBI) and angiogenin (ANG) in our proteomics data, while it was not upregulated in all of these COVID organs (SI Figure 4).

Besides the proteins that directly participate in angiogenesis, we also detected some potential regulatory ones. Firstly, the upstream hypoxia inducible factor 1 subunit alpha (H1F1A) was found to be increased in the kidney of COVID-19 cases, which actives VEGF production and angiogenesis. In addition, the integrin proteins (ITGA5, ITGB1, and ITGB3) are found up-regulated in the lung, heart, liver or renal cortex (SI Figure 4). Many molecules participate in the processes of angiogenesis, endothelial cells proliferation and invasive signal. Integrins are the principle adhesion receptors in forming extracellular microenvironment, and integrin-mediated interactions mainly regulate cell proliferation, migration, and survival^92^. The chemokine CX3CL1 could induce the monocytes accumulation and angiogenesis during inflammation^93^. Our data also found the overexpression of CX3CL1 in COVID-19 patients’ thyroid (SI Figure 4), which may indicate the angiogenesis potential in thyroid. It is found that SERPIN family is anti-angiogenesis. In the rat, blockage of SERPIN family would mediate the process of angiogenesis by interleukin 8 (IL-8) and VEGF. In assays of cellular events in angiogenesis, SERPIN family blocked the processes of invasion, migration and tube formation^94^. Our results showed a downregulation pattern of SERPIN family proteins in multiple organs of COVID-19 cases. The SERPINA1, SERPINF1, and SERPING1 are significantly reduced in heart (SI Figure 4). Besides, SERPINA1 was also found downregulated in the thyroid, and SERPING1 are found downregulated in the renal medulla and liver (SI Figure 4). Transmembrane Protein 100 (TEME100) is an intracellular transmembrane protein. It is involved in the differentiation of arterial endothelium and vascular morphogenesis, which can be activated by AKT1 receptor^95^. In addition, TEME100 plays a vital role in maintaining vascular integrity and vascular genesis^96^. In our proteomic data, TEME100 were downregulated in the lung of COVID-19 patients (SI Figure 4). It hints that the blood vessels of the lung in COVID patients might have poor stability, resulting in increased risk of angiogenesis in the lung under weak vascular integrity.

In conclusion, we suspected that the balance of angiogenesis is dysregulated in COVID-19 patients, which results in pathological angiogenesis and causes an increased blood-vessel formation.

### Cluster 6: Fibrosis

#### Fibrosis markers

The fibrosis process is associated with the overexpressed extracellular matrix, collagen proteins and proliferation of fibroblasts^97^. COL1A1 and COL1A2 are pro-alpha1 and 2 chains of type I collagen, and COL3A1 is collagen type III alpha 1 chain. All of them belong to fibril-forming collagens, which are produced by fibroblasts and act as the essential components of the myocardial interstitial matrix. These three proteins were downregulated in heart tissue samples from COVID-19 patients (SI Figure 5). It is reported that an increased expression of collagen type I and III are believed to be related to fibrosis^98,99^. Our control samples of heart were collected from patients with dilated cardiomyopathy (DCM), in which interstitial fibrosis is one of the major characteristics^100^. It is also reported that collagen type I and III were both elevated in patients with DCM^101^. Due to the potential fibrosis status in our control groups, minor heart fibrosis can be missed even if it exists in COVID-19 patients.

LAMB1, LAMB2 and LAMC1 are laminin subunits. Laminins are found in basement membrane as ECM associated glycoproteins. It is reported that laminins are good prognostic markers for liver fibrosis^102,103^. Though no significant change of expression level of laminins was observed in the liver, we noticed upregulated LAMB1 and LAMC1 in the white pulp of spleen, and downregulated LAMB2 and LAMC1 in renal medulla (SI Figure 5). It is reported that increased matrix metalloproteinase 7 and 9 (MMP7 and MMP9) are closely related to the development of fibrotic kidneys, and laminins are the substrate of MMP7^104^ and MMP9^105^. Although we didn’t find significant upregulation of MMP7 and MMP9, the downregulation of LAMB2 and LAMC1 may indirectly indicate renal fibrosis.

Matrix metalloproteinases (MMPs) and metalloproteinase inhibitors (TIMPs) maintain the balance of fibrogenesis and fibrosis^106,107^. In our dataset, we identified MMP2, MMP9, MMP14, IMMP2L, MMP28, TIMP1,2 and 3. They are all identified upregulated over the different organs (SI Figure 5). Fibrosis in pathological was only found in the lung, in which two kinds of inhibitors were upregulated. It might indicate a positive feedback loop between MMPs and TIMPs.

The pulmonary fibrosis, which is associated with fibrous exudation, hyaline membrane formation and alveolar space collapsed. We have identified the upregulation of lung fibrosis biomarkers including Chitinase-3-like protein 1 (CHI3L1)^108^ and pulmonary surfactant associated protein B (SFTPB) (SI Figure 5). CHI3L1 was upregulated in the widest distributed among five tissues including the lung, heart, liver, and kidney (SI Figure 5). SFTPB promotes the secreted alveolar surfactant and sustains alveolar structure^109^. The SFTPB was upregulated in the lung of COVID-19 patients. FABP family is similar to the surfactant^110^. We found FABP3, FABP1 and FABP5 were downregulated based in our dataset (SI Figure 5). The downregulated FABPs indicated the potential of fibrosis.

Lipid metabolism is also associated with fibrosis. The fibrosis influenced by lipid metabolism is the transformation between lipofibroblast and myofibroblast. TGF-beta can induce lipofibroblast differentiation to myofibroblast which is the process of fibrosis generation^111^. The markers of lipofibroblast can be the potential markers for fibrosis, such as perilipin-2 (PLIN2). In COVID-19 patients, PLIN2 upregulated in the lung, liver, heart, and spleen (SI Figure 5).

#### Fibrosis associated proteins

Fibrosis could be divided into four stages^69^. At the initiation stage, the cell and organ damage would initiate a cascade of stress and immune responses. In response to the initial stress, multiple inflammatory signaling pathways were activated, such as the chemokine signaling, complement system, macrophage activation, NF-κB signaling, interferon, platelet and neutrophil degranulation. Thirdly, chronic inflammation would drive the differentiation or proliferation of fibroblasts and wound-healing response. Fourthly, the extracellular matrix of immune cells and fibroblast cells would be further modified. Here we compared the dysregulated proteins in our studies with proteins in these pathways of the fibrosis and selected the top 20% dysregulated proteins with the highest fold changes in a specific organ for discussion.

##### Stage 1: Initiation

In COVID-19 patients, alcohol dehydrogenase 1B (ADH1B) was downregulated in the lung and liver, while it was upregulated in the white pulp of spleen, heart and thyroid. Alcohol dehydrogenase 6 (ADH6) was downregulated in the lung and renal cortex. Aldehyde dehydrogenase family 3 member A2 (ALDH3A2) was downregulated of in the liver, heart, and renal cortex. Alcohol dehydrogenase 1C (ADH1C) was downregulated in the liver. Alcohol dehydrogenase 4 (ADH4) was downregulated in the liver while it was upregulated in the heart (SI Figure 6). They all belong to the alcohol dehydrogenase family and are involved in the metabolic process of multiple substrates, such as ethanol, hydroxysteroids, and lipid peroxidation products. It has been reported that lipid metabolism dysregulation can induce hepatofibrosis in mouse model^112^. The downregulation of these proteins in the liver of COVID-19 patients might indicate the potential hepatic fibrosis. Moreover, the downregulated ALDH3A2 in COVID-19 patients, may lead to the accumulation of lipid precursors in the liver^113^. Besides, ADH1B, ADH6 are downregulated in the lung and kidney of the COVID-19 patient, respectively.

In COVID-19 patients, fatty acid synthase (FASN) was upregulated in the lung, renal cortex and thyroid, while it was downregulated in the liver and testis (SI Figure 6). It is a key lipogenic enzyme in de novo biogenesis of fatty acids^114^ and FASN is required for profibrotic TGF-β signaling^115^, which is a master regulator of fibrosis^116^. Compared with non-COVID-19 controls, the upregulation of FASN in the lung, kidney and thyroid and the downregulation of FASN in the liver and testis in COVID-19 patients suggest the initiation of fibrosis in lung, kidney and thyroid along with the impairment of the fatty acid synthesis process in the liver and testis in COVID-19 patients.

Fructose-1,6-bisphosphatase (FBP1) was downregulated in the white pulp of spleen, liver and renal cortex in COVID-19 patients (SI Figure 6). It is a tumor-suppressive function gene, which encodes a rate-limiting gluconeogenic enzyme. Reduced expression of FBP1 was observed by both quantitative real-time polymerase chain reaction(qRT-PCR) and Western Blot with increased liver fibrosis score in mouse^117^. More dramatic fibrosis can be histologically detected in FBP1-deficient liver^118^. The downregulation of FBP1 in spleen, liver and kidney of COVID-19 patients compared with non-COVID-19 controls suggests dysfunction of glucose metabolism and fibrosis potential in these tissues.

Lysosomal acid lipase (LIPA) was upregulated in the heart of COVID-19 patients (SI Figure 6). It is located in the lysosome to catalyze the hydrolysis of cholesteryl esters and triglycerides. The deficiency of LIPA will lead to the accumulation of fat and cause cholesterol ester storage disease (CESD). The accumulation of cholesteryl esters and triglycerides in hepatocytes result in hepatomegaly with progressive fibrosis and liver cirrhosis^119^. Our data showed no significant difference in LIPA expression in the liver but upregulation in the heart, which might indicate the fibrosis initiation in the heart.

Phospholipid transfer protein (PLTP) was upregulated in the liver of COVID-19 patients (SI Figure 6). It is responsible for lipid transfer and it can transfer phospholipids from triglyceride-rich lipoprotein to high-density lipoprotein (HDL) and regulate lipid metabolism^120^. The liver is one of the main sites for the production and degradation of lipoprotein and the expression of PLTP. Studies have shown that PLTP can amplify the pro-inflammatory effect of lipopolysaccharide (LPS). In the mouse model, overexpression of PLTP can induce atherosclerosis^121^. The overexpression of PLTP in COVID-19 liver tissue may be associated with potential liver injury.

Acyl-CoA desaturase (SCD) was upregulated in the lung of COVID-19 patients (SI Figure 6). It exists in the endoplasmic reticulum and is mainly involved in fatty acid biosynthesis. SCD is involved in inflammation and stress regulation of different cell types, including adipocytes, macrophages, endothelial cells, and muscle cells^122^. Upregulated SCD expression in the mouse model promotes hepatic fibrosis through Wnt/β-catenin signal and the lipid metabolism^123^. Here, the upregulated SCD in the lung tissue of COVID-19 patients may affect the pulmonary fibrosis process by regulating metabolism reprogramming.

Protein transport protein Sec61 subunit beta (SEC61B) was upregulated in the lung, white pulp of spleen and kidney of COVID-19 patients (SI Figure 6). It is one of the two subunits of SEC61 complex which is the core component of the protein transport device in the endoplasmic reticulum and is responsible for the transport of polypeptides across the endoplasmic reticulum. SEC61B is associated with the cytosolic fibrosis through dysregulation of endoplasmic reticulum^124^. The upregulated SEC61B might indicate the fibrosis initiation in multiple COVID-19 organs.

Transforming Growth Factor Beta Receptor 2 (TGFBR2) was downregulated in the lung, while it was upregulated in the heart of COVID-19 patients (SI Figure 6). It can form a heterodimer with TGF- receptor type 1 and bind TGF to participate in the transcriptional regulation of genes related to cell cycle, wound healing, immune suppression, and tumor formation. TGF signaling mediates fibrosis after the inflammatory damage in the heart, kidney, and intestines^125^. TGFBR2 is also a potential target for inhibition of pulmonary fibrosis^126^.

##### Stage 2: Inflammation

Aldo-keto reductase family 1 member B10 (AKR1B10) was upregulated in the renal cortex of COVID-19 patients (SI Figure 7). It belongs to AKR family 1 subfamily B (AKR1B)^127^. Recent studies have shown that AKR1B10 is secreted into the stroma from liver cells by binding to heat shock protein 90 and maybe a useful serum biomarker for advanced fibrosis in nonalcoholic steatohepatitis (NASH)^127,128^. The upregulation of AKR1B10 might indicate the potential fibrosis in the renal cortex.

Delta-aminolevulinic acid dehydratase (ALAD) was downregulated in the lung, white pulp of spleen, liver, renal cortex and thyroid of COVID-19 patients (SI Figure 7). It is the principal lead binding protein in erythrocytes, which carry over 99% of the lead in blood^129^. It is involved in the biosynthesis of heme^130^. Heme is a functional group of many heme proteins, including cytochromes and hemoglobin (Hb), so it is essential for many different cellular processes^131^. Excess free heme has been shown to exacerbate the pathogenesis of various inflammatory diseases, such as sepsis, malaria, sickle cell disease, kidney disease, and multiple organ failure^131–134^. Chronic obstructive pulmonary disease (COPD) patients show increased cell-free Hb, correlating with disease severity^135^. However, we identified ALAD was significantly downregulated in the multiple organs of COVID-19 patients.

Apolipoprotein was downregulated in COVID-19 patients compared with non-COVID-19 controls, mainly in liver, heart, kidney and thyroid (SI Figure 7). Based on the ability of apolipoprotein to inhibit inflammation, oxidative stress, and tissue remodeling, as well as to promote adaptive immunity and host defense, the emerging role of apolipoprotein in the pathogenesis and treatment of lung diseases is increasingly recognized^136^. Apolipoprotein A-I (ApoA-I), apolipoprotein A-II (ApoA-II), and apolipoprotein B (ApoB) are important components of lipoprotein particles that facilitate the transport of cholesterol, triglycerides, and phospholipids between plasma and cells^137^. Studies showed that an ApoA-I/ABCA1-dependent pathway inhibited experimental ovalbumin-induced neutrophilic airway inflammation by reducing colony-stimulating factor (G-CSF) production^138^. ApoA-I was first proposed to play a role in the pathogenesis of idiopathic pulmonary fibrosis (IPF). Based on proteomic analysis of bronchoalveolar lavage fluid (BALF), the ApoA-I levels are reduced compared to healthy subjects, while the treatment with ApoA-I would help to decrease the bleomycin-induced inflammatory cells and collagen deposition, which reflects the negative correlation between apolipoproteins and fibrosis^139^. Based on our analysis, apolipoproteins were significantly decreased in six tissues except for the testis to certain degrees, indicating a pro-fibrosis potential in different organs in patients with COVID-19 infection. Furthermore, Apolipoprotein C-II (ApoC-II) is a small exchangeable apolipoprotein found in triglyceride-rich lipoproteins (TRL), such as chylomicrons (CM), very-low-density lipoproteins (VLDL), and in high-density lipoproteins (HDL)^140,141^. ApoC-II plays a critical role in TRL metabolism by acting as a cofactor of lipoprotein lipase (LPL), the main enzyme that hydrolyzes plasma triglycerides (TG) on TRL^142^. The reduction of ApoC-II based on our analysis further indicated the reduced metabolic level of triglycerides-rich lipoproteins in patients with COVID-19 infection.

Antioxidant 1 (ATOX1) was downregulated in the white pulp of spleen, renal cortex and thyroid of COVID-19 patients (SI Figure 7). It plays a crucial role as a copper chaperone^143–145^. It is worth mentioning that ATOX1 was found to have a pivotal role in inflammation via inhibition of inflammatory responses. The decrease of ATOX1 suggested the occurrence of prophase inflammation^145^, which might indicate an inflammation stage before fibrosis in these organs.

Complement component 3 (C3) was downregulated in the lung, heart and thyroid in COVID-19 patients, but it was upregulated in the white pulp of spleen (SI Figure 7). The complement system is a crucial part of the immune system that is involved in the pathogenesis of various inflammatory lung conditions, including infectious disease and chronic obstructive pulmonary disease^146–149^. Complement component 3 (C3) plays a central role in the activation of the complement system^149–151^. Additionally, C3-targeted intervention may provide broader therapeutic control of complement-mediated inflammatory damage in COVID-19 patients^152^.

Catalase (CAT) was downregulated in the lung, white pulp of spleen, liver and renal cortex of COVID-19 patients (SI Figure 7). It is an important antioxidant enzyme in the process of defense against oxidative stress^153^. It is preferentially expressed in the alveolar epithelial cells. The downregulated CAT has been reported in the fibrotic lungs of human, which may be associated with the protective role of CAT against inflammation and subsequent fibrosis associated processes, such as TGF-β expression and collagen content^154^.

The downregulated CAT may trigger the proinflammation state and contribute to the fibrosis process in these organs.

Monocyte differentiation antigen (CD14) was upregulated in the white pulp of spleen and renal cortex of COVID-19 patients (SI Figure 7). It is a surface antigen and preferentially expressed on monocytes or macrophages, which plays multiple pivotal biological roles, such as innate immune response to binding bacterial lipopolysaccharide^155^. Additionally, it has been found that CD14 was highly expressed in the lung, liver, spleen, and its expression was dynamically changed in the course of inflammation^156^. Another study showed that systematic inhibition of CD14 could reduce organ inflammation, which was tested in lung, liver, spleen, and kidneys. They also reported that ICAM-1 and VCAM-1 were significantly inhibited in the kidneys by inhibiting CD14^157^.

Chitinase-3-like protein 1 (CHI3L1) was upregulated in the lung, liver, heart, renal cortex and medulla of COVID-19 patients (SI Figure 7). It is a member of chitinase family^158^ and is highly expressed in the liver^159^. In arthritic articular cartilage, CHI3L1 functions as a growth-factor for fibroblasts^160^. In the bleomycin-treated mice, it would decrease transiently due to its protective role in the injury. Then it would increase and play a profibrotic role in activating macrophages and inducing the fibroblast and matrix deposition^108^. The upregulated CHI3L1 in multiple organs of COVID-19 patients may indicate a profibrotic state.

C-reactive protein (CRP) was upregulated in six organs (except testis) of COVID-19 patients (SI Figure 7). Fibrosis is caused by scar tissue formation in internal organs. Two closely related human serum proteins, serum amyloid P (SAP), and C-reactive protein (CRP), strongly affect fibrosis^161^. In multiple animal models, and even Phase 1 and Phase 2 clinical trials, CRP indicates the potential of fibrosis^162,163^. It’s worth noting that CRP was significantly upregulated in almost all of organs based on our analysis. This prompts us that the follow of CRP levels could potentially indicate the fibrogenesis status of COVID-19 patients.

FGB and FGG were upregulated in the lung but downregulated in the thyroid of COVID-19 patients (SI Figure 7). They are both blood-borne glycoproteins and components of fibrinogen, which play a role in vascular injury and dysregulated in thrombophilia^164^, as well as a proinflammatory role under several pathologic conditions including pulmonary and kidney fibrosis^165^. Fibrinogen was substantially higher in COVID-19 patients than those in healthy controls^166^. Meanwhile, in the process of acute phase response, interleukin-6 (IL-6) could induce upregulation of the three fibrinogens, FGA, FGB and FGG, in liver and lung epithelium^167^. Consistent with fibrosis observed in the lung of COVID-19, FGB and FGG were evaluated in lung but decreased in thyroid.

Glutathione Peroxidase 2 (GPX2) was found to be elevated in the liver of COVID-19 patients (SI Figure 7). GPX2 is a glutathione peroxidase, which catalyzes the reduction of hydrogen peroxide (H_2_O_2_) by glutathione. Oxidative stress initiates epithelial cell injury and fibrogenesis. Hence decreasing of glutathione results in hepatic fibrosis^168^ and pulmonary fibrosis^169^. Elevated GPX2 in the liver predicts fibrogenesis in the liver of COVID-19 patients.

Intercellular adhesion molecule (ICAM)-1 was upregulated in the spleen, liver, heart, kidney and thyroid of COVID-19 patients (SI Figure 7). ICAM1 is a member of the immunoglobulin (Ig) superfamily expressed on fibroblasts and epithelial cells. ICAM deficiency suppresses cytokine production, white cell infiltration and attenuates fibrogenesis^170^. The upregulation of ICAM1 indicates the occurrence of inflammation and fibrosis.

Protein tyrosine phosphatase non-receptor type 1 (PTPN1) was upregulated in five organs (except the heart and testis) of COVID-19 patients (SI Figure 7). It is the founding member of the protein tyrosine phosphatase (PTP) family. PTP1B was upregulated in the fibrotic liver, which may be induced by TGF-β1^171^. Based on our analysis, the elevated level of PTPN1 can be observed in the lung, spleen, liver, kidney and thyroid, indicating a potential fibrosis status in these organs.

Serum Amyloid A1(SAA1) was found to be significantly upregulated in the lung, liver, heart and kidney of COVID-19 patients (SI Figure 7). SAA1 is a major acute phase protein that is highly expressed in response to inflammation and tissue injury^172^. Significant upregulation of SAA1 may indicate an inflammation status in these organs, which may contribute to fibrosis in the end.

Superoxide dismutase 1(SOD1) was found to be down-regulated in the spleen, heart, kidney and thyroid of COVID-19 patients (SI Figure 7). SOD1, a copper and zinc binding protein is an enzyme for cleaning free superoxide radicals. Superoxide dismutase can act as a potential antifibrotic drug for Hepatitis C related fibrosis^173^. In our proteomics data, the downregulation of SOD1 indicates a potential fibrosis status in these organs.

Vascular cell adhesion molecule 1(VCAM1) was upregulated in the kidney of COVID-19 patients (SI Figure 7). It is a cell surface sialoglycoprotein expressed by cytokine-activated endothelium. VCAM1 is upregulated in IPF, which is responsive to TGF- β1^174^. The upregulation of VCAM1 in the kidney may indicate the fibrosis potential in COVID-19.

##### Stage 3: Proliferation

β-arrestin1 (ARRB1) was downregulated in the lung, white pulp of spleen, liver and renal medulla of COVID-19 patients (SI Figure 8). It has been demonstrated to function as a molecular scaffold that regulates cellular function by interacting with other partner proteins, and involved in multiple physiological processes including immune response, tumorigenesis and inflammation^175^. The β-arrestins are universally expressed, but the neural and immune systems have a much higher expression level. Previous studies have reported that ARRB1 specifically regulates the survival and homeostasis of CD4+ T cells and thus affects the adaptive immune responses^176^. The downregulation of ARRB1 in lung, spleen, liver and kidney in COVID-19 patients further suggests an end-stage immune response in COVID-19 patients. In addition, ARRB1 has been reported to stimulate several signaling pathways involved in fibrosis, such as TGF-β pathway. These pathways would affect cell growth, extracellular matrix deposition and activation of inflammation^177^. The dysregulated ARRB1 may participate in the fibrosis process in COVID-19.

Cadherin 2 (CDH2) was downregulated in the renal cortex of COVID-19 patients (SI Figure 8). It can mediate the adhesion between homotypic cells by binding to the CDH2 chain on another cell’s membrane. In the process of IPF, CDH2 was transformed into CDH11, leading to the aggravation of pulmonary fibrosis^178^. Our data show that the expression of CDH2 in the kidney of COVID-19 patients is decreased. However, the reduction of CDH2 in the renal cortex may be due to the conversion of CDH2 to CDH11, which needs further investigation.

Stromal cell-derived factor 1 (CXCL12) was upregulated in the thyroid of COVID-19 patients (SI Figure 8). It has been reported as a pre-B cell growth factor and would contribute to lymphopoiesis and embryogenesis homeostatic processes via regulating the migration of cells^179^. Moreover, exogenous CXCL12 would promote the migration and proliferation of human lung fibroblasts and CXCR4/CXCL12 plays an important role in IPF^180^. The upregulated CXCL12 in the thyroid may be an underlying factor for fibrosis.

Hypoxia-inducible factor-1 (HIF-1) was upregulated in the renal cortex of COVID-19 patients (SI Figure 8). It regulates genes and processes in response to hypoxia. It was upregulated in chronic kidney disease and could promote fibrogenesis^181^. The upregulated HIF1A protein in COVID-19 patients may be an early fibrosis maker.

Insulin receptor (INSR) was upregulated in the white pulp of spleen, liver and thyroid of COVID-19 patients (SI Figure 8). It binds with insulin or other ligands to activate the insulin signaling pathway. In C57BL/6 mice, when insulin receptor (*InsR*) are haploinsufficient, the mice showed impaired hepatic insulin signaling and promoted liver fibrosis accumulation, which correlated with the induction of matrix stabilization protein Lysyl oxidase like 2 (Loxl2)^182^. The upregulated INSR may indicate the activated insulin signal and a more stabilized matrix.

Integrin alpha-1(ITGA1) was downregulated in the lung of COVID-19 patients (SI Figure 8). It belongs to a subunit of integrin protein family. Together with the beta 1 subunit, ITGA1 is the receptor for laminin and collagen. Integrins are involved in communicating with the extracellular matrix, inflammatory cells, fibroblasts and parenchymal cells. Hence, they are intimately involved in the processes of initiation and maintenance and resolution of tissue fibrosis. Fibrosis models of multiple organs have demonstrated that integrins have profound effects in fibrotic process. It is now known that integrins modulate the fibrotic process through activating TGF-β^183^. Our results found a declined ITGA1 in the lung, suggesting a potential downregulated activity of integrin in the lung.

Methyl-CpG-binding protein 2 (MECP2) was downregulated in the lung and liver of COVID-19 patients (SI Figure 8). It is expressed by hepatic stellate cells (HSCs) and involves in the liver fibrosis in mice by silencing the peroxisome proliferator-activated receptor gamma (PPARgamma)^184^. However, our data found downregulation of MECP2 in the liver.

Periostin (POSTN) was downregulated in the liver and heart of COVID-19 patients (SI Figure 8). It is a secreted protein that exists in the extracellular matrix and could promote tissue remodeling^185^. Overexpression of POSTN was reported in the lungs of patients with IPF and deficiency of POSTN would protect the lung from fibrogenesis induced by bleomycin^186^. It was upregulated after stimulation by TGF-β and would promote extracellular matrix deposition and mesenchymal cell proliferation. The dysregulated POSTN may participate in the fibrosis process in COVID-19.

Protein S100-A4 (S100A4) was downregulated in the spleen of COVID-19 patients (SI Figure 8). It is a fibroblast specific protein and contains two EF-hand calcium-binding domains, belonging to the S100 family of calcium-binding proteins. It has been studied in many kinds of fibrosis^187^.

Plasminogen activator inhibitor 1 (SERPINE1) was upregulated in the lung, liver, heart renal cortex and medulla of COVID-19 patients (SI Figure 8). It inhibits the activities of urokinase-type/tissue-type plasminogen activator (uPA/PLTA), plasmin and plasmin-dependent MMP^188^, which are all involved in the proteolytic degradation. In the fibrosis tissue, SERPINE1 is significantly upregulated. Based on our analysis, elevated SERPINE1 may indicate a potential fibrinolysis inhibition in these organs and result in fibrosis.

Signal transducer and activator of transcription 3 (STAT3) was upregulated in the white pulp of spleen, liver, heart and renal cortex of COVID-19 patients (SI Figure 8). It is an important regulator of inflammation which is the primary step of fibrosis. It is activated in the inflammatory stage of tissue fibrosis and promotes fibrosis by inducing ECM^189^. Based on our analysis, overexpression of STAT3 may induce fibrosis in COVID-19 patients.

Thrombospondin 1(THBS1) was upregulated in the liver, heart and kidney of COVID-19 patients (SI Figure 8). It is a glycoprotein that is released by platelet α-granules and mediates cell-to-cell and cell-to-matrix interactions. THBS1 involves the fibrosis in TGF-β-dependent or –independent mechanisms and its antagonist could reduce the fibrosis^190^. The elevated level of THBS1 in the liver, heart and kidney may suggest potential fibrosis in COVID-19 patients.

Tyrosine 3-monooxygenase/tryptophan 5-monooxygenase activation protein beta (YWHAB) was downregulated in the white pulp of spleen, heart and renal cortex of COVID-19 patients (SI Figure 8). It is a member of the 14–3–3 protein family, which mediates signal transduction by binding to phosphoserine protein and participates in metabolism, apoptosis and cell cycle regulation^191^.

##### Stage 4: Modification

Tyrosine-protein kinase receptor UFO (AXL) was downregulated in the white pulp of spleen while it was upregulated in the thyroid of COVID-19 patients (SI Figure 9). It belongs to the Tyro3-Axl-Mer (TAM) receptor tyrosine kinase subfamily. TAM recognizes growth factors and transduces extracellular signals into the cell, participating in cell survival and proliferation, migration, and differentiation. Studies on liver fibrosis indicate that the activation of AXL and ligand Gas6 is a necessary condition for hepatic stellate cell differentiation and targeting AXL can inhibit liver fibrosis levels^192^. Studies have shown that in the model of liver injury induced by lipopolysaccharide, AXL binds to the ligand Gas6, activates macrophage autophagy, and inhibits inflammasomes^193^. These studies indicate that AXL is involved in multiple stages of the fibrosis process after liver injury. However, there was no difference in the liver of COVID-19 patients. The upregulated AXL in the thyroid tissue of COVID-19 patients may participate in the fibrosis process.

BCL2 associated X (BAX) was downregulated in the white pulp of spleen, heart, renal medulla and thyroid of COVID-19 patients (SI Figure 9). It is a member of BCL2 apoptosis regulatory protein family. It can promote the release of Cytochrome C, the activation of caspase-3, and then apoptosis. TGF-β induced inflammation and fibrosis is BAX dependent and null mutation of BAX can influence the TGF-β stimulated TIMP and MMP expression^194^.

Fibrosis is the accumulation of ECM components, or simply called collagen, in given tissues^195^. Four types of collagens were dysregulated in COVID-19 patients (SI Figure 9). Collagen alpha-1(I) chain (COL1A1) was downregulated in the heart. It was identified as a potential biomarker for heart failure progression, and its upregulation was related to the percentage of heart fibrosis^196^. The downregulation of COL1A1 may be associated with myocardial hypertrophy in control groups. Collagen alpha-1(IV) chain (COL4A1) was found upregulated in the spleen of COVID-19 patients. COL5A1 was found upregulated in the liver and heart of COVID-19 patients (SI Figure 9). COLV is a minor component of the total hepatic collagen (10–16%), but it could proliferate rapidly in active fibrogenesis in response to liver injury^197^. COLV binds TGF-β1 in the ECM, thus controlling the availability of the pro-fibrotic cytokine, which mediates fibrogenesis of neighboring cells. TGF-β1 could stimulate COL5A1 mRNA expression in mouse HSC culture, which is followed by deposition of COLV in the culture matrix with resultant COLI and III fibrillar assembly in the matrix^198^. The upregulated COL5A1 may suggest damage and fibrogenesis in the liver and heart. COL6A3 was found upregulated in the spleen and downregulated in the liver, heart and kidney of COVID-19 patients (SI Figure 9). COL6, along with its derivatives are well-known biomarkers for hepatic fibrosis^199,200^.

Cathepsin L (CTSL) was upregulated in the lung, white pulp of spleen and thyroid of COVID-19 patients (SI Figure 9). Cathepsin D (CTSD) was observed upregulated in the lung, liver, heart and thyroid of COVID-19 patients (SI Figure 9). Cathepsin is a lysosomal cysteine proteinase that participates in a role of intracellular protein catabolism and has been implicated in myofibril necrosis in myopathies and myocardial ischemia, and the renal tubular response to proteinuria^201^. CTSL was reported as a potential fibrosis marker in the liver, which would regulate the extracellular matrix degradation and tissue remodeling^202^. Hepatic stellate cells (HSCs) are responsible for extensive synthesis and deposition of ECM during liver fibrosis. CTSD has important functions outside lysosomes including degradation of ECM components when secreted to the extracellular space^203^. CTSD was observed upregulated during the activation of rat HSCs and liver fibrogenesis^204^. It was reported that CTSD can drive HSCs proliferation and promote their fibrosis potential in an *in vivo* mice model^205^. The upregulated CTSD in COVID-19 patients may indicate the fibrosis process.

Tumor necrosis factor receptor superfamily member 6 (FAS) was upregulated in the lung, white pulp of spleen and thyroid of COVID-19 patients (SI Figure 9). It is a key member of the TNF-receptor superfamily as a receptor of TNFSF6/FASLG^206^. It has been reported that it played a central role in the programmed cell death, and has been implicated in the diseases of immune system leading to inflammation^207^. The elevated FAS was known in cystic fibrosis lungs^208^ as well induced inflammation and fibrosis in the liver by Hepatitis C virus^209^. The increased FAS may indicate inflammation and fibrosis in fatal patients.

Kallikrein B (KLKB1) was downregulated in the lung, renal cortex and thyroid of COVID-19 patients (SI Figure 9). It is a plasma glycoprotein that participates in the surface-dependent activation of blood coagulation, fibrinolysis, kinin generation and inflammation. The plasma kallikrein has been reported to activate the TGF-β1 signaling by depredating the latency-associated protein and is associated with liver fibrosis in patients^210^. The declined KLKB1 may be associated with dysregulated blood coagulation.

The transcription factor farnesoid X receptor (NR1H4) was upregulated in the renal cortex of COVID-19 patients (SI Figure 9). It is ligand-activated and highly expressed in liver, kidney, intestine and adrenal glands. NR1H4 can reduce liver cell damage and fibrosis by upregulating small heterodimer companion (SHP), thereby inhibiting the production of Type I collagen and TGF-β signaling^211^. The elevated expression of NR1H4 in COVID-19 patients may participate in the fibrosis process.

Plasminogen (PLG) was downregulated in the heart and thyroid of COVID-19 patients (SI Figure 9). Its main function is to degrade fibrin clots then bind and activate on fibrin clots. The function of plasminogen activator inhibitor type-1 (PAI-1) is to regulate micro-environmental homeostasis and wound healing by inhibiting plasmin-mediated MMP activation^212^. Plasminogen is dramatically increased in adults with ARDS. It is reported that additional plasminogen may be effective in the treatment of lung lesions and hypoxemia during COVID-19 infections^213^.

Prostaglandin-endoperoxide synthase 2(PTGS2) was upregulated in the liver of COVID-19 patients (SI Figure 9). It converts arachidonate to prostaglandin H2 (PGH2), a fundamental step in prostanoid synthesis^214,215^. Upregulation of PTGS2 results in an increased level of COX-2, which potentially reduces Th2 immune responses and promotes neutrophil recruitment in hepatic ischemia/reperfusion injury^216^. The injury may be related to fibrogenesis^217^. PTGS2 is overexpressed in the bleomycin-induced pulmonary fibrosis, which was MMP-19 dependent^218^. The elevated PTGS2 may indicate that COVID-19 patients are suffering from liver fibrogenesis by the activated immune system.

Metallopeptidase inhibitor 1(TIMP1) was upregulated in the lung, liver, renal medulla and thyroid of COVID-19 patients (SI Figure 9). It is a natural inhibitor of the matrix metalloproteinases. TIMP-1 is related to the deposition of ECM, leading to fibrosis, which is elevated in the profibrotic environments^219^. While there is no difference in fibrosis severity between wild type and *Timp1*^−/-^ mice, which may be due to the inhibition of inflammation by TIMP1. The upregulated TIMP1 may be due to the fibrosis generation in these organs.

## Multi-organs proteomic changes (supple for Figures 6 and 7)

### Lung and spleen

The lung is the primary and the only positive organ of virus infection (Table S1). To investigate the virus induced pathological features, we first focused on the lung proteome. The most significant pathologic injury in the lung of COVID-19 patients is fibrosis, accompanying with fibrosis associated fibrous exudation, hyaline membrane formation and alveolar space collapsed^220^ (Figure S1).

As discussed above (Cluster1: Receptor), the expression of ACE2, the known receptor of SARS-CoV-2, and the expression of CD209 and CLEC4M, two receptors of SARS, shown no significant changes in COVID-19 patients (Figure 2C). However, a couple of proteins acting as the cellular entry of other viruses, such as CTSL, CEACAM1 and NPC1, were found upregulated in the COVID-19 patients (Figure 2C).

During virus replication, double-stranded RNA (dsRNA) and uncapped mRNA of virus, known as viral pathogen-associated molecular patterns (PAMPs), could trigger the innate immune response once recognized by the pattern recognition receptors (PRRs) in the cytoplasm^221^. According to the PRRs list^221^, we found CGAS was upregulated in the lung of COVID-19 patients (Figure 5A, SI Figure 10a). The upregulation of CGAS further induces downstream response including Type I interferon secretion and the release of several other cytokines^222^.

During the transcription and replication of coronavirus, host gene expression could be blocked by mRNA suppression and translation shutoff. For example, the N protein has been adapted to protect viral mRNA from degradation^223^. The lung is the only organ where SARS-CoV-2 viral RNA was tested to be positive in our study. Therefore, we compared the virus associated pathways in the lung with the ones in other organs (Figure 4C). The trend of EIF2 signaling and tRNA charging showed a similar trend in the lung and liver (Figure 4A). We found an inhibition of ARE-mediated mRNA degradation pathway in the lung and an activation trend in all the other organs (Figure 4C). Eukaryotic translation initiation factor 4E (EIF4E) as the cap-binding protein in the host cell only increased in the lung of COVID-19 patients (Figure 5A, SI Figure 10a), which could be a potential target for human coronavirus 229E (HCoV-229E)^224^.

Once virus infected the host, viral pathogen-associated molecular patterns (PAMPs), such as dsRNA, would trigger host immune cells for defense. The major pathology in immune is hyper inflammation, such as C-reactive protein (CRP) increasing and lymphopenia in the peripheral blood and spleen of these patients (Figures S1,2). The white pulp of spleen consists of T and B lymphocytes while the marginal zone of white pulp and red pulp contains the APCs. Pathologically, we observed that white pulp was atrophic while no pathological changes were observed in the red pulp (Figure 1), the latter of which is consistent with our proteome data (Table S4).

In the lung of COVID-19 patients, high affinity immunoglobulin gamma Fc receptor Ⅰ (FCGR1A) and low affinity immunoglobulin gamma Fc receptor Ⅱ-a (FCGR2A) was upregulated (Figure 5A). While in the spleen, high affinity immunoglobulin epsilon receptor subunit gamma (FCER1G), FCGR1A and FCGR2A were all upregulated (Figure 5A). These receptors expressed on the monocytes surface for antigen presentation. TNF receptor-associated factor 6 (TRAF6) was upregulated in the spleen of COVID-19 patients, which plays a role in dendritic cell maturation and stimulation of naïve T cell^225^. TYRO protein tyrosine kinase-binding protein (TYROBP) was upregulated in the lung and spleen of COVID-19 patients, which is involved in NK cell anti-viral function and inflammatory reactions^226^. All these proteins participating innate cell activity indicate the activated antigen presenting process or anti-viral functions.

The activation of NK cells might also be negative feedback for the decreasing T-, B- lymphocytes. Consistently, the adaptive immune associated pathways including B cell receptors signaling, NFAT in regulation of the immune response were all inhibited (SI Figure 10b). Tyrosine-protein kinase (LCK), which actives CD8 T cell combined with LR2R^227^, was downregulated (Figure 5A). CEACAM1 as a negative regulator of adaptive immune also upregulated (Figure 5A) which indicated the immunosuppression state in the COVID-19 patients and might strengthen the proinflammatory cytokine secreted.

Except for the above proteins, a well-known immunosuppression molecular, CD274 (PD-L1), is upregulated significantly (Figure 5A) and PD-1, PD-L1 pathway was activated (SI figure 10c) in the spleen of COVID-19 patients. When mapping our data with an immune checkpoint database including 21 immune checkpoint proteins^228^, we found three dysregulated proteins, including CEACAM1, CD276 and CD274 (Figure 5A). Upregulation CEACAM1 and CD276 appeared in the lung and upregulation of CD274 was in the spleen. The upregulation of these checkpoint proteins may indicate the suppression of adaptive immune function.

The proinflammatory state including the excess cytokines would induce tissue injury. M2 macrophages mainly participate in the wound healing and tissue repair process. Sterol 26-hydroxylase (CYP27A1) was upregulated in the COVID-19 patients (Figure 5A), which promotes M2 conversion through increasing 27HC synthesis^229^. In addition, the maker of M2, CD163, was upregulated in six organs (except testis), indicating that the wound healing and tissue repair have been induced in the COVID-19 patients. Due to the excess export of lymphocytes to other injury organs and increased apoptosis, exhausted lymphocytes and the white pulp may atrophy^230^.

In summary, these dysregulated proteins and enriched pathways suggest that the innate immune was activated, the APCs were activated and increased with simultaneous wound healing, while the adaptive immune was inhibited, which indicates the hyper inflammation state in the COVID-19. To further validate the immune state in COVID-19, we performed the immunohistochemical (IHC) staining for immune cells in the spleen and found that total T cell (CD3), T helper cells (CD4), cytotoxicity T cells (CD8), and B cells (CD20) all decreased in the spleen (Figure S6). On the contrary, macrophage (CD68), especially M2 (CD163) all increased (Figure S6). These hyper inflammation state might further induce tissue over-repair and fibrosis.

### Liver

In the COVID-19 group, the main pathological changes in the liver are the steatosis and coagulative necrosis (Figure S1). Pathway analysis of 1,969 significant dysregulated proteins in the liver shown the acute phase response signaling, eIF2 signaling, fatty acid α and β-oxidation, necroptosis signaling pathway, apoptosis signaling and stearate biosynthesis were activated while the xenobiotic metabolism was inhibited (SI Figure 11a). Several key proteins related to necroptosis were shown in SI Figure 11b. The steatosis and necrosis processes are always accompanied by inflammation^231,232^. Thereby, we mapped our differentially expressed proteins in the liver with the immunological proteins from GSEA-immunologic gene sets and then employed IPA for pathway analysis for the overlapping proteins (SI Figure 11c). The key proteins participating in these pathways are shown in Figure 5B. The proinflammatory state in the liver was reflected with activated IL-6, IL-8 signaling and NF-κB signaling. Remarkably, we found that hepatic fibrosis signaling (SI Figure 11c) was activated and hepatic fibrosis markers including metalloproteinase inhibitor 1 (TIMP1) and plasminogen activator inhibitor 1 (SERPINE1)^233^ were upregulated (SI Figure 11b). These dysregulated proteins might indicate that the chronic inflammation and fibrosis process have initiated at molecular level though no obvious pathological changes were observed. This phenomenon might be clinically meaningful for patients who have recovered to take special care for the potential to develop fibrosis.

We hence propose a hypothetical model in the liver of COVID-19 patients according to the main pathological changes and the enriched pathways (SI Figure 11d). Firstly, the inflammation biomarkers, such as C reactive protein (CRP), serum ferritin, IL-6 were significantly elevated in COVID-19 patients (Figure S2), which might be stimulated by the inflammatory response derived from the lung through cytokines and immune cells. The upstream regulator, STAT3, and pathway analysis, NF-κB signaling pathways were enriched, which were induced by cycling IL-1, IL-6 and IL-8. As a result, the acute phase response was activated and the expression of CRP upregulated^234^. Due to the activated NF-κB signaling, the two related catalytic subunits of IKK kinase complex and its regulatory subunit, IKBKG, NEMO and IKKG, were all upregulated. The IKK kinase complex induced the degradation of IκB inhibitory molecules and lead to the nuclear translocation of the NF-κB dimers, which would activate transcription of genes participating in the immune and inflammatory response^235^. Besides, the NF-κB could also activate IL-8 signaling and TNF signaling, which may contribute to hepatic fibrosis process^236,237^.

Secondly, among the significantly dysregulated transcription factors (Table S5), we noticed the downregulated hepatocyte nuclear factor 4-alpha (HNF4A) (SI Figure 11b), which regulates lipid homeostasis in the liver^238^. The hypoxia state and upregulated STAT3 would reduce the expression of HNF4A^239,240^. Following its downregulation, very low-density lipoprotein (VLDL) mediated lipids exporting was reduced by downregulated microsomal triglyceride transfer protein large subunit (MTTP) and apolipoprotein B-100 (ApoB). Meanwhile, the hepatic uptake of fatty acids was enhanced by upregulated scavenger receptor class B member 1 (SCARB1) (SI Figure 11b). Both reduced lipids exporting and enhanced fatty acids uptake would contribute to the hepatic steatosis^241^.

The hepatic steatosis is always associated with insulin resistance, which may be induced by excess ROS leakage by upregulated fatty acid oxidation and mitochondrial dysfunction^242^. In COVID-19 patients, the enzymes and transporters involved in fatty acid oxidation, such as ACSL3, CPT2, SLC27A4, TMLHE, ECI1, HADHA/B, IVD and SCP2, were significantly upregulated, leading to the generation of reduced NADH. Moreover, the upregulated ATP synthase (ATP5F1A/B/C/D/E, ATP5MC1/E/F/B/D/F/O), NADH dehydrogenase and its oxidase involving oxidative phosphorylation catalyze more energy production, which is benefit for the translation of acute phase proteins and coagulation factors. Besides, it would lead to the excess generation of reactive oxygen species (ROS), which induce necrosis^243^ and insulin resistance. Moreover, the upregulated voltage-dependent anion-selective channel proteins (VDAC1/2/3), mitochondrial import receptor (TOMM22/40/40L/5/70) and mitochondrial import inner membrane translocase (TIMM9) induced the mitochondrial membrane permeabilization^244^, could all lead to the release of proapoptotic proteins such as DIABLO, AIFM1 and CYCS and activate apoptosis^243^.

Thirdly, the xenobiotic metabolism-PXR signaling pathway was downregulated in the liver of COVID-19 patients. The downregulation of its related enzymes, such as Cytochrome P450 2C9 (CYP2C9), Glutathione S-transferase A1 (GSTA1), UDP-glucuronosyltransferase 2B7 (UGT2B7), S-formylglutathione hydrolase (ESD) and Sulfotransferase 1E1 (SULT1E1), could be reasoned by antibiotics, antivirals and steroids treatment, the widely used therapies of COVID-19^245^.

In summary, we speculate the pathological damage in the liver of COVID-19 patients is mainly caused by the severe inflammatory response and the hypoxia status by the viral infection in the lung, as well as the drug-induced hepatotoxicity during therapy (SI Figure 11d).

### Heart

Two kinds of pathological changes were observed in the heart of COVID-19 patients. One is vasogenic edema with mild infiltration of inflammatory cells, and the other is myocardial cell edema (Figure S1).

Vasogenic edema is mainly caused by the change of vascular permeability, which induces the serum fluids to flow into interstitium. The increase of vascular permeability is usually caused by inflammation^246,247^. Several inflammatory mediators were elevated in the heart tissue of COVID-19 patients, including matrix metalloproteinases 9 (MMP9), C5a anaphylatoxin chemotactic receptor 1(C5AR1), interferon gamma receptor 1 (IFNGR1) and several other cytokines (Figure S7A, SI Figure 12a). Importantly, histamine N-methyltransferase (HNMT), which catabolizes histamine, was significantly downregulated and may lead to histamine accumulation and vascular hyperpermeability^247^. The accumulation of inflammatory mediators, such as C5a and histamine, could induce the P-selectin expression and result in leukocyte extravasation^248^ (SI Figure 12b). The above findings were also supported by the upregulation of intercellular adhesion molecule 1 (ICAM-1), which involves in leukocyte adhesion and trans-endothelial migration^249^ and MMP9, which facilitates matrix degradation and is believed to contributes to inflammatory cell invasion^250^. Physiologically consistent, we observed inflammatory cell infiltration by hematoxylin-eosin staining sections (Figure S1).

Myocardial cell edema is caused by the movement of osmotically active molecules from the extracellular to the intracellular space including sodium, chloride and water. One of the underlying pathological mechanisms was the reduced ATP availability in the mitochondria due to dysfunction of sodium (Na^+^), potassium (K^+^) ATPase^251^ under hypoxia, inflammation and other conditions^252^. These observations strongly indicated a hypoxia situation in COVID-19 patients, and we also observed consistent molecular changes under hypoxia (SI Figure 12 c). Under aerobic conditions, the hypoxia-inducible factor 1-alpha (HIF-1α) can be ubiquitinated by ubiquitin-activating enzyme (E1), ubiquitin-conjugating enzymes E2 (E2), and a substrate-specific ubiquitin-protein ligase (E3) and then degraded by hypoxia-inducible factor 1-alpha inhibitor (HIF1AN)^253^. In COVID-19 patients, we identified a decreased expression of HIF1AN, ubiquitin-conjugating enzyme E2 G1 (UBE2G1) and ubiquitin-conjugating enzyme E2 H (UBE2H) (Fiugre S7A), suggesting the stabilization of HIF-1α under hypoxic conditions. Besides, the heme protein myoglobin (Mb) specializing in oxygen transport and storage was downregulated. These observations strongly indicated a hypoxia situation in COVID-19 patients. What’s more, thioredoxin-2 (TXN2), a redox protein, which is essential for mitochondrial reactive oxygen species (ROS) homeostasis, is downregulated in our analysis. It is reported that TXN2 deficiency could lead to increased reactive oxygen species (ROS) levels and impaired mitochondrial function^254,255^.

In conclusion, the molecular changes in the heart of COVID-19 patients were consistent with the HE staining result of vasogenic edema and cytotoxic edema. It’s worth noting that the control samples were collected from dilated cardiomyopathy (DCM) patients. Therefore, the molecular changes involved in DCM patients, such as pathways related to ATP generation^256^ should be considered carefully.

### Kidney

All COVID-19 patients in our study showed a varying degree of acute kidney injury (AKI) (Table S1). Diffuse proximal tubule injury, brush border lost and microthrombus were observed in the pathological sections of the cortex, while occasional cellular swelling and atrophy were also observed in the medulla (Figure S1). Generally, more damage was observed in pathological sections in the renal cortex than in the renal medulla.

Notably, according to the 2016 Third International Consensus Definition for Sepsis and Septic Shock^257^ (Table S1), sepsis was observed in the most COVID-19 patients as a complication (Table S1), with a decrease of platelet, an increase of D-dimer and neutrophil (Figure S2, Table S1). At the molecular level, we identified a total of 9,544 proteins in the kidney with 1,585 dysregulated proteins in the renal cortex and 642 in the renal medulla (Figure 1). By IPA analysis, the LPS/IL-1 mediated inhibition of RXR function, acute phase response, IL-8 signaling, necroptosis and neuroinflammation signaling pathways were found to be upregulated in both cortex and medulla (SI Figure 13a). Meanwhile, the activated status of the enriched pathways in the renal cortex are more drastic than in the renal medulla (SI Figure 13a). Besides, the upregulated pathways including signaling by Rho Family GTPase, Toll−like receptor signaling, IL-6 signaling and NF –κB signaling and the downregulation pathways including fatty acid beta−oxidation I, glycolysis and gluconeogenesis were also specifically enriched in the renal cortex. Thus, at both pathological and molecular levels, SARS-COV-2 has a greater impact on the cortex than the medulla (SI Figure 13a, Figure S1).

In COVID-19 patients, lipopolysaccharide-binding protein (LBP), interleukin-1 receptor 1 (IL1R1) and vWF were upregulated in renal cortex (Figure S2). LBP could bind to the pathogen lipopolysaccharides (LPS) and enhance CD14 and Toll-like receptors, which may be the cause of pathogenic infection and sepsis in these COVID-19 patients^258^. Another inflammatory cytokine, Interleukin-1 (IL1) could also induce sepsis^259^. The IL1R1 was found to be upregulated in COVID-19 patients, as well as sepsis^259,260^. Upregulation of the downstream NF-κB complexes and CCAAT enhancer binding protein beta (CEBPB) were also observed, leading to activation of IL-6, IL-8, iNOS and TNFR1 signaling pathways^261^. Besides, activated signal transducer and activator of transcription 1 (STAT1) and receptor interacting serine/threonine kinase 1 (RIPK1) could play a role in apoptosis and tissue damage^262–264^.

In addition to inflammatory and infection-related proteins, we also detected the upregulation of hypoxia inducible factor 1 subunit alpha (HIF1A) in the cortex, which could promote TRL2, vWF overexpression and nitric oxide (NO) generation. These could lead to insulin resistance^265^. Furthermore, HIF1A could suppress energy metabolism-related pathways including fatty acid β-oxidation and gluconeogenesis to exacerbate insulin resistance^266^. As the main ATP provider in the renal proximal tubular cells, the inhibition of fatty acid β-oxidation would block energy supply and lead to aggravate kidney injury^266^. Interestingly, among seven organs, kidney is the only one in which ACE2 was downregulated (Figure 2C), ACE2 not only acts as the receptor of SARS-CoV-2 but could also catalyze the cleavage of angiotensin II (Ang II) into vasodilator angiotensin 1–7 (Ang 1–7). The downregulation of ACE2 could mediate Ang II accumulation and induce the upregulation of vascular cell adhesion molecule 1 (VCAM1)^267^, a marker for sepsis and kidney injury^268,269^. Likewise, Ang II regulates RhoGTPase via G Protein-Coupled Receptor (GPCR), indeed the Gα, Gβ and RhoGTPase including RHO and CDC42 were all upregulated in COVID-19 patients. ROCK1, a downstream molecular of RHO, was upregulated as well, which could lead to renal tubular injury, inflammation and fibrogenesis^270,271^. CDC42 induced CDC42EP4 is an upstream regulator of SEPTIN family and the upregulated SEPTIN6, SEPTIN7, SEPTIN8 and SEPTIN9 could induce fibrogenesis in kidney as well, which means SARS-CoV2 infection may cause chronic injury and affect prognosis in COVID-19 patients^272^. Meanwhile, the use of non-steroidal anti-inflammatory drugs and β-lactam antibiotics are also possible precipitating factors of kidney injury^273^ (Table S1).

In conclusion, renal tissue damage is more severe in renal cortex than in the medulla of COVID-19 patients, and the AKI may be caused by the combinational effects of hypoxia, bacterial infection, sepsis induced cytokine storm and drug-induced nephrotoxicity.

### Thyroid

Lymphoid infiltration was found in some interfollicular regions in the thyroid of COVID-19. In our study, 1,297 proteins in the thyroid were dysregulated ((Figure 1) and the top ten pathways are enriched for these proteins (SI Figure 14a).

Among these enriched pathways, eIF2 and mTOR upregulation might be associated with protein synthesis and indicate the dysregulation of thyroid function at molecular level. IL-8 and NF-κB signaling are associated with inflammatory response physically. The pathology associated pathways including angiogenesis (VEGF signaling), coagulation and thrombin signaling, cardiac hypertrophy signaling and renin-angiotensin signaling were also enriched (SI Figure 14A). The key dysregulated proteins in these pathways are listed in the Figure S7D. We proposed a hypothetical model for thyroid pathology by these key proteins and pathways (SI Figure 14B). The thyroid is adjacent to the superior respiratory system, thereby would be easily influenced by the inflammation. Some cytokines and their receptors, such as CXCL12, IL1R1 and IL6ST, were dysregulated, which could stimulate the NF-κB or mTOR signaling and then induce a series of downstream cascade reactions. CXCL12 is involved in promoting cell growth and angiogenesis^274^, which is associated with mTOR signaling^275^. In thyroid of COVID-19 patients, mTOR and CXCL12 were both upregulated (Figure S7D). IL-1 and IL-6 are associated with a variety of chronic inflammatory response, such as coagulation^276^ and insulin resistance^277^, through activating NF-κB signaling.

In the thyroid of COVID-19 patients, Cell division control protein 42 (CDC42) and G protein subunit alpha Q (GNAQ) were both upregulated. CDC42 belongs to GTPase family involved in the thrombin formulation process^278^ and GNAQ also belongs to G proteins and is involved in thrombin signaling^279^. The upregulated CDC42 and GNAQ may indicate the coagulation response in the thyroid. Besides, two kinds of coagulation associated protease inhibitors, serine protease inhibitors (SERPINs) family^280^ and alpha-2-macroglobulin (A2M), were all downregulated in the thyroid of COVID-19 patients (Figure S7D), which could suppress plasmin and kallikrein^53^. Kininogen-1 (KNG1), an alpha-2-thiol protease inhibitor, was also downregulated in the thyroid of COVID-19 patients (Figure S7D), which could disturb kinin-kallikrein system^281^.

Neuroendocrine convertase 2 (PCSK2), insulin receptor (INSR) and carboxypeptidase E (CPE) all participate the insulin processing and their secretion is regulated by mTOR in insulin resistance^282^, which also matched with our data (Figure S7D).

Altogether, the thyroid is mainly affected by the inflammatory response after virus infection, in which the activated NF-κB and mTOR signaling could have an association with coagulation, insulin resistance and VEGF involved angiogenesis.

### Testis

Because of one of the dysregulated proteins in the testis do not have GO annotation, nine differential expressed proteins in the testis are discussed in this part (SI Figure 15). We performed a functional enrichment of the 9 downregulated proteins by STRING, and found that 5 out of 9 proteins are enriched in cholesterol biosynthetic process (GO:0006695, FDR = 9.19e-07) and cholesterol metabolic regulation (GO:0090181, FDR = 9.19e-07), including Squalene synthase (FDFT1), ATP-citrate lyase (ACLY), squalene monooxygenase (SQLE), FASN, and Delta(14)- sterol reductase TM7SF2 (TM7SF2).

Although those proteins are unrelated to the virus directly, the cholesterol synthesis is associated with steroid hormones such as testosterone. The low intracellular concentration of sterols would induce the cleavage of membrane bound sterol response element-binding protein (SREBP) by SREBP cleavage-activation protein (SCAP), then cleaved SREBP in the cytosol would enter the nucleus to activate expression of proteins participated in cholesterol biosynthesis and uptake process^283^.

Delta (14)-sterol reductase is encoded by *TM7SF2*, which can be activated in response to low cell sterol levels mediated by SREBP2^284^. TM7SF2 is detected in Leydig cells specifically by antibody staining^285^, the decrease of which suggested impaired Leydig cell population or function. We also found a Leydig cell biomarker, INSL3, was decreased (SI Figure 15). INSL3 was first isolated from the boar testis cDNA library and is expressed uniquely in fetal and postnatal Leydig cell^286^. INSL3 is mainly produced in gonadal tissues in male and female^287^ and is known to be associated with testicular descent^288,289^. Three independent studies reported the association of INSL3 gene mutation with human cryptorchidism, which is a common congenital abnormality in testis^290–292^. INSL3 seems to function within the testis as an important supplementary downstream effector of the hypothalamic–pituitary–gonadal (HPG) axis^287^. Free from acute regulation of HPG axis, it can reflect the differentiation status and the number of the Leydig cells present^293^. The unique INSL3 receptor, RXFP2, has been identified at mRNA and protein levels on both Leydig cells themselves^294^ and also on germ cells within the seminiferous compartment^295^. The most significantly decreased proteins in the testis is INSL3, which indicates Leydig cell reduction and might associates with diabetes^296^ as well.

The hypothalamic–pituitary–gonadal (HPG) axis comprises of pulsatile GnRH from the hypothalamus. It will impact on the anterior pituitary to induce expression and release of both LH and FSH into the circulation. These will, in turn, stimulate receptors on testicular Leydig and Sertoli cells, respectively, to promote steroidogenesis and spermatogenesis. Both Leydig and Sertoli cells exhibit negative feedback to the pituitary and/or hypothalamus via their products testosterone and inhibin B, respectively, thereby allowing tight regulation of the HPG axis. In particular, LH exerts both acute control on Leydig cells by influencing steroidogenic enzyme activity, as well as chronic control by impacting on Leydig cell differentiation and gene expression^287^.

Significantly decreased INSL3 indicates the decreased number of mature Leydig cells in the patients with COVID-19, which is consistent with the HE staining images (Figure 7B, C). Interestingly, Ma et al observed significantly elevated serum luteinizing hormone (LH) and dramatically decrease in the ratio of testosterone (T) to LH and the ratio of follicle stimulating hormone (FSH) to LH, while no statistical difference in serum T (p = 0.0945) or FSH

(p = 0.5783)^297^. We inferred the change on LH might be caused by the reduced number of mature Leydig cells, which would cause the decreased serum T. Weakened negative feedback from Leydig cells might promote the secretion of LH.

FASN, ACLY, SQLE and FDFT1 all participate in lipid synthesis directly. Fatty acid synthase is a key enzyme in de novo lipogenesis. FASN is markedly inactivated under conditions of insulin resistance and is a potential biomarker for insulin resistance^298^. Reduced FASN suggested the progression of insulin resistance in COVID-19 patients. This protein has been discussed in the fibrosis process associated proteins.

ACLY is the enzyme primarily responsible for the production of extramitochondrial acetyl-CoA and is thus strategically positioned at the intersection of glycolytic and lipidic metabolism^299^, which make it a potential target for lipid lowing^300^. FDFT1 and SQLE play an important role in the cholesterol biosynthesis pathway, and both of them are effective targets in the treatment of hypercholesterolemia^301,302^. ACLY, SQLE and FDFT1 were observed downregulated in the testis tissue of COVID-19 patients compared with non-COVID-19 controls (SI Figure 15), which was consistent with a previous study discovering lower levels of low-density lipoprotein cholesterol and total cholesterol levels in COVID-19 patients as compared with normal subjects^303^.

There are two dysregulated proteins associated with sperm generation. RNF216 is essential for spermatogenesis and male fertility^304^. The deletion of *Drc7* leads to aberrant tail formation in mouse spermatozoa that phenocopies patients with multiple morphological abnormalities of the sperm flagella (MMAF)^305^. These two proteins were reduced in COVID-19 patients (SI Figure 15), suggesting the impairment of spermatogenesis and sperm motility caused by SARS-CoV-2 infection.

Store-operated calcium entry-associated regulatory factor (SARAF) was downregulated in the testis of COVID-19 patients (SI Figure 15), which inactivates the store operated calcium entry machinery to prevent excess calcium refilling^306^.

In conclusion, reduced INSL3 and five cholesterol biosynthesis related proteins suggests damage to Leydig cells and their steroidogenic functions (Figure 7D). Decrease of DRC7 indicates impaired sperm mobility. Future investigations of Leydig cell functions and sperm mobility of alive COVID-19 patients are needed since our observations are based on autopsies.

**SI Figure 1.**
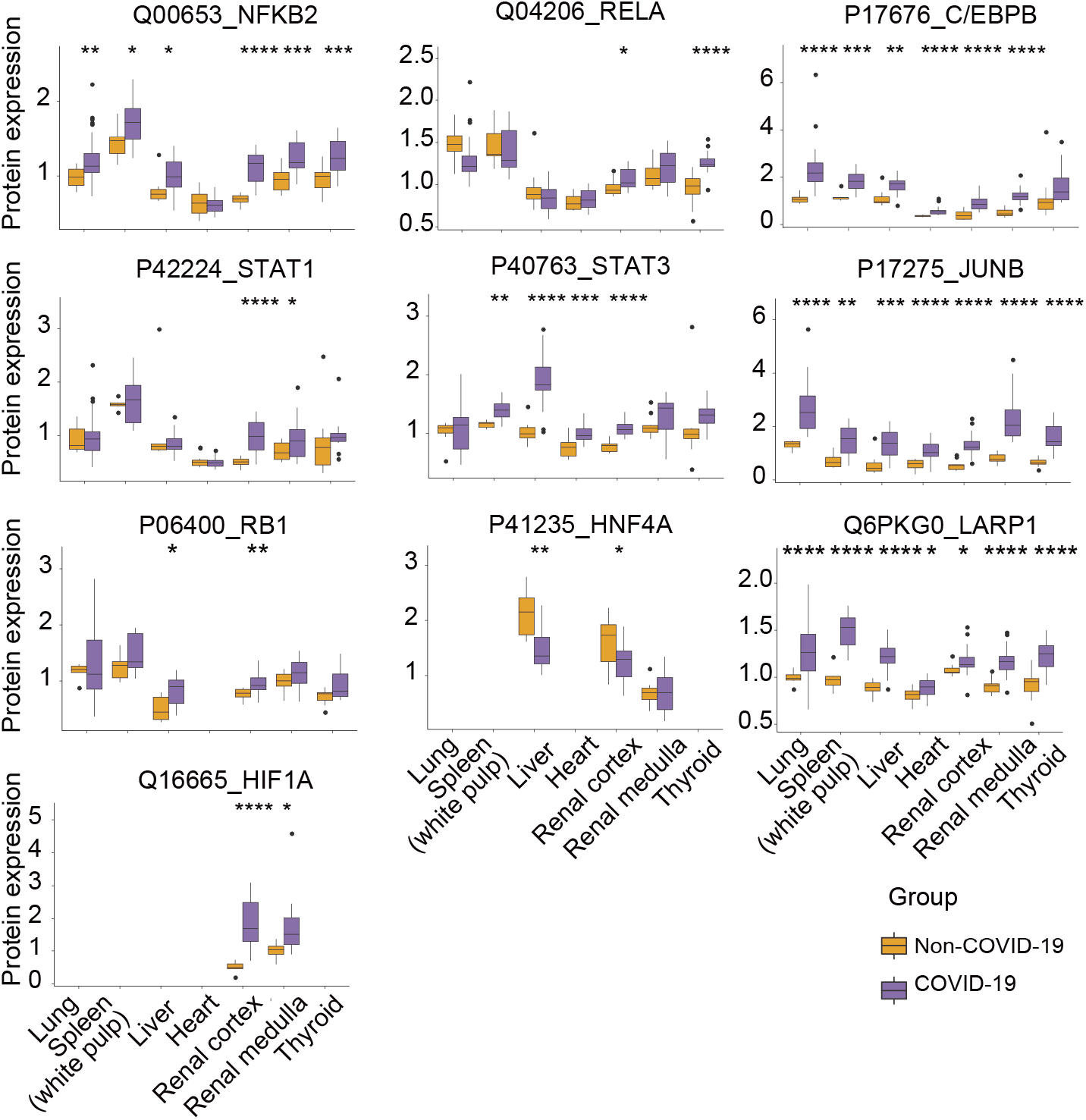
Protein expression of transcription factors with same fold-change trend in experiential FC and the predicted activation state by IPA. The y-axis stands for the protein expression ratio by TMT-based quantitative proteomics. Pair-wise comparison of each protein between COVID-19 patients and control groups was performed with student’s *t* test. *, p < 0.05; **, p < 0.01; ***, p < 0.001; ****, p < 0.0001.

**SI Figure 2.**
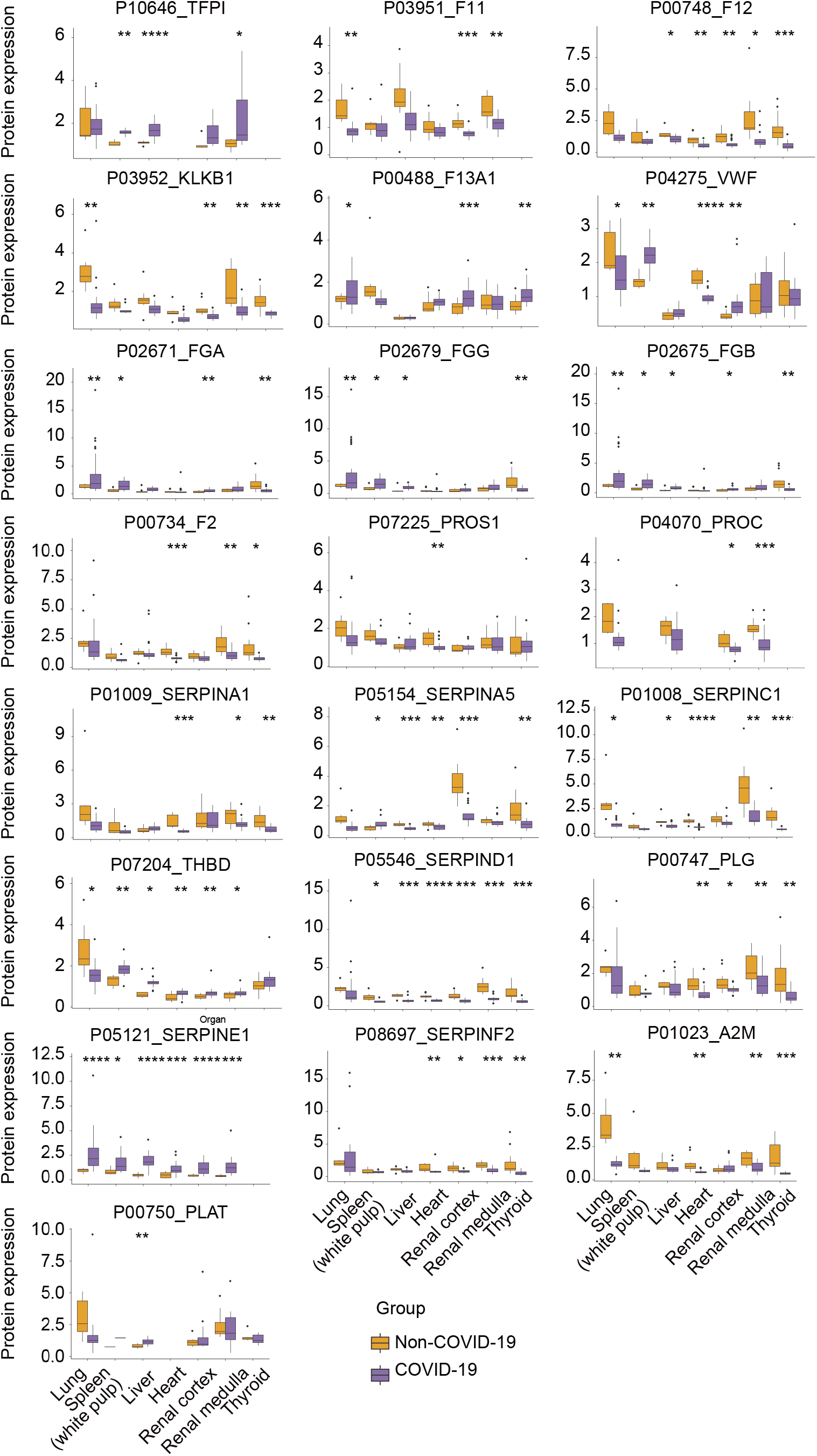
Expression of dysregulated proteins in coagulation system. The y-axis stands for the protein expression ratio by TMT-based quantitative proteomics. Pair-wise comparison of each protein between COVID-19 patients and control groups was performed with student’s t test. *, p < 0.05; **, p < 0.01; ***, p < 0.001; ****, p < 0.0001.

**SI Figure 3.**
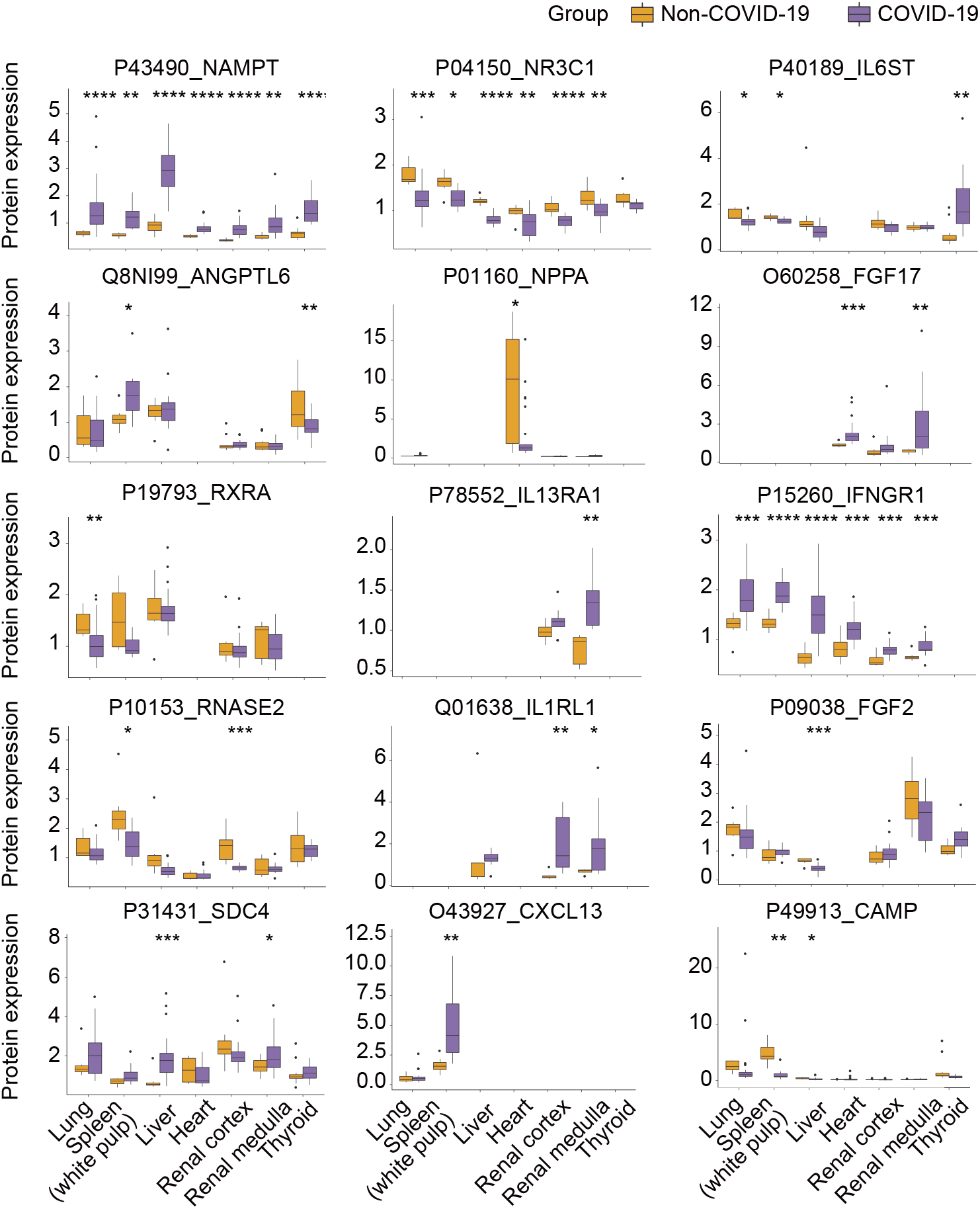
Protein expression of dysregulated cytokines. The y-axis stands for the protein expression ratio by TMT-based quantitative proteomics. Pair-wise comparison of each protein between COVID-19 patients and control groups was performed with Student’s T test. *, p < 0.05; **, p < 0.01; ***, p < 0.001; ****, p < 0.0001.

**SI Figure 4.**
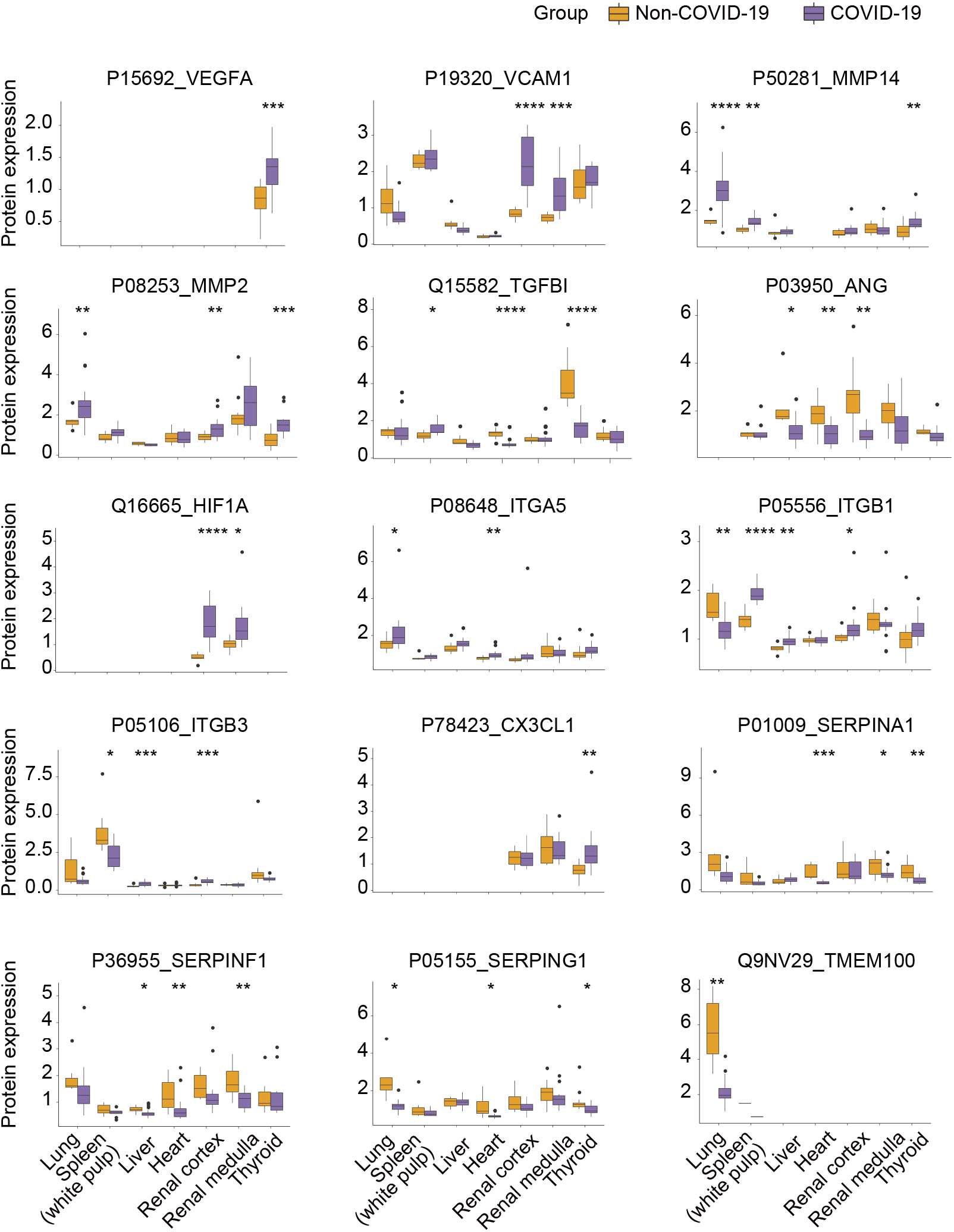
Expression of dysregulated proteins involved in angiogenesis. The y-axis stands for the protein expression ratio by TMT-based quantitative proteomics. Pair-wise comparison of each protein between COVID-19 patients and control groups was performed with Student’s T test. *, p < 0.05; **, p < 0.01; ***, p < 0.001; ****, p < 0.0001.

**SI Figure 5.**
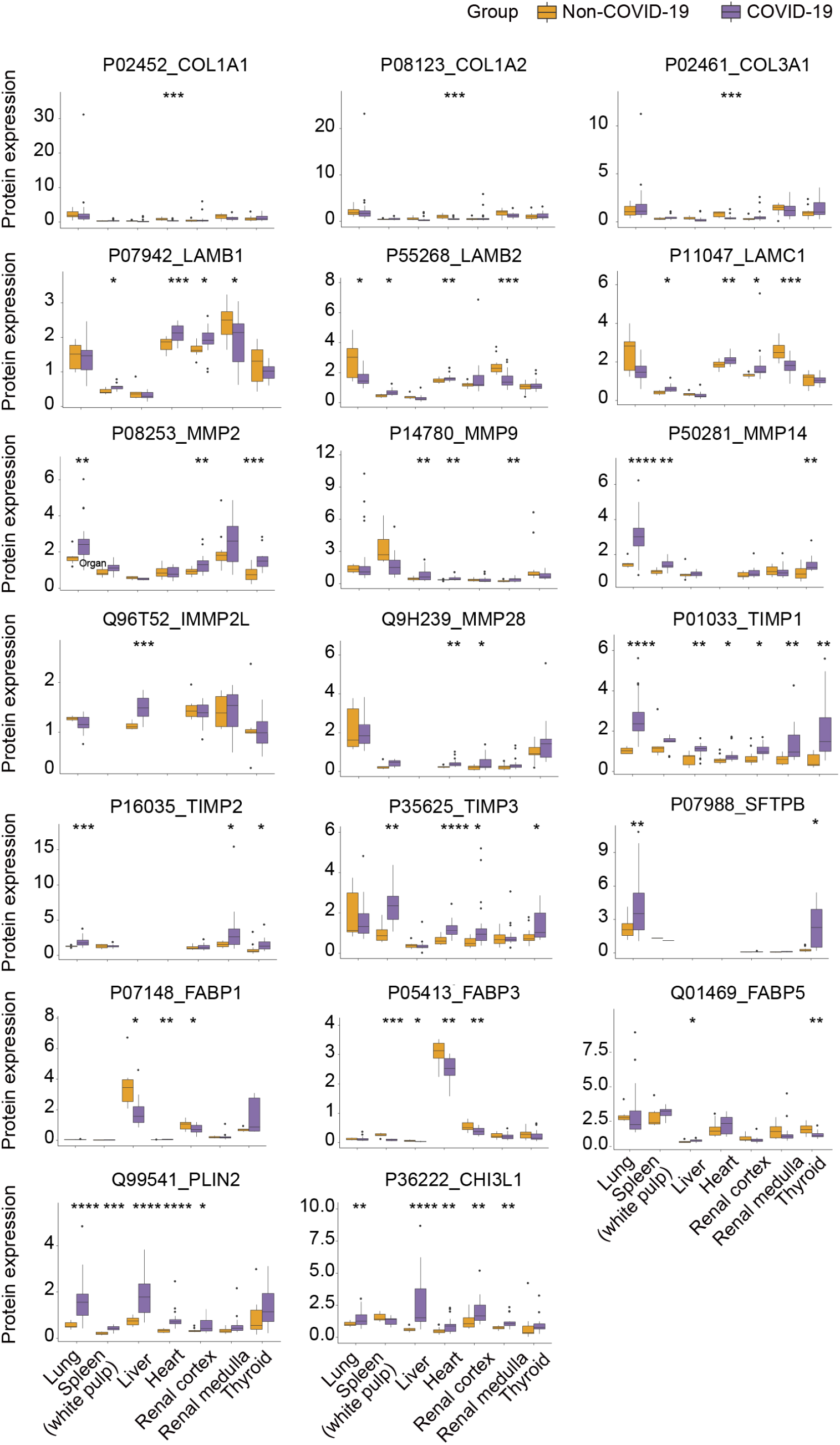
Expression of fibrosis markers dysregulated between COVID-19 patients and the control group. The y-axis stands for the protein expression ratio by TMT-based quantitative proteomics. Pair-wise comparison of each protein between COVID-19 patients and control groups was performed with Student’s t Test. *, p < 0.05; **, p < 0.01; ***, p < 0.001; ****, p < 0.0001.

**SI Figure 6.**
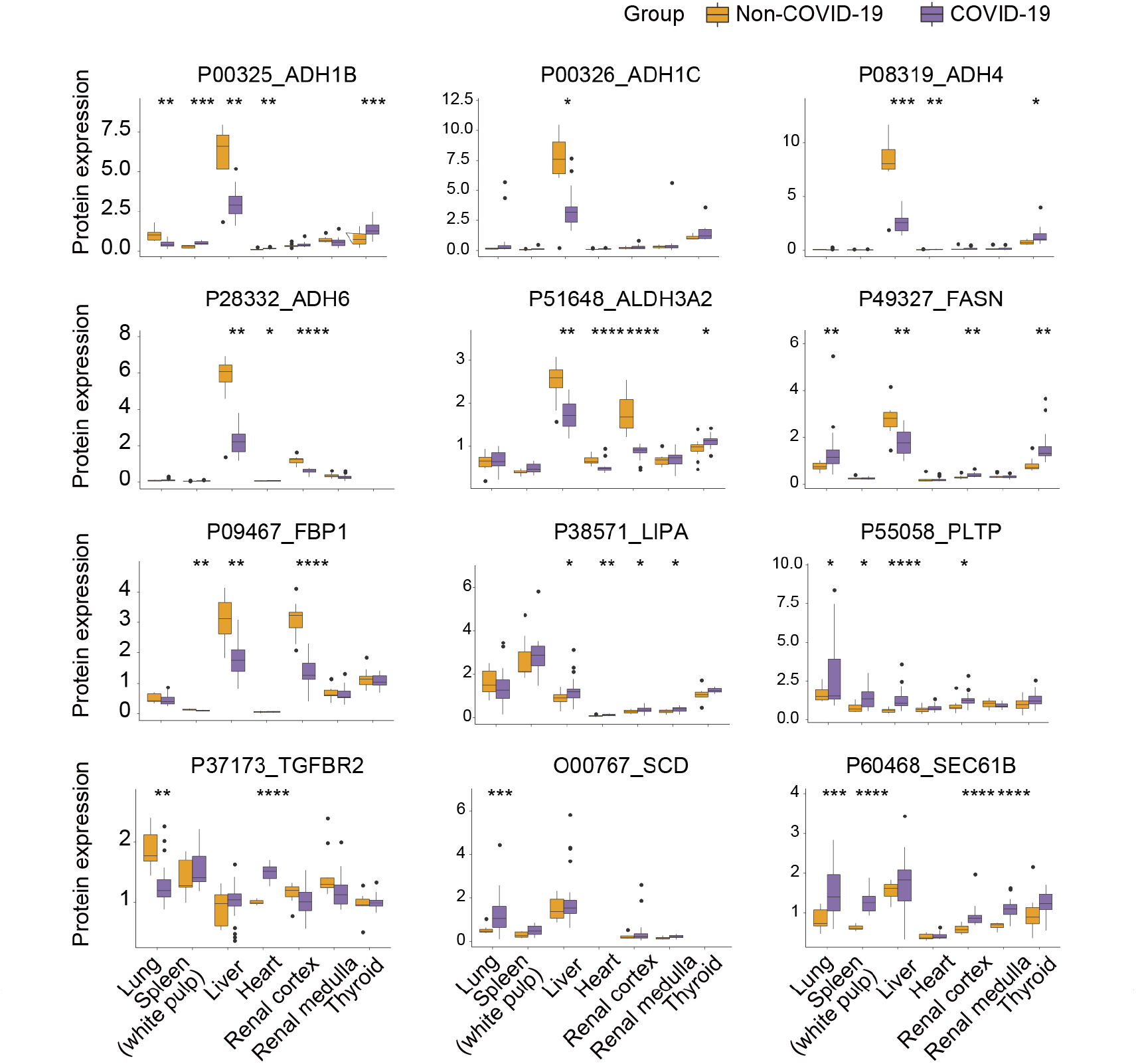
Expression of dysregulated proteins involved in initiation of fibrosis process. The y-axis stands for the protein expression ratio by TMT-based quantitative proteomics. Pair-wise comparison of each protein between COVID-19 patients and control groups was performed with student’s t test. *, p < 0.05; **, p < 0.01; ***, p < 0.001; ****, p < 0.0001.

**SI Figure 7.**
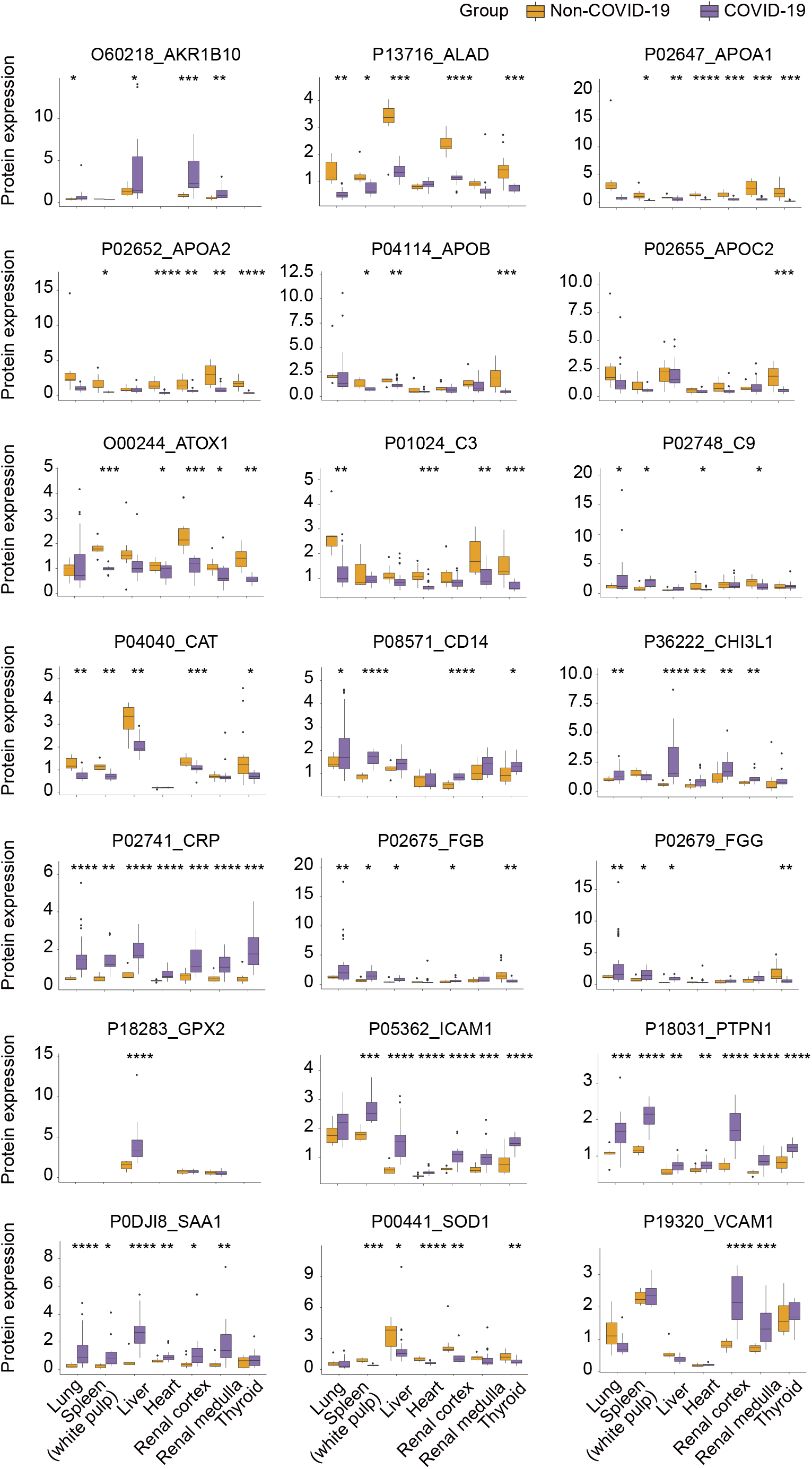
Expression of dysregulated proteins involved in inflammation of fibrosis process. The y-axis stands for the protein expression ratio by TMT-based quantitative proteomics. Pair-wise comparison of each protein between COVID-19 patients and control groups was performed with student’s t test. *, p < 0.05; **, p < 0.01; ***, p < 0.001; ****, p < 0.0001.

**SI Figure 8.**
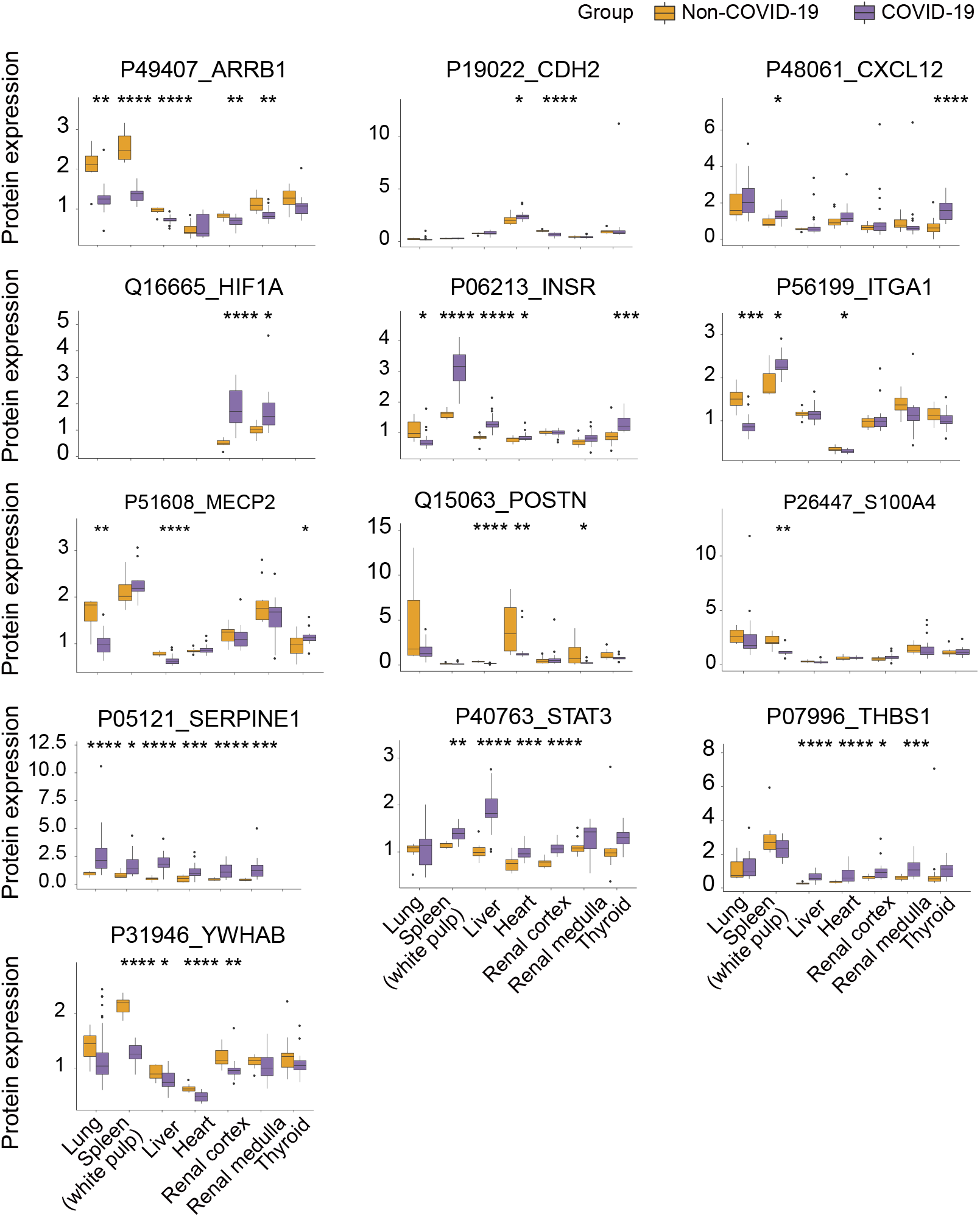
Protein expression of dysregulated proteins involving in proliferation of fibrosis process. The y-axis stands for the protein expression ratio by TMT-based quantitative proteomics. Pair-wise comparison of each protein between COVID-19 patients and control groups was performed with Student’s T-test. *, p < 0.05; **, p < 0.01; ***, p < 0.001; ****, p < 0.0001.

**SI Figure 9.**
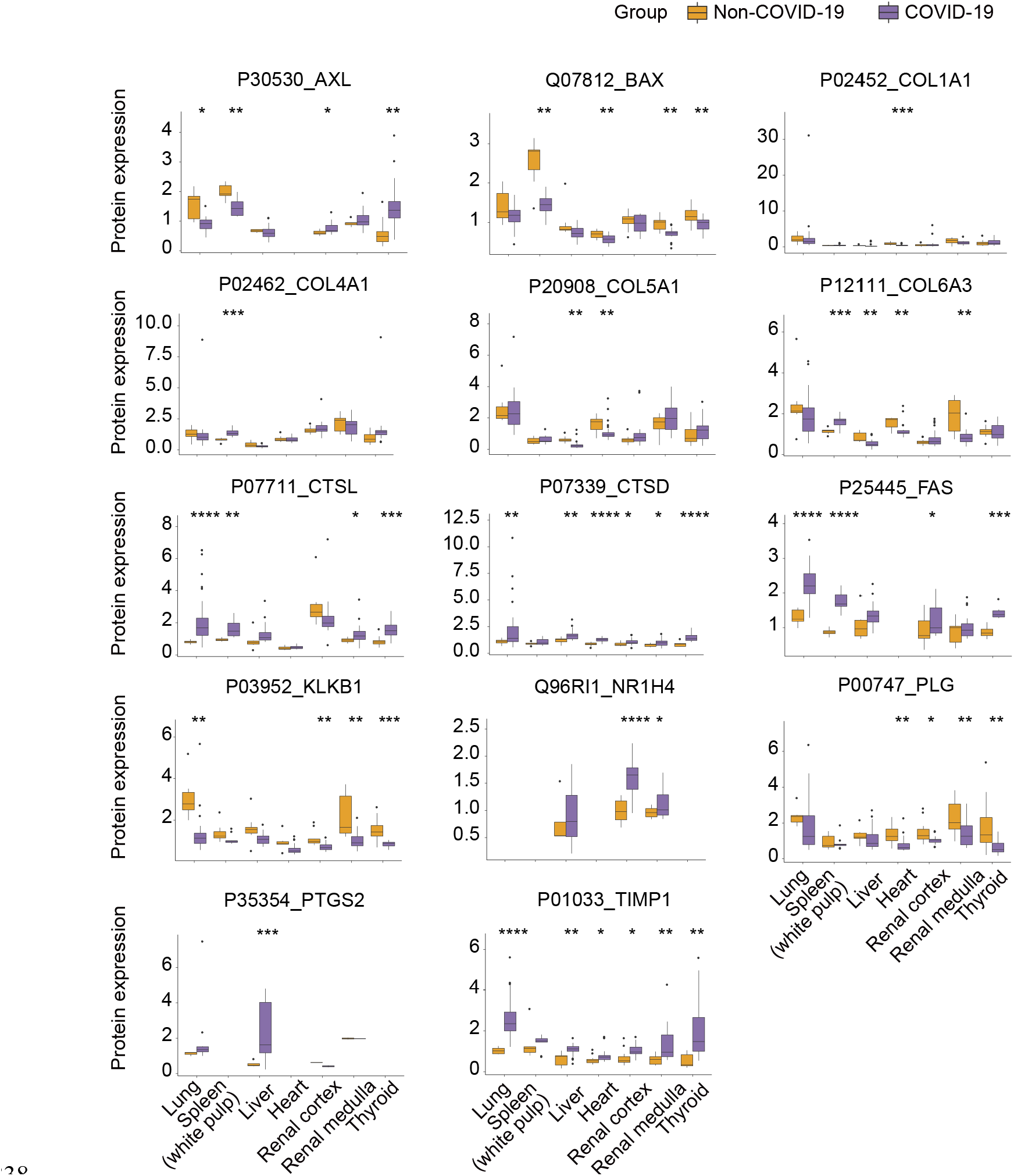
Expression of dysregulated proteins involved in modification of fibrosis process. The y-axis stands for the protein expression ratio by TMT-based quantitative proteomics. Pair-wise comparison of each protein between COVID-19 patients and control groups was performed with student’s t test. *, p < 0.05; **, p < 0.01; ***, p < 0.001; ****, p < 0.0001.

**SI Figure 10.**
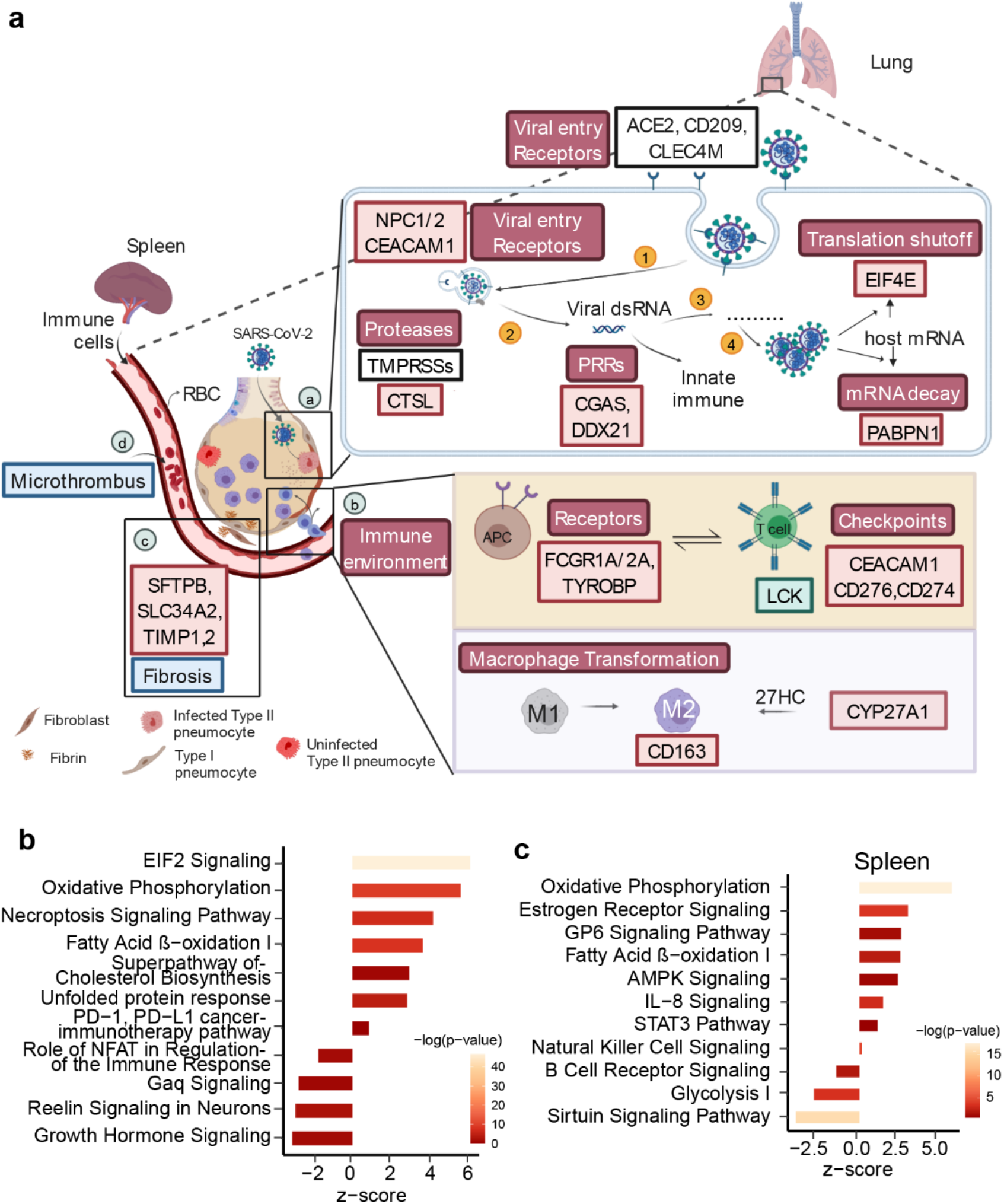
Characteristic proteins in lung and spleen of COVID-19 patients. a. Hypothetical crosstalk model between lung and spleen in COVID-19 patient. Illustration of four processes of the COVID-19 patients after SARS-CoV-2 infection including virus-host interaction, immune microenvironment of COVID-19 patients, fibrosis and microthrombus formation associated proteins. (1)-(4) in (a) shows four processes of virus infection the host cells including virus entry, virus RNA release, dsRNA translation and virus replication. Red box with black font is the upregulation protein and green box with black font is the downregulation protein. Upregulation pathway (or biological process) is red box with white font and downregulation pathway (or biological process) is green box with white font. Blue box represents the pathology of lung. **b**. Pathways enriched by IPA for 1606 pulmonary dysregulation proteins. **c**. Pathways enriched by IPA for 1726 splenic dysregulation proteins.

**SI Figure 11.**
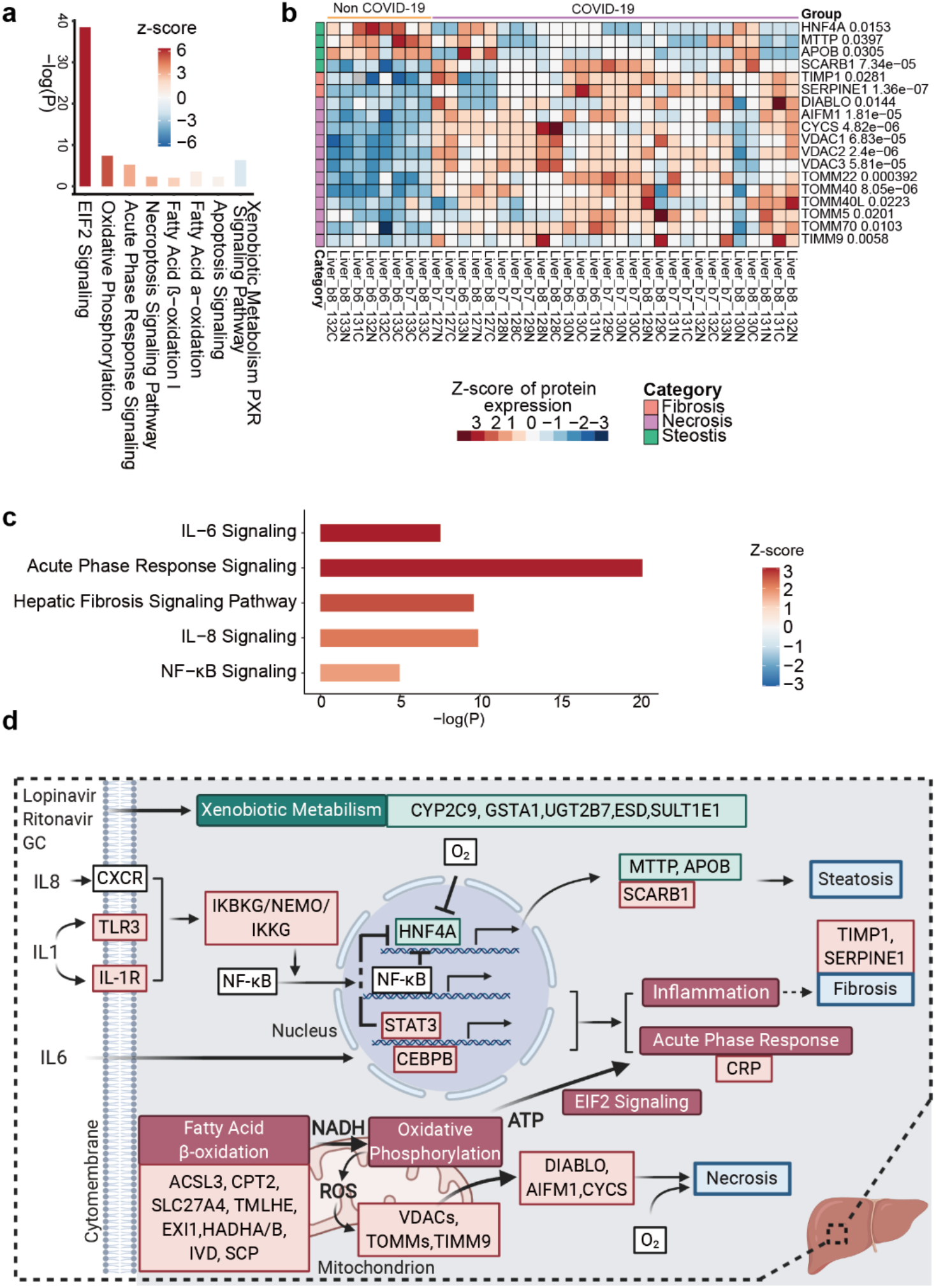
Characteristic proteins in the liver of COVID-19 patients. **a**. Pathways enriched by IPA for 1969 hepatic dysregulation proteins. **b**. Dysregulated proteins related to the pathological changes in the liver of COVID-19 patients. **c**. Pathways enriched by IPA for dysregulated immunological proteins enriched from GSEA immunologic gene sets. **d** Hypothetical model of the liver in COVID-19 patient. Red box with black font indicates the upregulation of the protein in the COVID-19 patients and green box with black font indicates the downregulation of protein. Upregulation pathway (or biological process) is shown in red box with white font and downregulation pathway (or biological process) is green box with white font. Blue box represents the pathology of liver. GC, glucocorticoid; ROS, reactive oxygen species; ATP, adenosine triphosphate; NADH, nicotinamide adenine dinucleotide.

**SI Figure 12.**
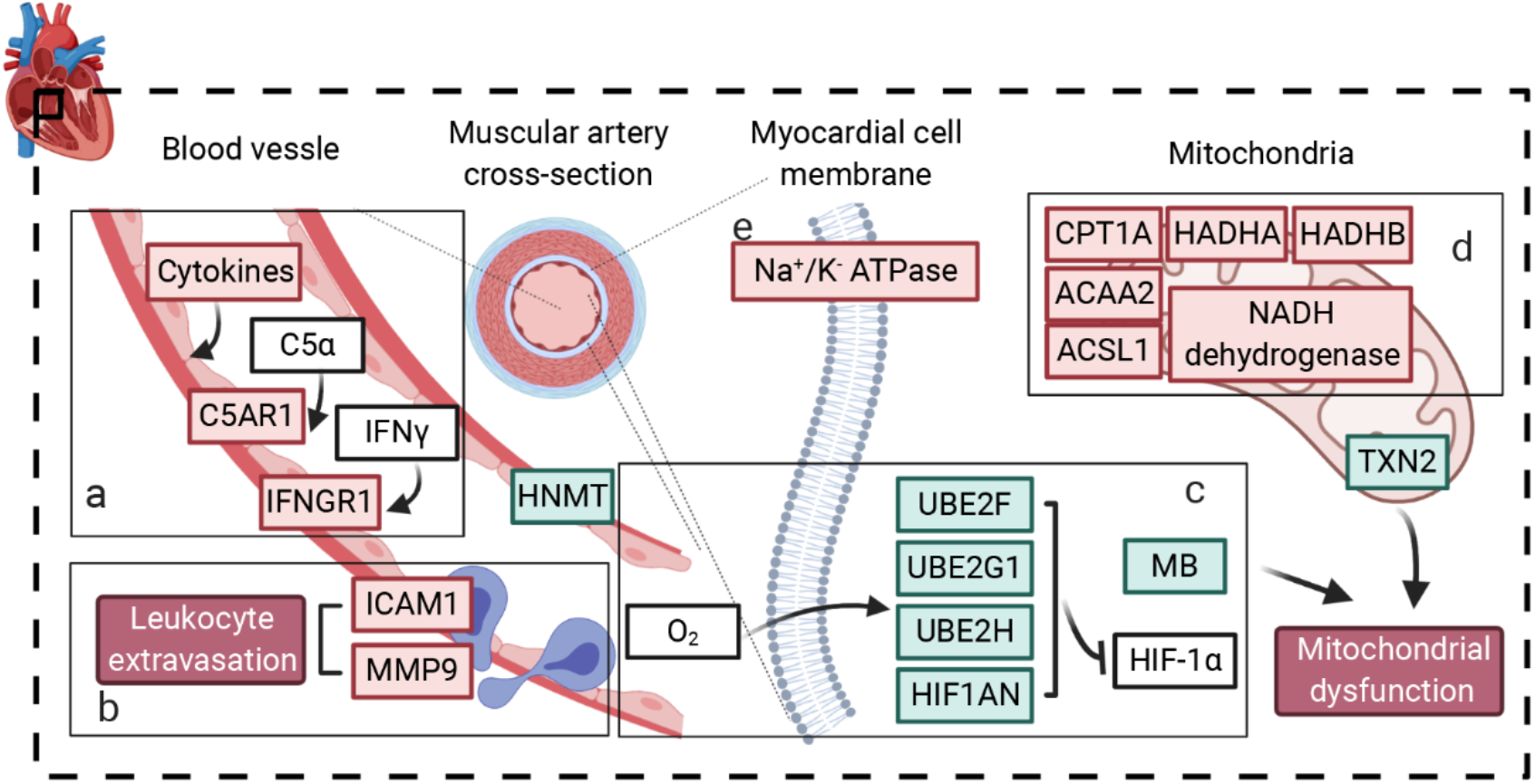
Characteristic proteins in the heart of COVID-19 patients. (a), (b) The stimulated inflammatory mediators and dysregulated endothelial cells would cause vascular hyperpermeability. (c) Hypoxia related protein alteration. (d) ATP generation in mitochondria. (e) NA+/K- ATPase dysfunction. Red box with black font is the upregulation protein and green box with black font is the downregulation protein. Upregulation pathway (or biological process) is present by red box with white font and downregulation pathway (or biological process) is green box with white font. Blue box represents the pathology of heart.

**SI Figure 13.**
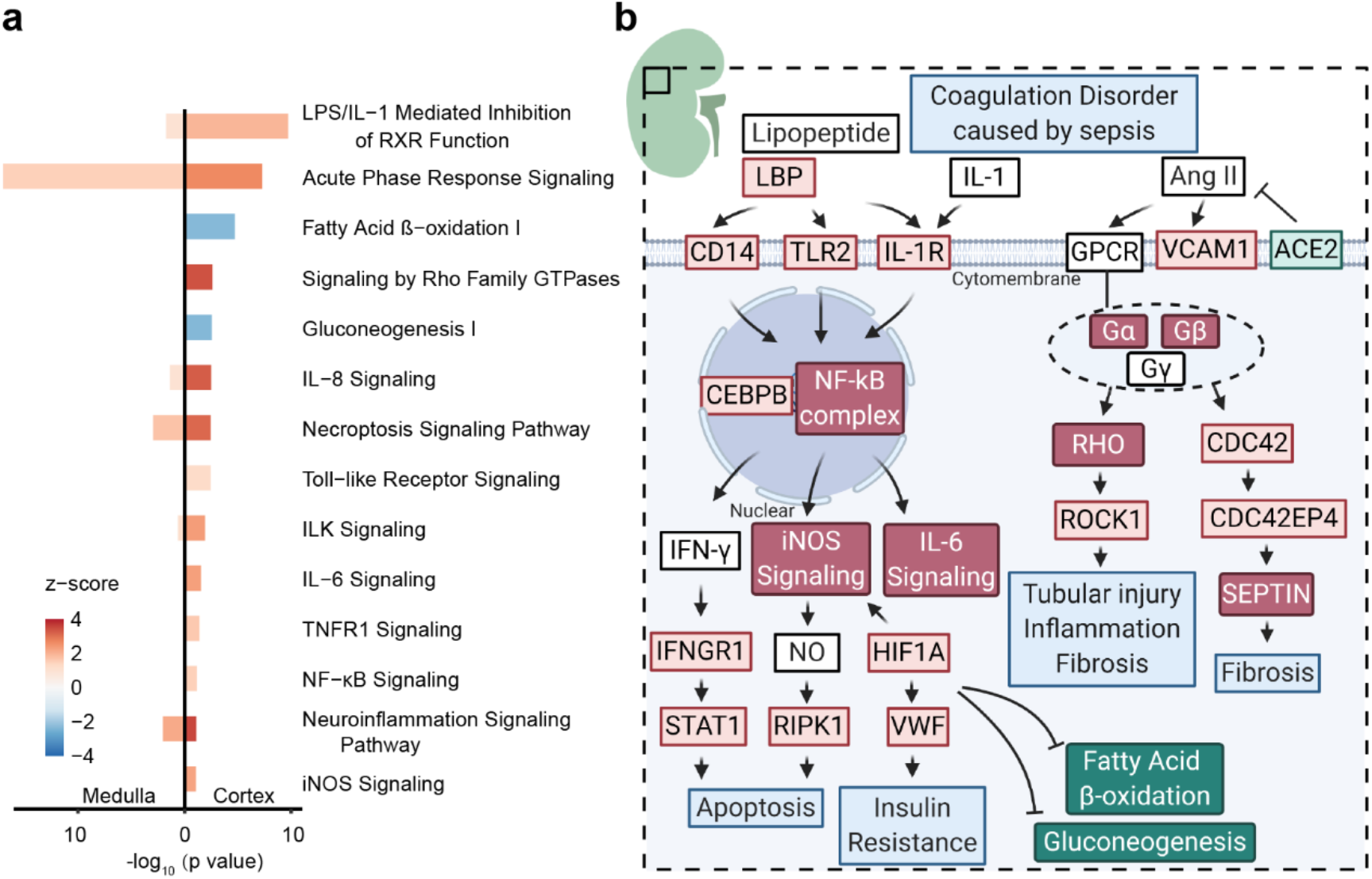
Characteristic proteins in the kidney of COVID-19 patients. **a.** Pathways enriched by IPA for 1585 dysregulated proteins in renal cortex (right) and 642 dysregulated proteins in renal medulla (left). **b**. Hypothetical model of kidney in COVID-19 patient. Red box with black font is the upregulation protein and green box with black font is the downregulation protein. Upregulation pathway (or biological process) is red box with white font and downregulation pathway (or biological process) is green box with white font. Blue box represents the pathology of kidney.

**SI Figure 14.**
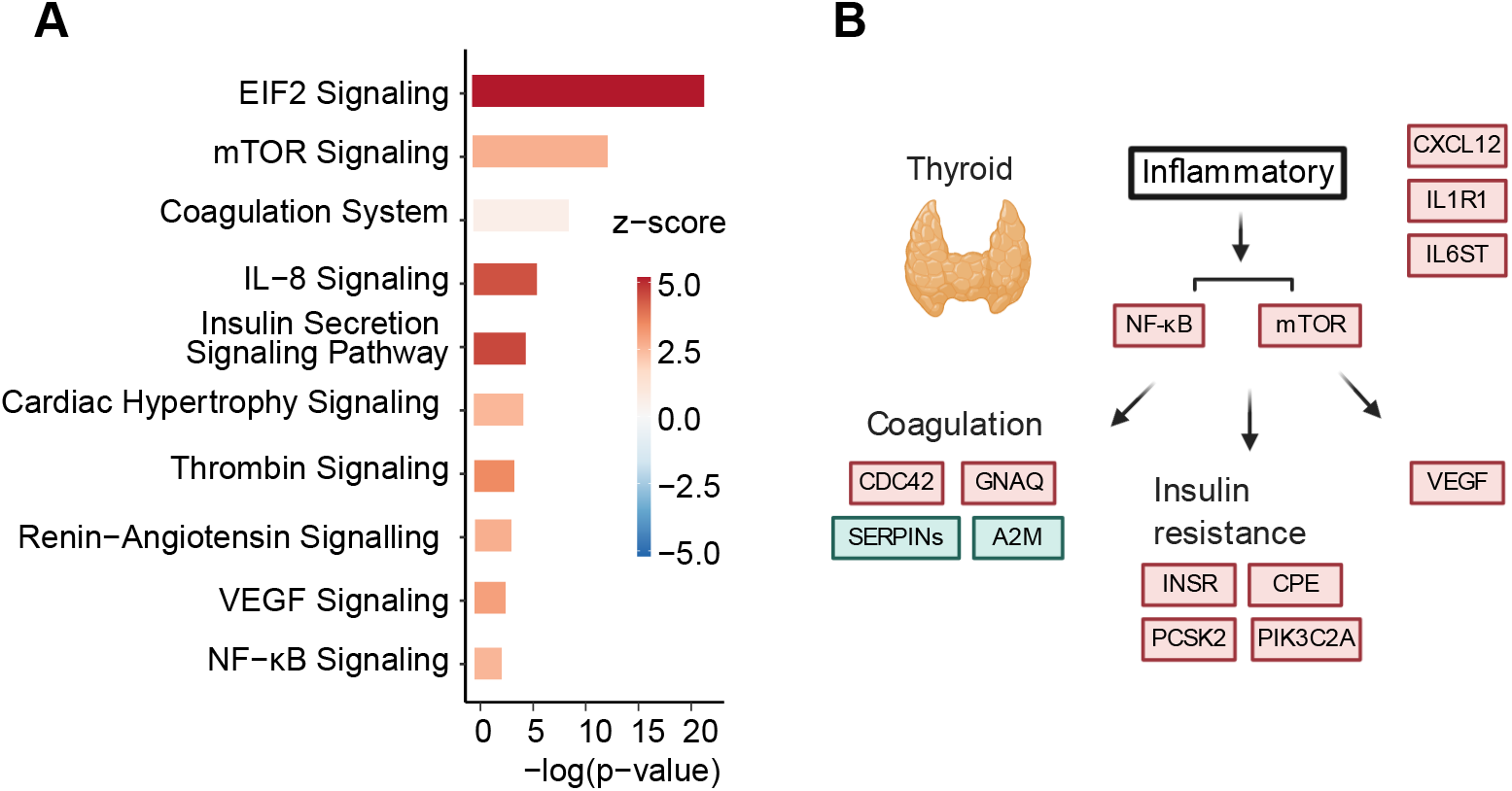
Characteristic proteins in the thyroid of COVID-19 patients. **a**. Pathways enriched by IPA for 1297 dysregulated proteins. **b**. Hypothetical model of the response of thyroid in COVID-19 patient. Red box with black font is the upregulation protein and green box with black font is the downregulation protein. Upregulation pathway (or biological process) is red box with white font and downregulation pathway (or biological process) is green box with white font.

**SI Figure 15.**
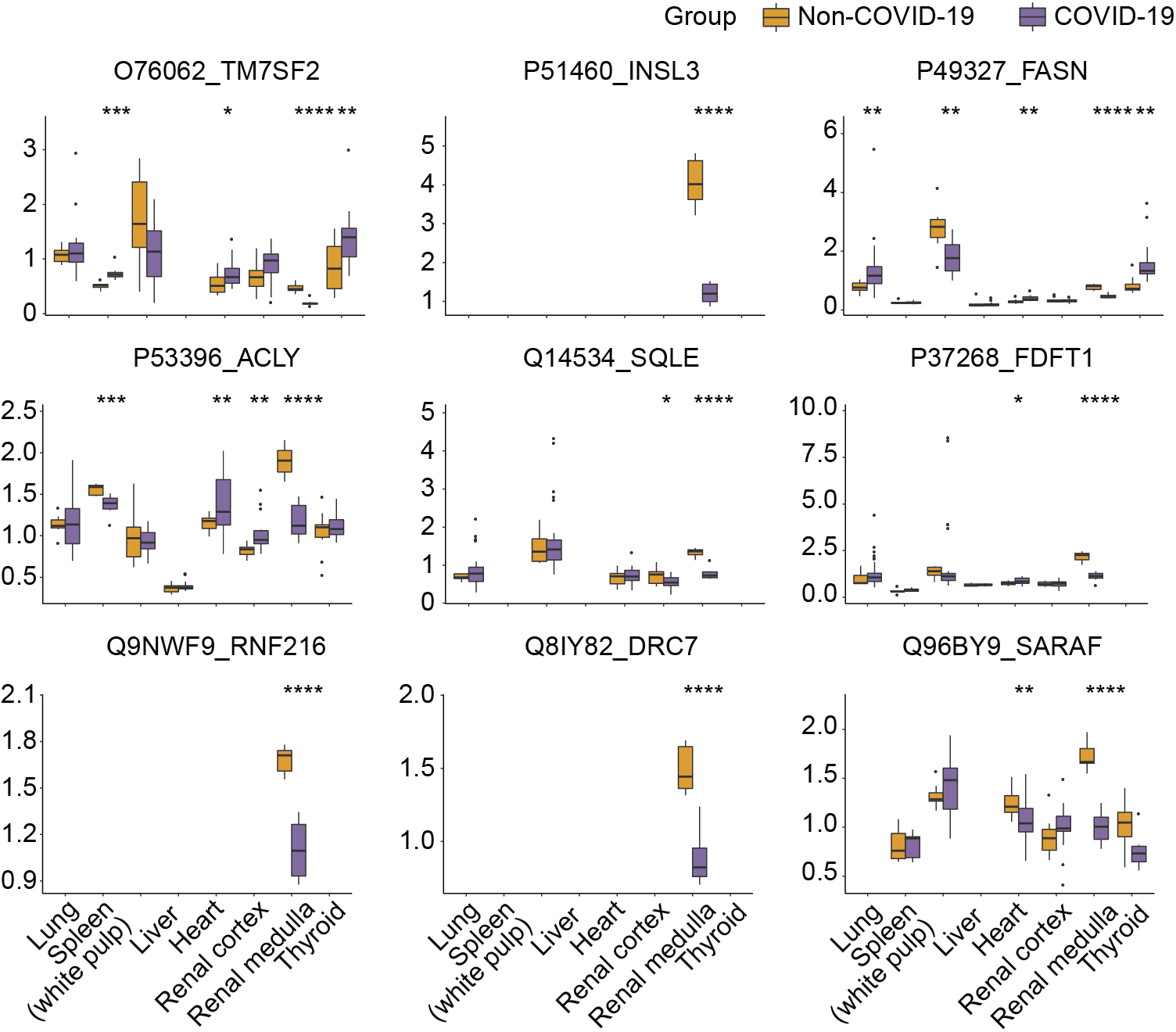
Expression of dysregulated proteins in the testis of COVID-19 patients. The y-axis stands for the protein expression ratio by TMT-based quantitative proteomics. Pairwise comparison of each protein between COVID-19 patients and control groups was performed with student’s t test. *, p < 0.05; **, p < 0.01; ***, p < 0.001; ****, p < 0.0001.

## Notes

### Competing Interest Statement

The authors have declared no competing interest.

### Author Declarations

This study was approved by the Medical Ethics Committee of Union Hospital affiliated to Tongji Medical College of Huazhong University of Science and Technology (No.2020-0043-1) and Medical Ethical Committee of Westlake University (No.20200329GTN001).

### Summary of Updates

The first name and last name were switched

